# Integrated metagenomic and bile acid metabolomic analysis of human fecal microbiota transplantation for recurrent Clostridioides difficile and/or inflammatory bowel diseases

**DOI:** 10.1101/2022.02.11.22268733

**Authors:** Ruben Jesus Faustino Ramos, Chencan Zhu, Dimitri F Joseph, Shubh D Thaker, Joseph F LaComb, Katherine Markarian, Eric R. Littmann, Hannah J Lee, Jessica C Petrov, Farah Monzur, Bradley M Morganstern, Juan Carlos Bucobo, Jonathan M Buscaglia, Anupama Chawla, Lesley Small-Harary, Grace Gathungu, Jeffrey A Morganstern, Jie Yang, Jinyu Li, Charles E Robertson, Eric G. Pamer, Daniel N Frank, Justin R Cross, Ellen Li

**Affiliations:** Donald B. and Catherine C. Marron Cancer Metabolism Center, Memorial Sloan Kettering Cancer Center, New York City, New York, United States of America; Department of Applied Mathematics and Statistics, Stony Brook University, Stony Brook, New York, United States of America; of Medicine, Stony Brook University, Stony Brook, New York, United States of America; Department of Pediatrics, Stony Brook University, Stony Brook, New York, United States of America; Department of Medicine, University of Chicago, Chicago, Illinois, United States of America; Department of Family, Population and Preventive Medicine, Stony Brook University, New York, United States of America; Biostatistics and Data Science Stony Brook Cancer Center Shared Resource, Stony Brook University, New York, United States of America; Department of Pathology, Stony Brook University, Stony Brook, New York, United States of America; Department of Medicine, University of Colorado - Denver, Aurora, Colorado, United States of America

**Author notes:** D.O-Ph.D program, Michigan State University, East Lansing, United States of America. Internal Medicine-Pediatrics Residency Program, University of Texas Southwestern, Dallas, United States of America (Hannah J Lee). Internal Medicine Residency Program, Yale University, New Haven, Connecticut, United States of America (Jessica C Petrov). New York University-Langone Hospital-Long Island, Mineola, New York, United States of America (Bradley M Morganstern). deceased (Eric R. Littmann).

## Abstract

**Background:** Fecal microbiota transplantation (FMT) is an effective treatment of recurrent *Clostridioides difficile* infections (rCDI). In comparison, FMT has more limited efficacy in treating either ulcerative colitis (UC) or Crohn’s disease (CD), two major forms of inflammatory bowel diseases (IBD). We hypothesize that FMT recipients with rCDI and/or IBD have baseline fecal bile acid (BA) compositions that differ significantly from that of their healthy donors and may be normalized by FMT.

**Aim:** To study the effect of single colonoscopic FMT on the microbial composition and function of recipients with rCDI and/or IBD.

**Methods:** Multi-omic analysis was performed on stools from 55 pairs of subjects and donors enrolled in two prospective single arm FMT clinical trials [ClinicalTrials.gov ID:NCT03268213, 479696, (IND) 15642, ClinicalTrials.gov ID: NCT03267238, IND 16795]. Fitted linear mixed models were used to examine the effects of four recipient groups (rCDI - IBD, rCDI + IBD, UC - rCDI, CD - rCDI), FMT status (Donor, pre-FMT, 1-week post-FMT, 3-months post-FMT) and first order Group*FMT interactions.

**Results:** FMT was effective in preventing rCDI for up to one year in 92% of the rCDI - IBD group and 75% of the rCDI + IBD recipients. The donor-recipient Sørensen similarity index was < 0.6 in all donor-CDI recipient pairs but > 0.6 in some donor-IBD only recipient pairs at baseline. Increasing post-FMT similarity indices > 0.6 in IBD recipients, was not clearly associated with reduced fecal calprotectin levels. Fecal secondary BA levels were lower in rCDI ± IBD and CD – rCDI recipients compared to donors. FMT restored secondary BA levels in these recipients. Metagenomic *baiE* gene and some of the 8 bile salt hydrolase (BSH) phylotype abundances were significantly correlated with fecal BA levels.

**Conclusion:** Restoration of multiple secondary BA levels, including those recently implicated in immunomodulation, are associated with restoration of fecal *baiE* gene counts, suggesting that the 7-α-dehydroxylation step is rate-limiting.

## Introduction

FMT of healthy donor stool is a highly effective treatment of rCDI [1–4]. Patients with IBD, particularly UC, indeterminate colitis (IC) and Crohn’s colitis (CC), are at increased risk of developing CDI [5–9], and FMT is an effective treatment of rCDI in these patients [10–15]. However, studies thus far on the effectiveness of FMT in the treatment of IBD, absent concomitant CDI, have utilized different protocols and have yielded mixed results [16–27].

Restoration of bacterial metabolism of primary BA to secondary has been implicated in the mechanism by which FMT promotes resistance to CDI [28]. Primary conjugated BA synthesized in the liver are released into the gut lumen upon feeding and serve to emulsify dietary fats. Within the gut lumen, conjugated BA undergo deconjugation by bacterial bile salt hydrolases (BSH) in the small intestine [29]. While most of BA from the small intestine by enterohepatic circulation and transported back to the liver, a fraction reaches the colon and undergoes further conversion by bacterial enzymes, such as 7-alpha dehydroxylase, to form secondary BA. 7-alpha dehydroxylating bacteria, such as *Clostridium scindens*, have been shown to be reduced in rCDI patients and replenishment of these bacteria was demonstrated to inhibit CDI in an animal model [28]. More recently, epimers of deoxycholic and lithocholic acids (e.g. iso-deoxycholic, 3-oxo-lithocholic, isoallo-lithocholic acids) have been demonstrated to have *in vitro* immunomodulatory activity [30–32]. Modified BA pool composition has been observed in IBD patients [33–35], with altered levels of some BA epimers that exhibit potential immunomodulatory effects.

At Stony Brook University Medical Center (SBUMC), we have been conducting two prospective single arm clinical trials: 1.) ClinicalTrials.gov ID:NCT03268213, 479696, UC only Investigational New Drug (IND) 15642 and 2.) ClinicalTrials.gov ID: NCT03267238, CD only IND 16795, to examine the longitudinal microbiological and clinical effects of single donor FMT on the fecal microbiome in four groups of recipients: rCDI without IBD (rCDI – IBD); rCDI with IBD (rCDI + IBD); UC without rCDI (UC – rCDI); CD without rCDI (CD – rCDI, see Fig 1). Each FMT recipient selected their own donor, who was subsequently screened for pathogens and antibiotic resistant bacteria. This FMT study is unique in that it includes prospective collection of a relatively large set of healthy donor stools. The results of 16S rRNA sequence analysis of single donor colonoscopic FMT performed on a subset of 19 recipients was previously reported [36], which included: 1) 11 rCDI – IBD; 2) 3 rCDI + UC; and 3) 5 UC - rCDI. The previous results indicated marked changes in β diversity (i.e. overall microbiome composition differences between groups) in the rCDI + UC and rCDI – IBD groups (compared to donors). The rCDI - IBD and rCDI + UC recipients had a marked decreased abundances of anaerobic taxa in the Ruminococcaceae, (e.g. Faecalibacterium) and Lachnospiraceae groups, and marked increased abundances of microaerophilic taxa in the Bacilli group, and Proteobacteria and Fusobacteria phyla, which were restored post FMT. In contrast the pre-FMT differences in β-diversity were much more subtle in the UC - rCDI group compared to their healthy donors. These differences were characterized by increased abundances of microaerophilic taxa in the Bacilli, Proteobacteria and Fusobacteria in the UC - rCDI group.

**Fig 1.**
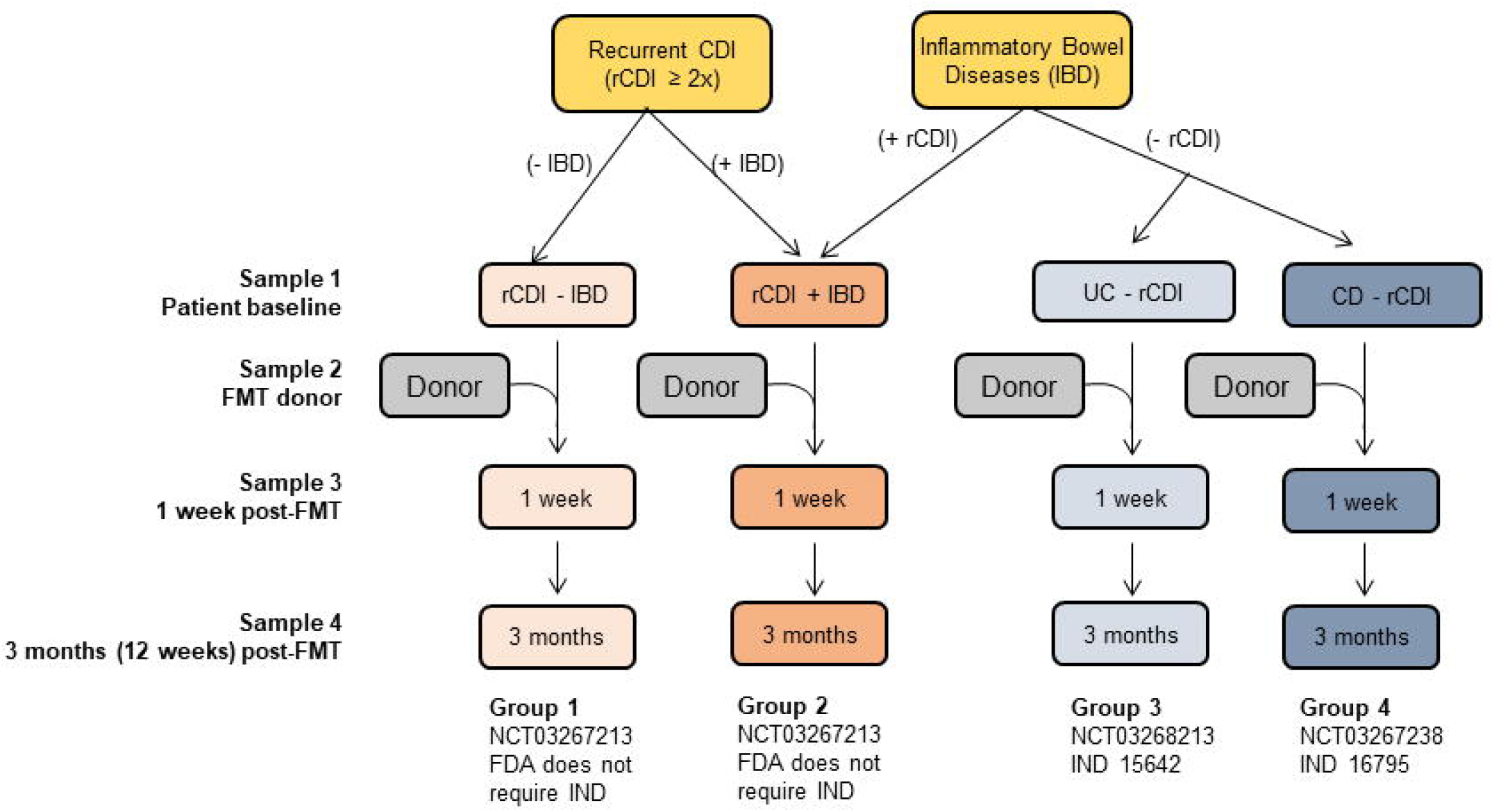
Schematic diagram of recipient group and donor fecal sample collection.

We now report the results of multi-omic analysis (16S rRNA gene sequencing, metabolomics analysis and shotgun DNA metagenomics sequencing) of the effect of colonoscopic single donor FMT in an expanded set of recipients separated into four categories: 1.) rCDI - IBD; 2.) rCDI + IBD; 3.) UC - rCDI; 4.) CD - rCDI (see Fig 1).

## Materials and methods

### Study protocols

The patient and donors were recruited through two open label study protocols (See S1-S2 text for IRB approved protocols). ClinicalTrials.gov ID NCT03268213, 479696, UC no rCDI ≥ 2x IND 15642, was approved by the Stony Brook Institutional Review Board (479696) on 11/14/2013. ClinicalTrials.gov ID NCT03267238, IND 16795, was approved by Stony Brook Institutional Review Board (973349) on 06/16/2017 (see S1 Table Transparent Reporting of Evaluations with Nonrandomized Designs (TREND) checklist). The initial recipient/donor pair was recruited on 12/05/2013. The final recipient/donor included in this report were recruited 06/19/2019. There was a delay in the registration of NCT03268213 on ClinicalTrials.gov relative to the initial recruitment of the first recipient/donor pairs as described previously [37], but no delay in the registration of NCT03267238 relative to the initial recruitment of patient/donor pairs. For the patients with rCDI (with or without IBD), the inclusion criteria were ≥ 2 recurrences despite treatment with antibiotics, documented by ≥ 3 positive stool tests for CDI (see Fig 1). For the patients with UC without a history of rCDI ≥ 2x, the inclusion criterion was medication refractory UC, requiring step up therapy beyond mesalamine alone. Only one of the patients in this group had a distant prior history of CDI with the last episode 2 years prior to the procedure. For the patients with CD without a history of rCDI ≥ 2x, the inclusion criterion was medication refractory CD, requiring step up therapy beyond mesalamine alone. None of the CD patients had a history of previous CDI. Exclusion criteria for all of the recipients included: a) scheduled for abdominal surgery within 12 weeks of the study, b) pregnancy, c) Grade 4 anemia (Hemoglobin < 6 g/dL), d) Grade 1 neutropenia (Absolute Neutrophil Count <1500), e) known diagnosis of graft vs. host disease, f) major abdominal surgery within the past 3 months, g) administration of any investigational drug within the past 2 months., h) use of a TNF-α antagonist within 2 weeks of the proposed date of transplantation, i) bacteremia within past 4 weeks. (28 days), and j.) adults ≥18 years unable to or unwilling to give informed consent. The CONSORT flow diagram is outlined in Fig 2. Recruitment and collection of data and processing of patient samples were conducted as previously described in Mintz *et al.* [36]. The IBD subphenotypes were classified using the Montreal classification in terms of disease location (L1, ileal CD; L2, Crohn’s colitis; L3, ileocolitis; L4, upper gastrointestinal) for CD and extent of disease (E1, proctitis; E2, left-sided UC; E3, extensive UC proximal to splenic flexure) for UC [37]. Mucosal disease activity in the colon and the terminal ileum at the time of the colonoscopic FMT was scored using the Mayo endoscopic subscore for UC - rCDI recipients [38] and the simple endoscopic score for CD - rCDI (SES-CD) [39].

**Fig 2.**
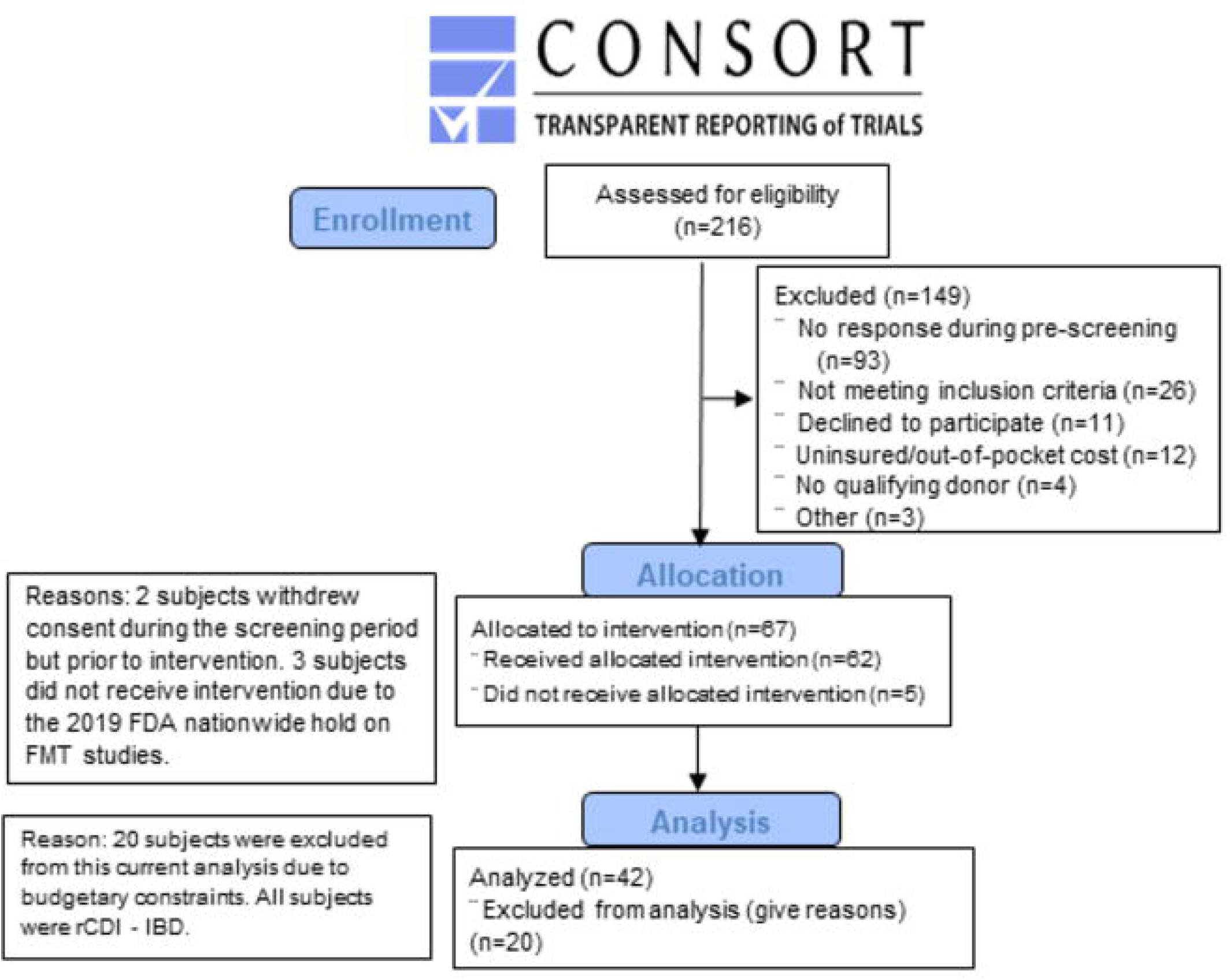
CONSORT 2010 flow diagram.

### Dietary information

The recipients and donors were asked to record daily dietary intake on the Stony Brook University Medical Center Department of Pediatrics Food Diary/Calorie Count for 7 days prior to the collection of each stool sample. Patients were grouped into three categories on review of their dietary logs and interviews: 1.) animal protein-meat (mostly standard “western” diet); 2.) vegetarian (predominantly plant based but animal protein from eggs and dairy); and 3.) plant based protein source (vegan but could include honey).

### Fecal calprotectin analysis

Levels of the neutrophilic fecal calprotectin protein, a clinical marker of gut mucosal inflammation, was measured using the PhiCal Test (Calpro AS, Norway) enzyme-linked immunosorbent assay (ELISA) following the manufacturer’s recommended protocol as previously described [36].

### 16S rRNA sequence analysis

16S rRNA sequencing targeting the V3V4 variable region of the 16S rRNA gene: primers 338F (5’ ACTCCTACGGGAGGCAGCAG) and 806R (5’ GGACTACHVGGGTWTCTAAT) was carried out by the Frank laboratory at the U of Colorado as previously described [36]. All de-multiplexed, paired-end 16S rRNA sequencing data along with the metadata were deposited into the NCBI Sequence Read Archive under BioProject accession number PRJNA739170.

### BA metabolomics analysis by liquid chromatography-mass spectrometry (LC-MS)

Human stool samples (∼400 mg) were weighed into 7 mL bead tubes containing 1.4 mm ceramic beads (Omni International) and extracted using 80% methanol containing deuterated labeled internal standards [ISTDS: chenodeoxycholic acid–d4 (CDCA-d4),taurochenodeoxycholic acid-d4 (T-CDCA-d4), glycochenodeoxycholic acid-d4 (G-CDCA-d4), cholic acid-d4 (CA-d4), taurocholic acid-d4 (T-CA-d4), glycocholic acid-d4 (G-CA-d4), deoxycholic acid-d4 (DCA-d4), taurodeoxycholic acid-d4 (T-DCA-d4), glycodeoxycholic acid-d4 (G-DCA-d4), lithocholic acid-d4 (LCA-d4), tauro-lithocholic acid (T-LCA-d4), glycolithocholic acid-d4 (G-LCA-d4), ursodeoxycholic acid-d4 (UDCA-d4), T-UDCA-d4, and β-muricholic acid-d5 (β-MCA-d5)] for a final concentration of 100 mg/mL (see S2 Table). The ISTDs were purchased from Cambridge Isotope Laboratories (Tewksbury, MA); except β-MCA-d5 that was purchased from Isosciences (Ambler, PA). Samples were homogenized using a Bead Ruptor (Omni International, Kennesaw, GA) at 6 m/s for 6 cycles of 30 s at 4°C, then stored at −80°C overnight. Stool extracts were then vortexed, transferred to 5 mL Eppendorf tubes, centrifuged at 20,000 × g for 20 min at 4°C and diluted with Milli-Q water to 50% methanol content. Samples were then filtered using 96-well Acroprep Advance plates (Pall Corporation, Port Washington, NY) and analyzed by LC-MS analysis BA were separated using two chromatographic methods: 1.) Agilent 1290 Infinity LC system coupled to an Agilent 6550 iFunnel Q-TOF with a 1.6 μm, 2.1 × 50 mm CORTECS T3 column (Waters) and 2.) Agilent 1290 Infinity II LC system coupled to an Agilent 6546 Q-TOF with a 1.7 μm, 2.1 × 150 mm ACQUITY UPLC BEH Shield RP18 column (Waters). Active reference mass correction was used according to the manufacturer’s instructions. Mobile phase A was 0.1% formic acid in water, mobile phase B was 0.1% formic acid in acetone. LC gradient for Method 1: 30% B to 65% B over 14 min (flow rate: 0.4 mL/min) and for Method 2: 30% B to 80% B over 30 min (flow rate: 0.3 mL/min). Both methods ended with 100% B for 2 min and re-equilibration for 6 min. Acquisition was from m/z 50-1,700 at 2 Hz. Mass spectrometer source conditions for both methods: gas temperature: 240°C; drying gas flow: 10 L/min; nebulizer: 60 psi; sheath gas temperature and flow: 400°C and 10 L/min, respectively. The voltages used were: fragmentor: 175 V; skimmer: 45 V; capillary: 4000 V; and nozzle: 500 V. Sample injection volume was 5 µL, and data was collected in negative ionization mode. A pooled sample was injected regularly throughout the batch to monitor instrument performance. Data files were converted to Agilent SureMass format and analyzed in Agilent MassHunter Quantitative Analysis software (version 10.1, Agilent Technologies, Santa Clara, CA). Concentrations were estimated by referencing peak areas to a specified ISTDs of known concentration (S2 Table), except for sulfated-BAs and 3-oxo-LCA, which used an external calibration curve.

### Shotgun DNA metagenomics analysis

Shotgun DNA metagenomics sequencing was carried out on fecal DNA samples at Novogene Corp Inc. (Beijing, China). DNA samples were assessed using Qubit Fluorimeter while agarose gel electrophoresis was carried out to check for DNA degradation. The NEBNext Ultra^TM^II DNA Library PrepKit for Illumina® (Ipswich, MA) was used to prepare the sequencing libraries according to the manufacturer’s protocols and Illumina 150 bp paired end (PE150) sequencing was conducted on the HiSeqX instruments with a targeted depth of 20 million. Quality filtering (e.g., removal of adapters, human sequences, low-quality sequences) was conducted using KneadData version 5.1 (http://huttenhower.sph.harvard.edu/kneaddata) as previously described [33]. All de-multiplexed, paired-end shotgun DNA metagenomic sequencing data along with the metadata were deposited into the Sequence Read Archive under BioProject accession number PRJNA739170. Reads corresponding to BA metabolizing enzymes were identified by a translated search with DIAMOND [40] and a manually curated protein sequence database (see S3 Table), respectively at two threshold levels: 1.) 80% identity over 80% sequence and 2.) 50% identity over 80% sequence. Reads corresponding to antibiotic resistance genes were identified using the Comprehensive Antimicrobial Resistance Database (CARD).

### Statistical analysis

Baseline patient characteristics were compared between the donors and the four groups of FMT recipients Alpha diversity indices (e.g. Chao1, Shannon complexity H, Shannon Evenness H/Hmax) were calculated inferred through 1000 replicate resamplings using Explicet [41], and β-diversity (Bray-Curtis distances) were calculated between the recipient samples and their paired donor samples as previously described [36]. The Sørensen similarity score is defined as 1-Bray-Curtis index, and 0.6 was selected as the threshold level for successful implantation of the donor microbiota [27]. P-values less than 0.05 were considered as statistically significant. Because many of the 385 operational taxonomic units (OTUs) exhibited zero counts across multiple libraries, generalized linear mixed models (GLMM) or generalized estimating equation (GEE) models were used to analyze 108 OTUs after eliminating OTUs with an average relative abundance of < 0.001% in the donor and recipient pre-FMT samples, and after discarding OTUs where more than 75% of the samples had a zero count. To compare the relative abundance of each OTU between donors and recipient time points before and after FMT (pre-transplant, 1 week post-FMT, 3 months post-FMT) within each disease group (rCDI - IBD, rCDI + IBD, UC - rCDI, CD - rCDI), GLMMs or GEEs were used by taking the actual counts of each OTU as the outcomes that were assumed to follow a negative binomial distribution [36]. The log-transformed overall sequence count for each individual at each time point was considered as an offset. Two-way interaction terms (Group*FMT) were used to estimate the difference between the time points within a specific disease group. Possible covariance structures to model correlation among longitudinal measurement from the same patient and measurement in the corresponding donor were unstructured (UN) and compound symmetry (CS). In GEE, the dependence structure was chosen based on Quasi Information Criteria (QIC). Pair-wise P-values were based on the T-test for GLMM, and the Z-test for GEE. The P-values were adjusted for multiple comparisons by the Bonferroni correction (P threshold = 0.05/number of multiple comparison) or by the Benjamini-Hochberg method (false discovery rate or FDR < 0.05) as described further in the text. Kruskal-Wallis tests were used to compare BA levels between donor and four pre-FMT recipient groups. Post-hoc Dunn’s tests with Benjamini-Hochberg adjustments were then performed in comparison of recipient groups and donor. Linear mixed models were used to estimate the pairwise BA differences between pre-FMT recipient and donor as well as response post-FMT in each disease group. UN or CS covariance structures were selected to model the correlation among longitudinal measurements from the same patient and his/her corresponding donor based on (Aikaike Information Criteria (AIC). Log-transformation was applied if the normality assumption was not satisfied. If log-transformation was performed, pre-FMT recipient/donor ratios were shown; otherwise, donor-pre-FMT recipient differences were shown. Kruskal-Wallis tests were also used to compare bacterial gene abundances (cpm) at 80% identity between donor and four pre-FMT recipient groups. Estimated correlation coefficients were generated from linear mixed models treating FMT groups as clustering effect, in order to assess the correlations between bacterial gene abundances (cpm). Log transformation was applied to bacterial gene abundances if normality assumption was not met. All analysis was performed in SAS 9.4 (SAS institute Inc., Cary, NC) and R 3.6.1 (R Foundation for Statistical Computing, Vienna, Austria).

## Results

### Patient characteristics of FMT donors and recipient samples analyzed

The characteristics of the 55 recipient/donor pairs, segregated according to donors and the four groups of recipients, are summarized in Table 1. The median age in the rCDI – IBD group was significantly higher than the donor and the other three recipient groups. Significant differences in the proportion of males between groups was also observed, being particularly high in the UC - rCDI and CD - rCDI groups. Donors and recipients in all groups had predominantly White/European Ancestry (EA) and no Black/African Ancestry (AA) subjects. While only a minority of patients were current smokers, there were differences in the proportion of former smokers and never smokers between groups. All of the rCDI recipient groups received antibiotics within a month prior to the FMT in order to prevent recurrence of CDI, whereas none of the donors (within 3 months of donating the stool sample), UC - rCDI or CD - rCDI recipients (within one month of undergoing FMT) received antibiotics. 20% of rCDI - IBD recipients and 56-62% of the IBD recipients with or without rCDI had abnormal fecal calprotectin levels (> 120 µ*g/g*). Of the 43 donors measured, only two had borderline values (> 50 µ*g/g*, ≤ 120 µ*g/g*) and the rest had normal values (≤ 50 µ*g/g*).

**Table 1.** Baseline characteristics of donors and the four groups of FMT recipients. The median values and interquartile range (IQR) are listed for each group. Fecal calprotectin ≤ 50 *µg/g*, normal; 50.1-120 *µg/g*, borderline; > 120 *µg/g*, abnormal.

| Variable | Donor<br>n = 55 | Group 1<br>rCDI - IBD<br>n = 27 | Group 2<br>rCDI + IBD<br>n = 8 | Group 3<br>UC - rCDI<br>n = 11 | Group 4<br>CD - rCDI<br>n = 9 |
| --- | --- | --- | --- | --- | --- |
| Age (y) ± IQR | 43 ± 25 | 66 ± 21 | 37 ± 29 | 37 ± 18 | 18 ± 4 |
| **Sex n (%)** |  |  |  |  |  |
| Males | 23 (42%) | 8 (30%) | 3 (38%) | 6 (54%) | 8 (89%) |
| **Race n (%)** |  |  |  |  |  |
| White/EA | 52 (95%) | 27 (100%) | 8 (100%) | 10 (82%) | 7 (78%) |
| Black/AA | 0 (0%) | 0 (0%) | 0 (0%) | 0 (0%) | 0 (0%) |
| Asian | 3 (5%) | 0 (0%) | 0 (0%) | 2 (18%) | 2 (22%) |
| **Ethnicity** |  |  |  |  |  |
| Hispanic n (%) | 8 (15%) | 2 (7%) | 2 (25%) | 2 (18%) | 0 (0%) |
| **Smoking n (%)** |  |  |  |  |  |
| Current | 5 (9%) | 4 (15%) | 0 (0%) | 1 (9%) | 0 (0%) |
| Former | 9 (16%) | 13 (48%) | 2 (25%) | 4 (36%) | 1 (11%) |
| Never | 41 (75%) | 10 (37%) | 6 (75%) | 6 (55%) | 8 (89%) |
| **Body mass index (BMI)** |  |  |  |  |  |
| **kg/m<sup>2</sup> ± IQR** | 23.8 ± 7 | 27.1 ± 7.6 | 23 ± 7 | 27 ± 9 | 20 ± 5 |
| **Diet n (%)** |  |  |  |  |  |
| Meat | 49 (89%) | 27 (100%) | 8 (100%) | 10 (91%) | 8 (89%) |
| Vegetarian (dairy) | 5 (9%) | 0 (0%) | 0 (0%) | 1 (9%) | 1 (11%) |
| Plant based | 1 (2%) | 0 (0%) | 0 (0%) | 0 (0%) | 0 (0%) |
| **Antibiotics n (%)** |  |  |  |  |  |
| **within 3 month** | 0 (0%) | 27 (100%) | 8 (100%) | 1 (9%) | 0 (0%) |
| **IBD medications n (%)** |  |  |  |  |  |
| 5-ASA | 0 (0%) | 0 (0%) | 6 (75%) | 7 (64%) | 5 (56%) |
| Steroids | 0 (0%) | 0 (0%) | 1 (12%) | 4 (36%) | 3 (33%) |
| Immunomodulators | 0 (0%) | 0 (0%) | 5 (62%) | 4 (36%) | 4 (44%) |
| Biologics | 0 (0%) | 0 (0%) | 2 (25%) | 4 (36%) | 4 (44%) |
| None | 55 (100%) | 27 (100%) | 1 (12%) | 1 (12%) | 2 (22%) |
| **Fecal calprotectin n (%)** |  |  |  |  |  |
| Abnormal | 0 (0%) | 6 (22%) | 5 (62%) | 7 (64%) | 5 (56%) |
| Borderline | 2 (4%) | 6 (22%) | 3 (38%) | 2 (18%) | 0 (0%) |
| Normal | 41 (74%) | 15 (56%) | 0 (0%) | 2 (18%) | 3 (33%) |
| Missing | 12 (22%) | 0 (0%) | 0 (0%) | 0 (0%) | 1 (11%) |

### Clinical outcomes for the four recipient groups

The primary endpoint for rCDI was defined as no further rCDI at 12 months after FMT in rCDI recipients with or without IBD. FMT was 92% effective in preventing rCDI in the rCDI - IBD group and 75% effective in the rCDI + IBD group. These results compare favorably to the previously reported 91% efficacy in in donor FMT compared to 67% effectiveness of the sham FMT [2] and previously reported uncontrolled FMT studies performed in rCDI patients with IBD [10–15].

The primary endpoint for gut inflammation as measured by fecal calprotectin levels was one week after FMT for all recipients because many IBD patients were prescribed changes in their medications between one week and 3 month time points. We did not observe a clear effect of a single colonoscopic FMT on gut inflammation in IBD recipients as measured by decreased fecal calprotectin.

Serious adverse events (SAEs) were reported in six patients as listed in S4-S7 Tables. The SAEs in three rCDI – IBD recipients and one UC - rCDI recipient, were deemed unrelated to the FMT procedure on review by the Data Safety Monitoring Board. The remaining two SAEs observed in one rCDI + IBD (CD) and one CD – rCDI recipient could have reflected progression of their underlying disease but could also have been related to the procedure, since flares have been previously reported in other IBD patients post FMT [12].

### 16S rRNA gene sequencing results

16S rRNA gene profiling of fecal bacterial communities was accomplished for 165 fecal DNA samples, collected from 43 of 55 donors, 18 of 27 rCDI - IBD recipients, 8 of 8 rCDI + IBD recipients, 9 of 11 UC - rCDI recipients and 8 of 9 CD - rCDI recipients.

Linear mixed models comparing the ShannonH α-diversity index between donor and recipient pairs in each of the four groups (Fig 3) revealed significant differences between the donor group and both the rCDI - IBD (*P* = 0.0001) and rCDI + IBD (p <0.0001) groups at baseline (Pre-FMT), with resolution of these differences at the 1 week and three month time points. In contrast, differences in α-diversity between donor and recipient did not reach significance for either the UC - rCDI or CD - rCDI groups.

**Fig 3.**
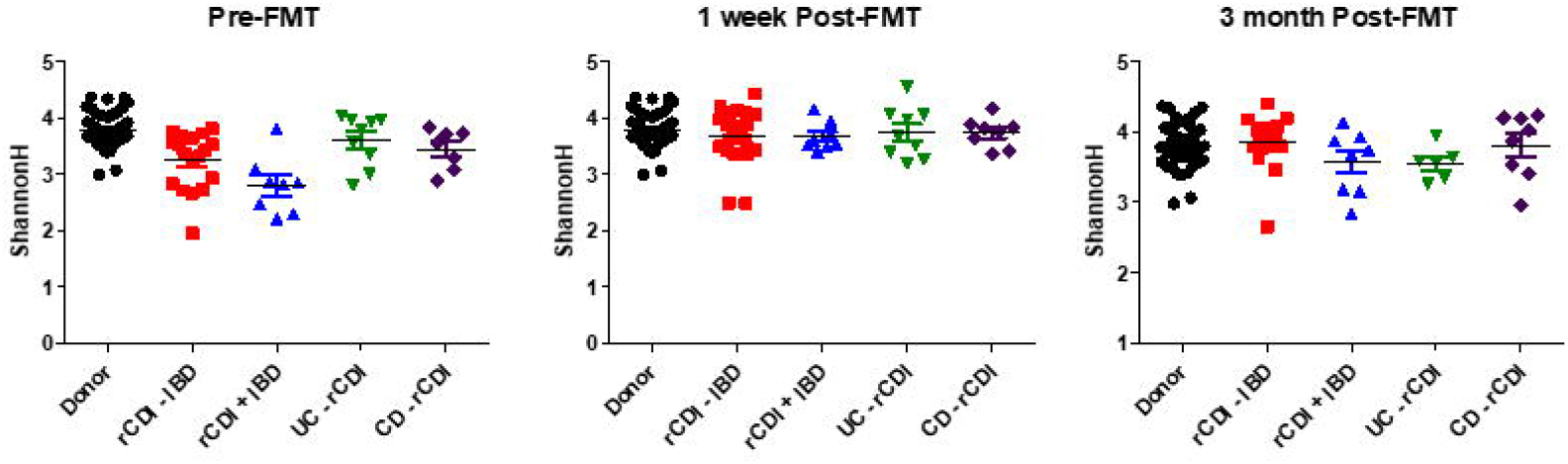
α-diversity (ShannonH) in the donor and four recipient groups Pre-FMT, 1 week Post-FMT and 3 month Post-FMT.

A recent report of a clinical FMT trial in CD patients [27] set successful implantation of donor stools as achieving a Sørensen similarity index > 0.6 (the maximum value of 1 indicates identical communities) between paired donors and recipients. We therefore adopted the same endpoint in analyzing our 16S rRNA gene sequencing results. The pre-FMT Sørensen similarity indices were below the endpoint in <u>al</u>l of the rCDI recipients with and without IBD (Table 2), with overall increases in these indices post-FMT. Individually the endpoint of > 0.6 was reached post FMT in 73%of the rCDI recipients. Among the IBD - rCDI groups prior to FMT, 5 of 9 (56%) UC - rCDI recipients and 5 of 7 (71%) CD – rCDI recipients had donor-recipient pre-FMT Sørensen similarity indices below the endpoint. Three of these recipients had evidence of active disease based on their initial fecal calprotectin level and endoscopic score (Mayo or SES-CD). Three of the 5 UC – rCDI donor-recipient pairs and 3 of 5 5 CD - rCDI recipients reached the endpoint post-FMT at either the 1 week and/or 3 month time point. However, achieving this endpoint was not necessarily associated with reduction of gut inflammation measured using fecal calprotectin levels. Note that <u>none of the CD recipients</u> in the previously published sham controlled FMT study of CD recipients achieved a Sørensen similarity index > 0.6 at the 6 week post-FMT time point [27].

**Table 2.** Median Sørensen similarity index ± IQR between paired donors and recipients at Pre-FMT, 1 week post FMT and 3 months post FMT.

| Group | Pre-FMT | 1 week Post-FMT | 3 month Post-FMT |
| --- | --- | --- | --- |
| **rCDI - IBD**<br>(n =18) | 0.26 ± 0.28 | 0.65 ± 0.09 | 0.66 ± 0.16<br>(3 missing) |
| **rCDI + IBD**<br>(n = 8) | 0.08 ± 0.26 | 0.55 ± 0.28 | 0.49 ± 0.15 |
| **UC - rCDI**<br>(n = 9) | 0.54 ± 0.27 | 0.62 ± 0.14 | 0.61 ± 0.22<br>(3 missing) |
| **CD - rCDI**<br>(n = 8) | 0.52 ± 0.14<br>(1 missing) | 0.62 ± 0.19 | 0.63 ± 0.17 |

We next analyzed how abundances of individual bacterial taxa (from 16S rRNA gene sequencing) differed between recipient groups (rCDI – IBD, rCDI + IBD, UC – rCDI, CD – rCDI), and healthy donors as previously described [36]. Pairwise comparisons of the estimated recipient/donor ratios were then carried out. 72 OTUs demonstrating significant estimated pre-FMT recipient/donor ratios with 95%CI in at least one of the four recipient groups with a FDR (q) <0.05 are listed in S7 – S14 Tables and selected taxa are highlighted in Table 3.

**Table 3.**
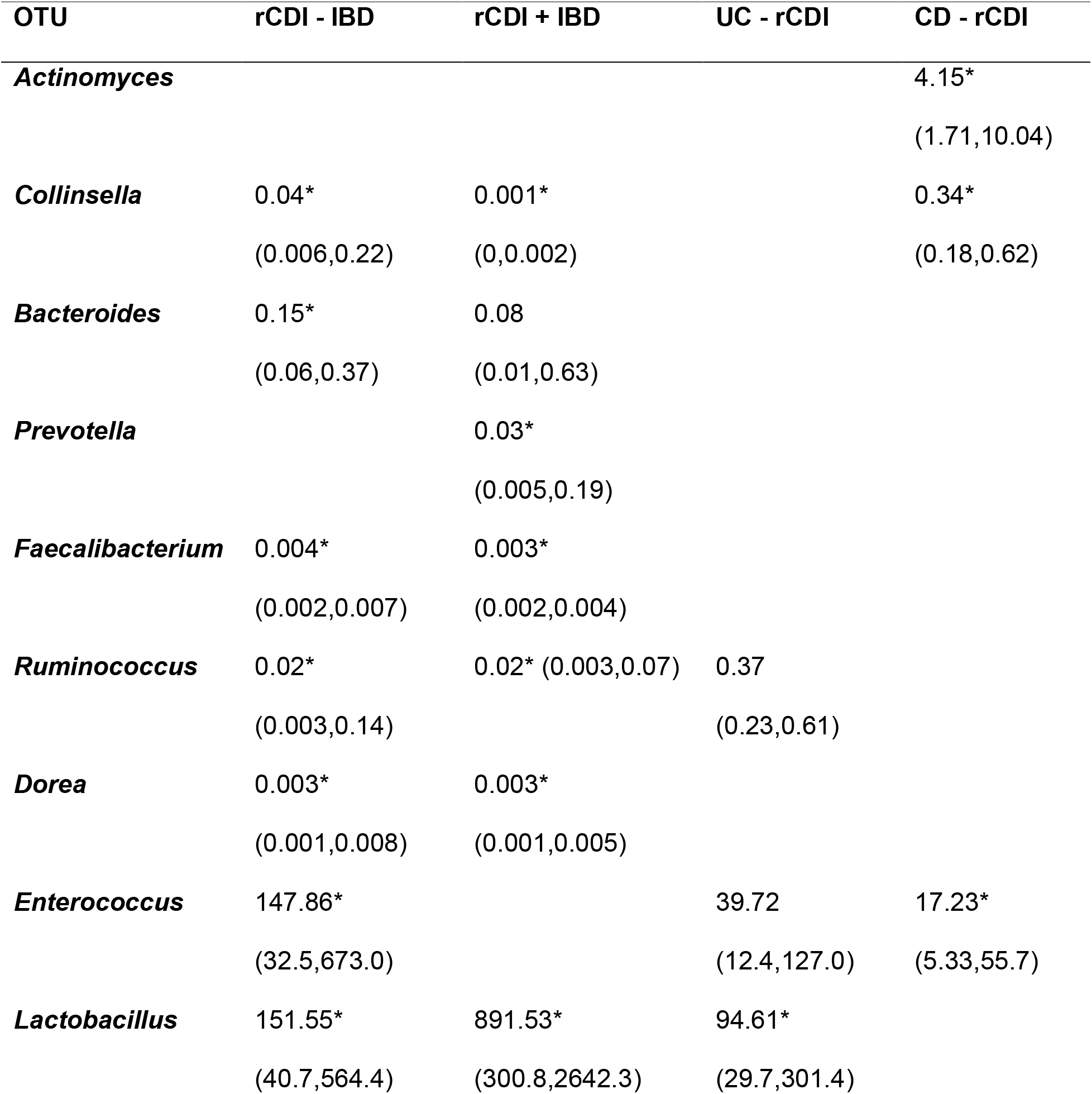
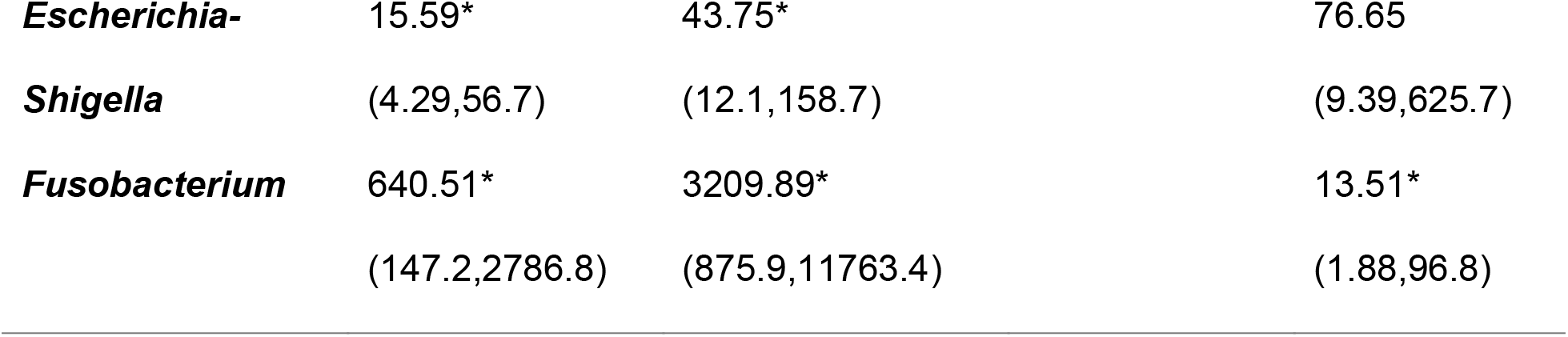
Significant (FDR <0.05) estimated pre-FMT recipient/donor ratios (95% CI) of the relative abundances of selected genus level OTUs for each of the four recipient groups. See also S7 – S14 Tables). A ratio > 1 indicates ↑ relative abundance in recipient pre-FMT compared to donor samples. A ratio of < 1 indicated ↓ relative abundance in pre-FMT samples compared to donor samples. The 95% CI are shown in parenthesis. Only the estimated ratios with an FDR < 0.05 are listed. The * indicates those OTUs that also had significant estimated ratios of post-FMT/pre-FMT relative abundances at with 1 week and/ or 3 month time point, indicating that FMT had a significant effect.

The expanded dataset with 16S rRNA gene sequence data generated from 7 additional recipients in the rCDI – IBD group, confirmed 47 of 51 OTUs previously reported [36] with a significant estimated pre-FMT recipient/donor ratio. The expanded dataset for the rCDI + IBD recipients generated from 5 additional recipients, including 3 with underlying CD [36] confirmed 54 of 71 OTUs previously reported with a significant estimated pre-FMT recipient/donor ratio. The expanded dataset for the UC -rCDI recipients generated from 4 additional recipients [36], confirmed only 15 of 29 OTUs previously reported with a significant estimated pre-FMT recipient/donor ratio. Increased recipient/donor ratios for Bacilli, Proteobacteria and Fusobacteria OTUs were among those confirmed for the UC - rCDI group. The results of 16S rRNA gene sequence analysis for the fourth recipient group, CD - rCDI, detected significant estimated pre-FMT recipient/donor ratios in 27 OTUs, including decreased ratios in Actinobacteria, Bacteroides and *Ruminococcaceae* OTUs and increased ratios in Bacilli, Gamma-proteobacteria and Fusobacteria OTUs.

### Quantitative targeted analysis of BA metabolites

We initially observed relative peak differences in BA on a subset of samples, which suggested that secondary BA metabolites were reduced not only in a subset of the rCDI – IBD and rCDI + IBD groups [36], but also in the CD - rCDI group (data not shown). To further pursue these findings, 23 bile acid metabolites including secondary BA epimers (see Fig 4), which have recently been implicated in regulating immune function [30–32], were quantitatively measured in 212 samples in 55 donor/recipient pairs by targeted LC-MS (S15 Table). Prior to FMT, 19 BA metabolites differed significantly from the donor group by a post-hoc Dunns test with Benjamini-Hochberg adjustments (see S16 Table). The unconjugated secondary BA, lithocholic acid (LCA) and deoxycholic acid (DCA), were the predominant metabolites in the healthy donors, but low levels of secondary bile acid epimers, 3-oxo-lithocholic acid (3-oxo-LCA), iso-lithocholic acid (iso-LCA) and isoallo-lithocholic acid (isoallo-LCA) were also detected (see Fig 4 and 5). In contrast, the unconjugated primary BA, chenodeoxycholic acid (CDCA) and cholic acid (CA), were the predominant metabolites detected in the rCDI-IBD recipients, with significant reduction of secondary BA and epimers. For the rCDI + IBD recipients, in addition to the increased levels of primary unconjugated BA (CDCA and CA) and reduced secondary BA levels, increased levels of <u>conjugated</u> primary BA, taurocholic acid (T-CA), glycochenodeoxycholic acid (G-CDCA) and glycocholic acid (G-CA), were also observed. The BA metabolite profile for the UC - rCDI recipients was very similar to that of the healthy donors, whereas a reduction of unconjugated secondary BA and epimers was observed for the CD - rCDI recipients (Fig 5).

**Fig 4.**
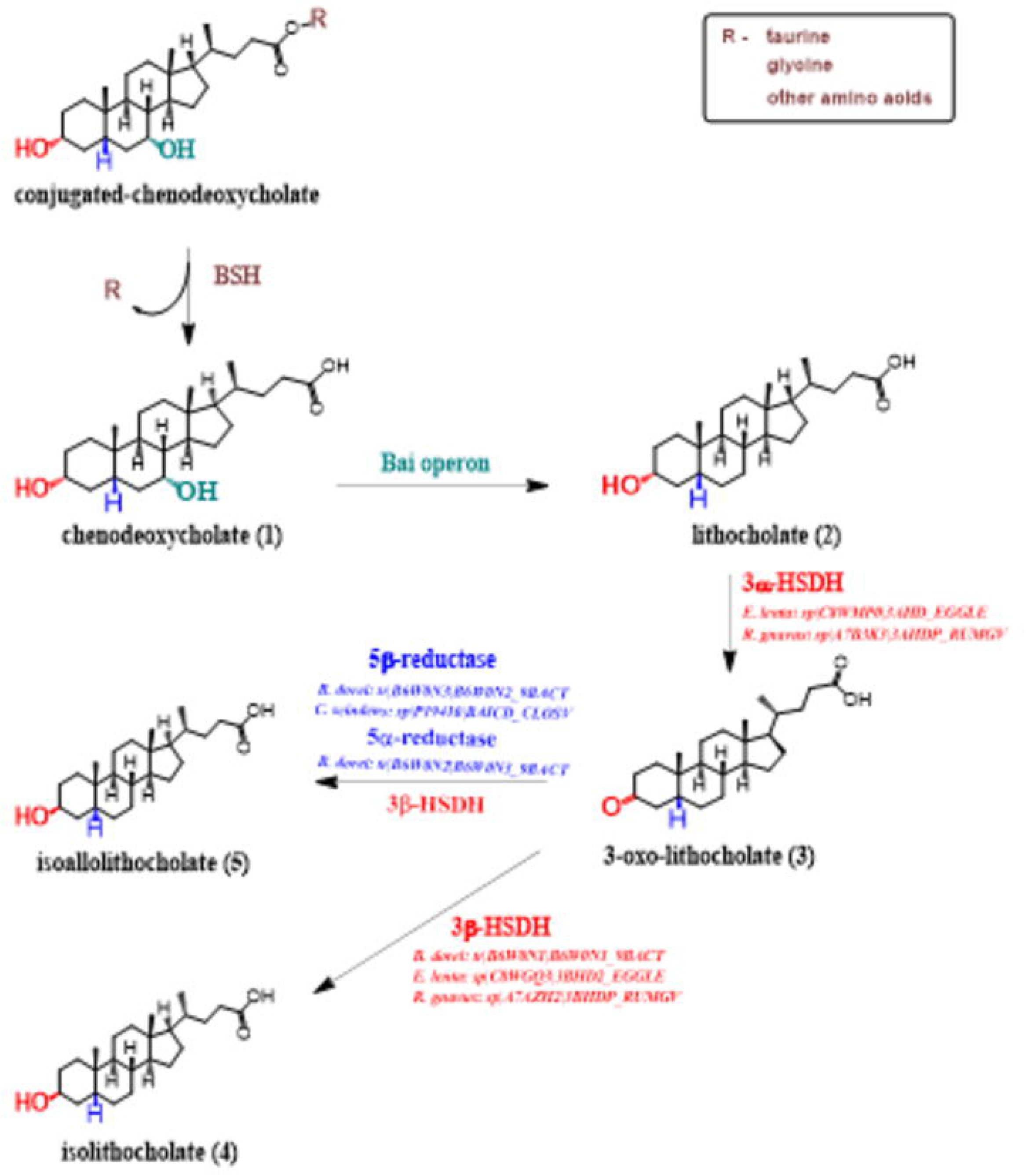
BA metabolic pathways. Chenodeoxycholate (CDCA) is synthesized in the liver and conjugated with taurine and glycine before being secreted to the small intestine. Taurochenodeoxycholate (T-CDCA) and glycochenodeoxycholate (G-CDCA) are then deamidated to chenodeoxycholate (CDCA) (1) by gut bacteria containing the bile salt hydrolase (BSH) enzyme. CDCA can be further 7α-dehydroxylated to lithocholic acid (LCA) (2) by the concerted action of 6 enzymes in the *bai* operon (*baiA2, baiB, baiCD, baiE, baiF,* and *baiH*). LCA can be further transformed to 3-oxo-LCA (3) by action of 3α-hydroxysteroid dehydrogenases (3α-HSDH), and transformed to iso-LCA (4) if the ketone at position C3 of 3-oxo-LCA is reduced to a 3β-alcohol, by action of 3β-HSDH, or (5) isoallo-LCA by the concerted action of 5β/α-reductases and 3β-HSDH.

**Fig 5.**
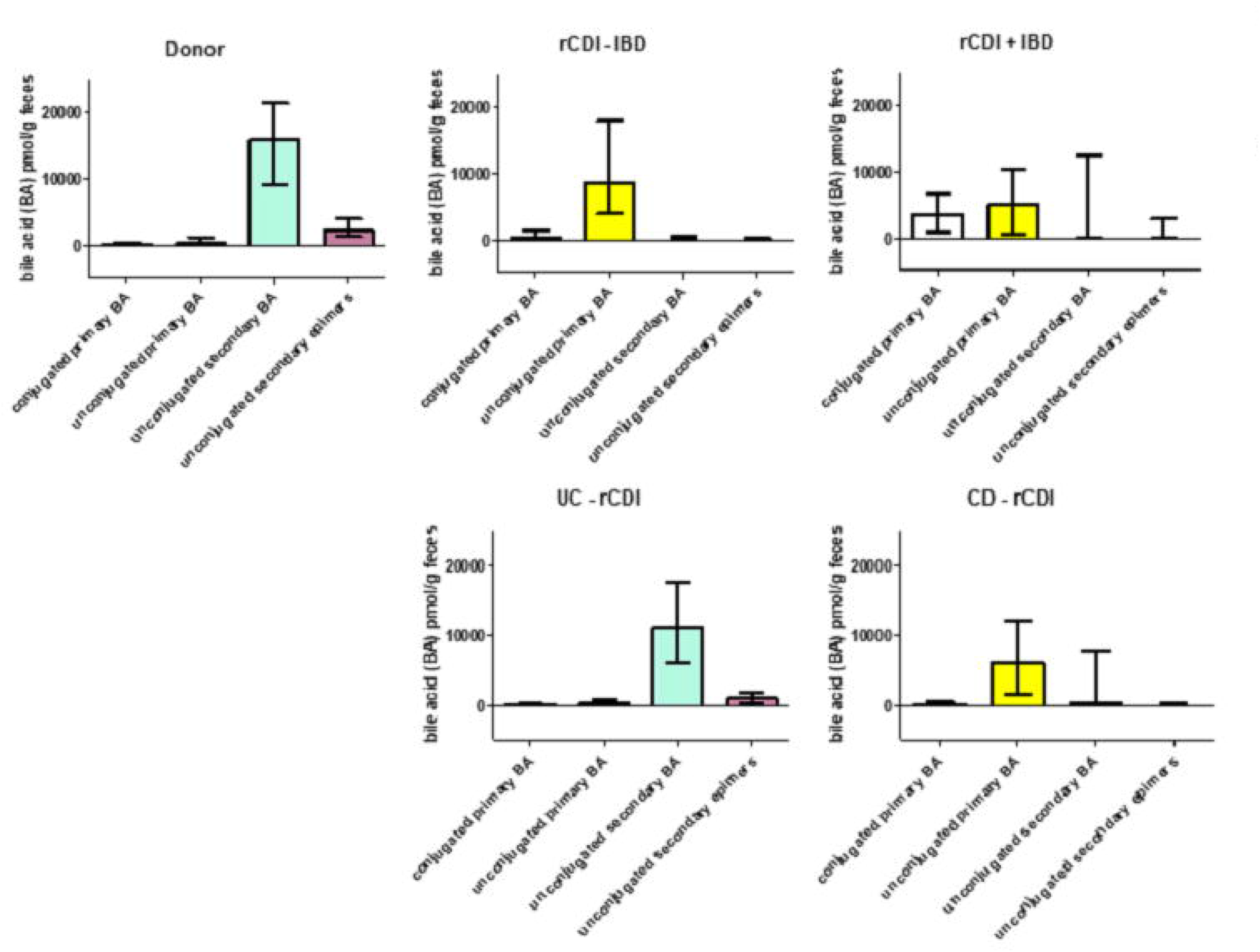
Fecal BA metabolites in donors and the four recipient groups at baseline. The total median ± IQR concentrations of conjugated primary BAs (T-CA, G-CDCA, G-CA), unconjugated primary BAs (CDCA, CA), unconjugated secondary BAs (LCA, DCA) and unconjugated secondary BA epimers implicated in immune regulation (3-oxo-LCA, iso-LCA, isoallo-LCA) are shown.

In order to measure the effect of FMT on the BA metabolites, fitted linear mixed models with pairwise comparisons were conducted for 16 BA metabolites that required log transformation to measure the estimated pre-FMT recipient donor ratios and response post-FMT (see Table 4). Fitted linear mixed models with pair wise comparisons were conducted for the remaining 3 BA metabolites that did not require log transformation to measure the estimated donor-pre-FMT recipient differences and response post-FMT (see Table 5). These results indicate that FMT has a significant effect on restoring secondary BA metabolite levels in rCDI – IBD, rCDI + IBD and CD - rCDI recipients.

**Table 4.**
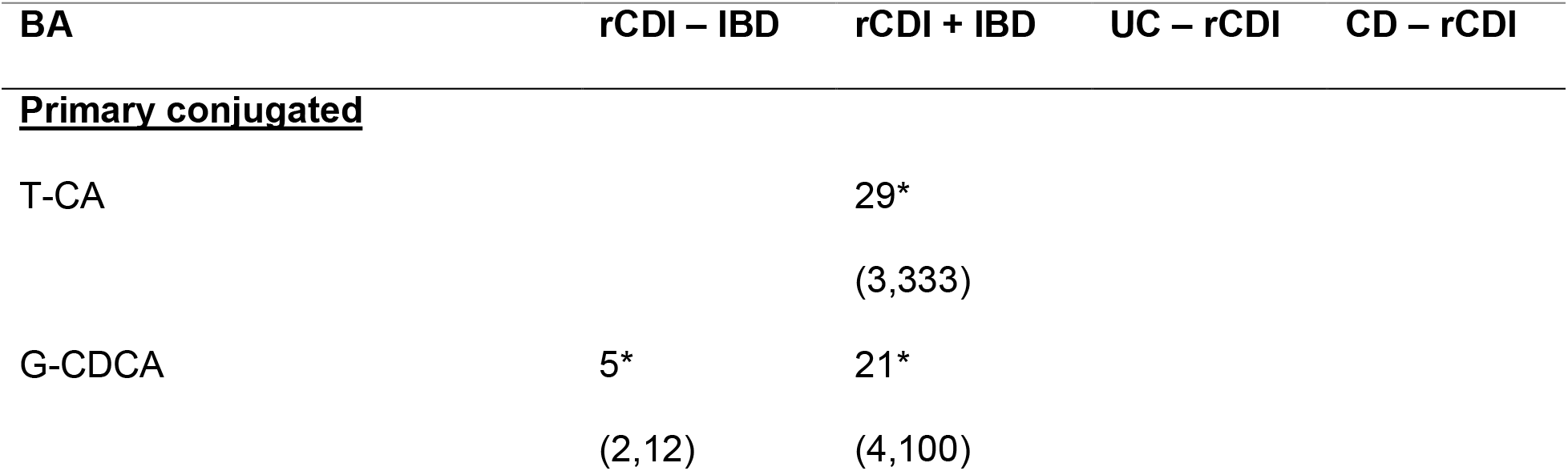
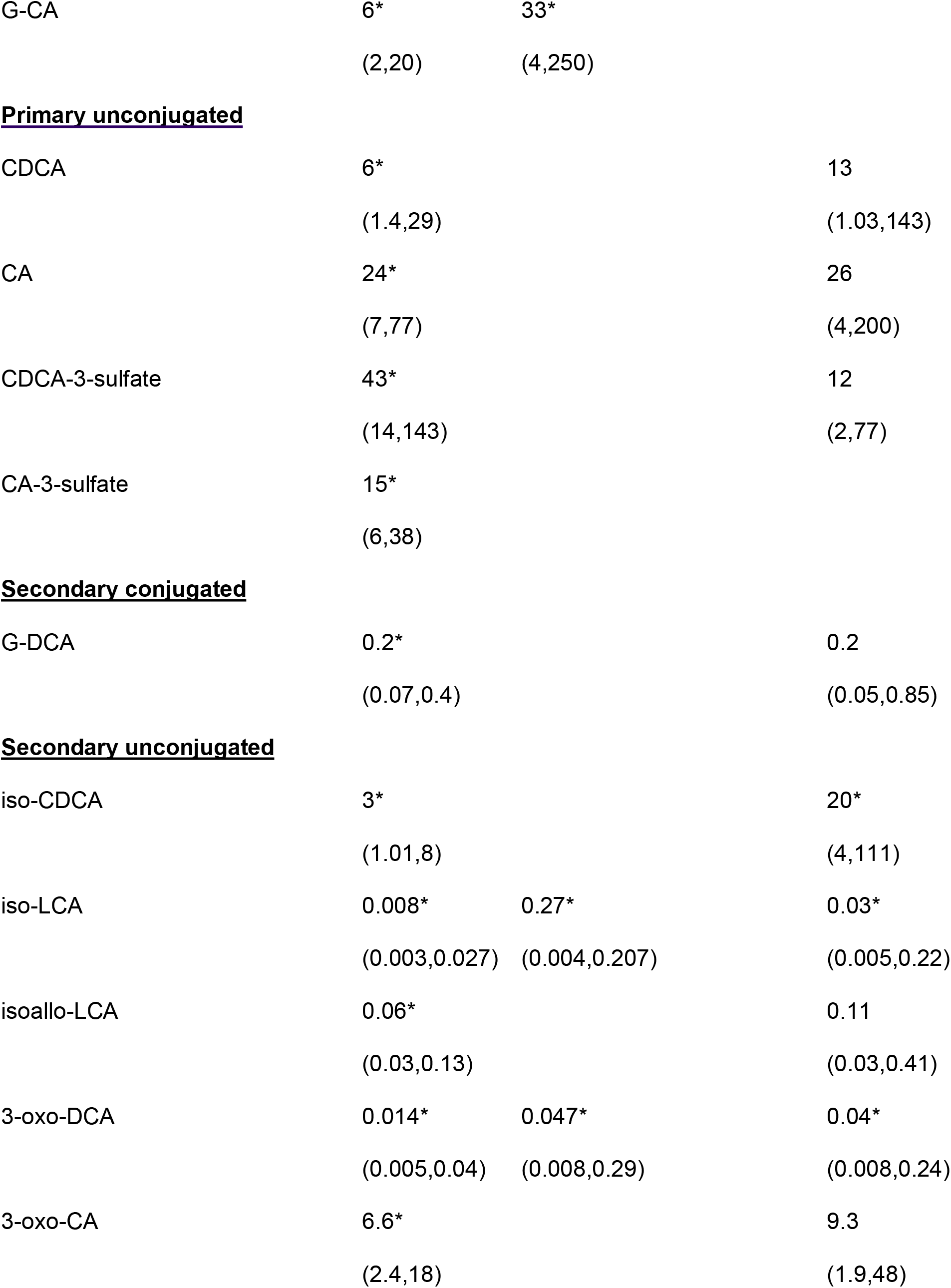
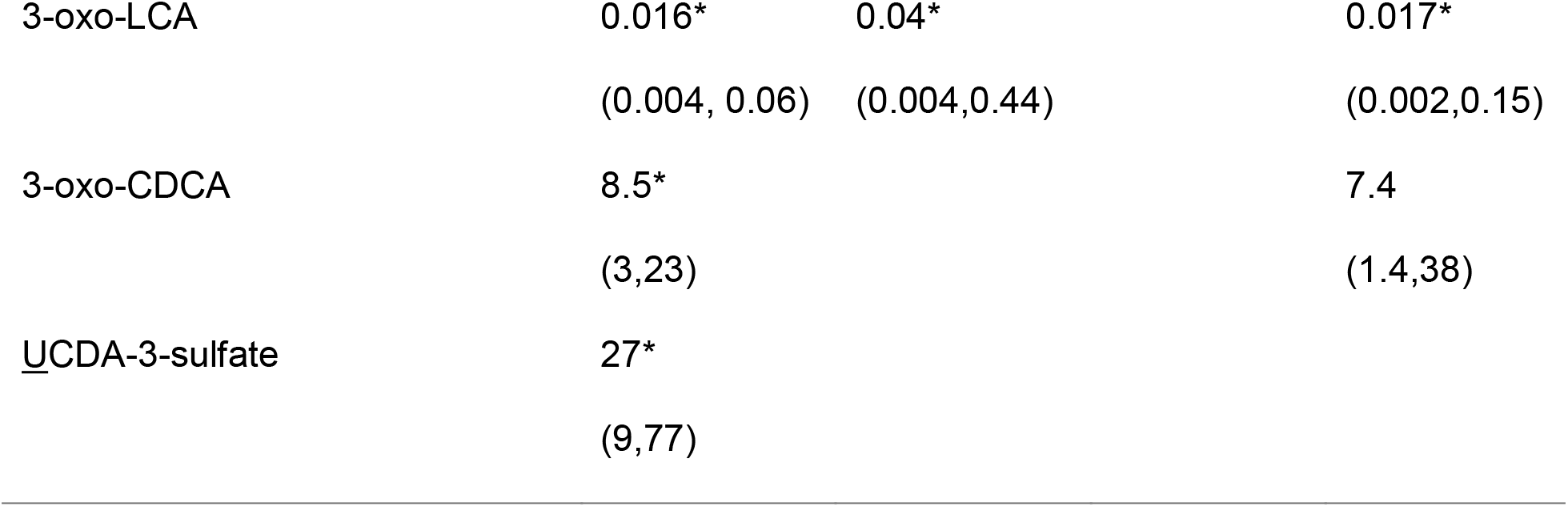
Estimated pre-FMT recipient/donor BA ratios (95% CI) in the four recipient groups. A ratio > 1 indicates ↑ recipient pre-FMT BA level compared to donor. A ratio of < 1 indicated ↓ recipient pre-FMT level compared to donor. Only the estimated ratios with an unadjusted p-value < 0.05 are listed. The * indicates those BA that also had a significant estimated ratios of post-FMT/pre-FMT ratios at the 1 week and/or 3 month time point, indicating that FMT had a significant

**Table 5.** Estimated donor – pre-FMT recipient BA differences (*pmol/g* wet weight stool or *µM*, 95% CI) in the four recipient groups. A positive difference indicates that the donor secondary unconjugated BA level was greater than the pre-FMT recipient level. The (*) indicates significant paired end differences between the pre-FMT and the 1 week and/or 3 month post-FMT (p-value < 0.05).

| Secondary unconjugated BA | rCDI – IBD | rCDI + IBD | UC – rCDI | CD – rCDI |
| --- | --- | --- | --- | --- |
| DCA | 6204* | 3749 |  |  |
|  | (4444,7694) | (685,6813) |  |  |
| LCA | 10596* | 7968 |  | 5621 |
|  | (7785,13408) | (3063,12873) |  | (996,10246) |
| iso-DCA | 1871* | 1314 |  | 715* |
|  | (1491,2251) | (656,1972) |  | (94,1335) |

### Shotgun DNA metagenomics data

Shotgun DNA metagenomics sequencing was performed on samples from 32 donor/recipient pairs (118 samples). After removing human sequences, an average of 21,513,616 sequences (range 220,028 - 59,164,588) was obtained for each sample. We initially screened the samples for extended-spectrum beta lactamase (ESBL) genes given the recent report of a fatality from transmission of an ESBL producing *E. coli* from donor to recipient [42]. OXA-137, an ESBL gene was detected in one rCDI no IBD recipient prior to the procedure and at one week post–FMT but not at three months post-FMT [43]. No ESBL genes were detected in the 32 donor stools sequenced.

Conversion of unconjugated primary BA to secondary BA (e.g. CA to DCA, and CDCA to LCA), is mediated by enzymes encoded by 8 genes in the BA-inducible (*bai*) operon (see Fig 4), which is carried by a small population of commensal bacteria, including *Clostridium scindens*, *Clostridium hylemonae*, *Clostridium hiranonis* [29]. It has been reported that fecal *baiCD* gene copy numbers is negatively correlated with CDI in humans [44], although concomitant presence of *C. scindens* and *C. difficile* has been reported in a metagenomics study [45]. The abundances of the *baiCD* gene, (coding the 5β-reductase), and the *baiE* gene, (which encodes the 7α-dehydroxylase), were estimated by translated searches of the shotgun metagenomics sequence data with DIAMOND [40] using the *C. scindens* protein sequence for the *baiE* and b*aiCD* genes at two threshold levels: 1.) 80% identity over 80% sequence and 2.) 50% identity over 80% sequence to calculate the counts per million (cpm). There were high correlations between the *baiE* cpm and *baiCD* cpm at both homology thresholds, consistent with both genes being in the same operon (see S17 Table). Lowering the threshold for identity from 80% to 50% markedly increased the *baiCD* gene cpm, but had only a modest effect on the *baiE* gene cpm. This suggested that a much larger number of bacterial genes, which may include genes not involved in the BA metabolism pathways, share protein sequence homology with the *baiCD* protein than the *baiE* encoded 7α-dehydroxylase.

Previous studies have reported a correlation between *baiCD* gene counts measured using degenerate PCR primers and rCDI [28, 43]. Using these degenerate primers, we observed multiple amplicons with a large range of sizes after analyzing the PCR reactions by gel electrophoresis, but only observed single amplicon bands using specific *C. scindens* and *C. hiranonis* primers. Consistent with previous studies, a higher proportion of rCDI ± IBD recipients lacked detectable *baiE* cpm compared to donors (see Fig 6).

**Fig 6.**
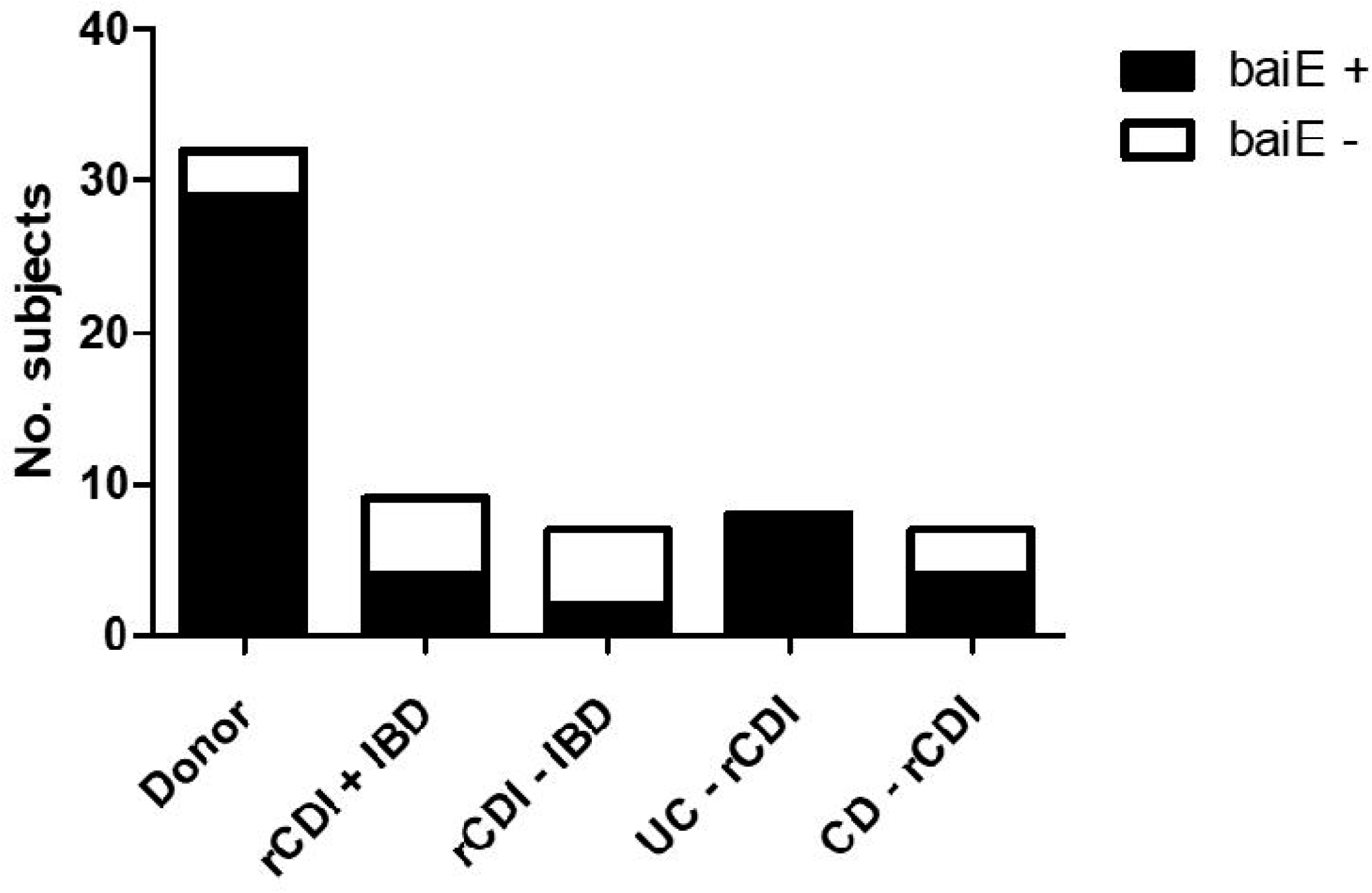
Detection of *baiE* gene (50% identity DIAMOND search of shotgun metagenomics data) in donors and the 4 recipient groups at baseline.

In contrast to the limited number of baiE 7α-dehydroxylase protein sequences in relatively few strains, there are hundreds of bacterial bile salt hydrolase (BSH) protein sequences distributed among 591 intestinal bacterial strains within 119 genera in the human microbiome [46]. Song *et al*. [46] categorized these sequences into 8 BSH phylotypes (BSH-T0 to BSH-T7). The custom DIAMOND search used in this study, included 31 BSH protein sequences from eight BSH phylotypes (S1 Table). The abundance of each BSH phlyotype was calculated as the sum of the individual BSH protein sequence cpms at 80% identity. As shown in Fig 7, the pattern of BSH phylotypes for the donors, UC-rCDI and CD-rCDI observed in this study was similar to the pattern described for the US population by Song *et al*. [46], with a relative prominence of the BSH-T6 phylotype. In contrast, the relative abundances of all BSH phylotypes were reduced in the rCDI – IBD and rCDI + IBD recipients prior to FMT.

**Fig 7.**
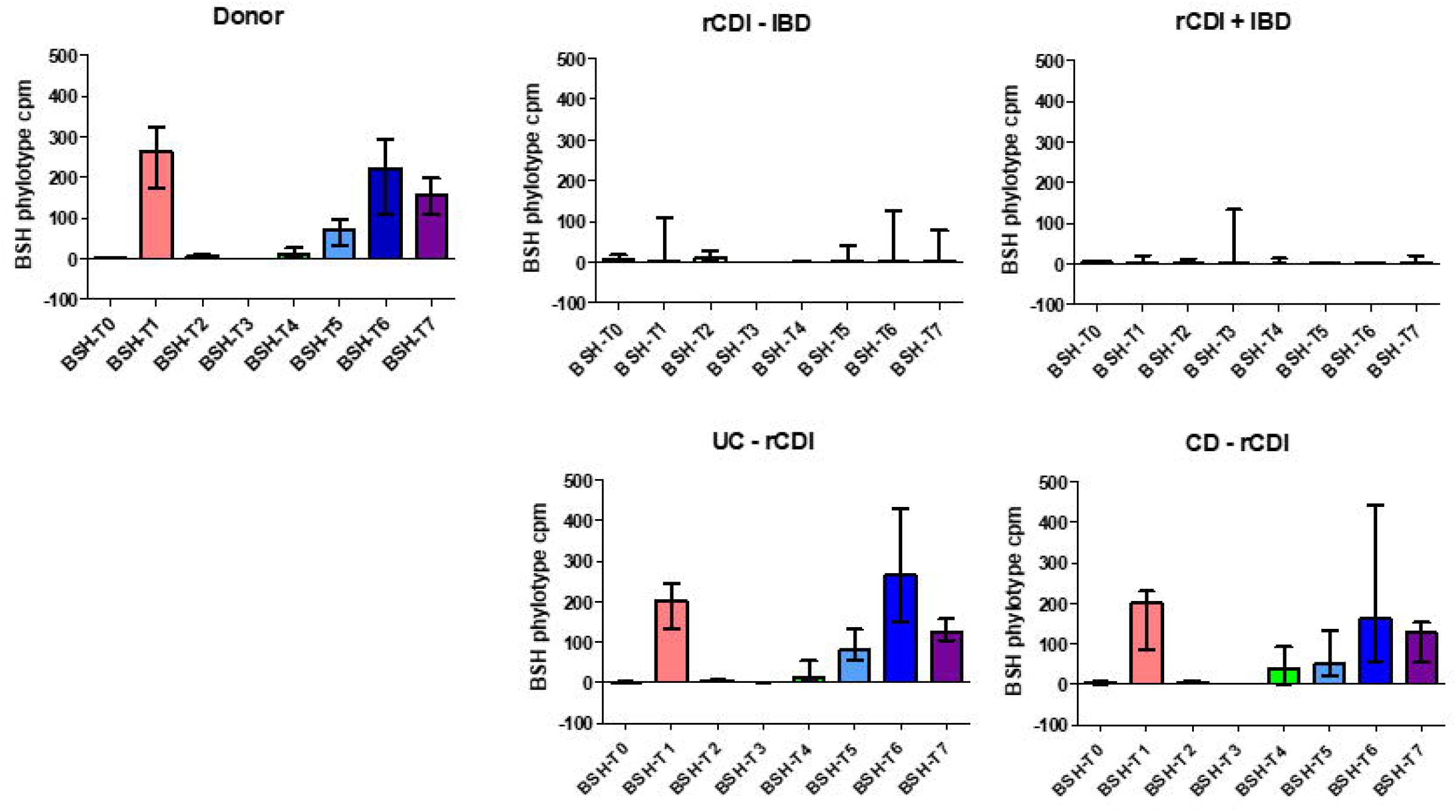
Median BSH phylotype abundances ± IQR (80% identity DIAMOND search of shotgun metagenomics data) in donors and 4 recipient groups at baseline.

The 3-α- and 3-β-hydroxysteroid dehydrogenases (HSDHs) from *R. gnavus* and *E. lenta*, metabolize LCA to 3-oxo-LCA and then to iso-LCA (see Fig 4), which exhibit *in vitro* activity on T-cell regulation [30–32]. Not surprisingly, the correlations were significant between the gene counts of HSDHs (80% identity) from the same taxa, however the correlation between the 3β-HSDH between *R. gnavus* and *E. lenta* was not significant (see S18 Table). No significant differences in the median *R. gnavus* 3α-HSDH cpm or 3β-HSDH cpm were observed between the donors and the pre-FMT levels of the four recipient groups, although decreased estimated pre-FMT recipient/donor ratios for Ruminococcus genus rRNA gene was observed in the rCDI - IBD and rCDI + IBD groups (see Table 3). A significant difference was observed for median *E. lenta* β-HSDH cpms between the donors and the Pre-FMT levels of the four recipient groups (*P* = 0.0117), with significant difference between the rCDI - IBD and the UC - rCDI groups (*P* = 0.0184), and between the rCDI + IBD and UC - rCDI groups (*P* = 0.0200). Similarly, a significant difference was also observed for median *E. lenta α-HSDH* cpms between the donors and the pre-FMT levels of the four recipient groups (*P* = 0.0065), with significant difference between the rCDI - IBD and the UC - rCDI groups (*P* = 0.0252), and between the rCDI + IBD and UC - rCDI groups (*P* = 0.0157).

Because *B. dorei* has been reported to convert iso-LCA to isoallo-LCA, a DIAMOND search was conducted for the putative *B. dorei* 5β-reductase, 5α-reductase and 3β-HSDH enzymes identified by BLAST searches reported by Li *et al.* [32]. All three of these protein sequences are significantly correlated with each other (see S19 Table). The median cpms for each of these candidate genes were significantly decreased in the rCDI + IBD group compared to donors (*P* = 0.0003, 0.0003, <.0001 for 3β-HSDH, 5β-reductase and 5α-reductase respectively) and the UC - rCDI groups (*P* = 0.0076, 0.0059, 0.0056 for 3β-HSDH, 5β-reductase and 5α-reductase respectively) and decreased in the rCDI - IBD groups compared to donors (*P* = 0.0002, 0.0007, <0.0001 for 3β-HSDH, 5β-reductase and 5α-reductase respectively) and the UC - rCDI groups (*P* = 0.0144, 0.0202, 0.0167 for 3β-HSDH, 5β-reductase and 5α-reductase respectively). This is consistent with the marked decrease in the estimated pre-FMT recipient donor ratios for the Bacteroides genus rRNA gene in the rCDI - IBD and rCDI + IBD groups (see Table 3).

In order to integrate the shotgun metagenomics data with the BA metabolite data, we measured the correlation between the abundances of the BA metabolizing genes (cpms) and fecal BA levels using linear mixed models, which clustered values from each FMT group, to calculate the estimated coefficient and 95% confidence interval (CI) (see Tables 6 and 7). Log transformation was applied if the normality assumption was not satisfied. As shown in Table 6, significant negative relationships between the *baiE* gene abundance (cpm) were observed for fecal chenodeoxycholic acid and cholic acid levels with negative estimated coefficients and p-values < 0.05. Significant positive relationships between the *baiE* gene abundance were observed for fecal secondary BA (DCA and LCA) levels with positive estimated coefficients and p-values < 0.05. Significant positive relationships between the *baiE* gene abundance were also observed for fecal secondary BA epimer, (iso-LCA acid and isoallo-LCA) and the secondary BA intermediate (3-oxo-LCA) levels. No significant correlation was observed between any of the four HSDHs and either 3-ox-LCA or iso-LCA levels. No significant relationships were observed between the gene counts for any of the three candidate *B. dorea* enzymes (5β-reductase, 5α-reductase and 3β-HSDH) and fecal isoallo-LCA levels, which is present in very low levels across donors and all four recipient groups. Taken together these results suggest that the *baiE* encoded 7α-hydroxylase step may be also rate limiting for the generation of the secondary BA epimers implicated in immunomodulation.

**Table 6.**
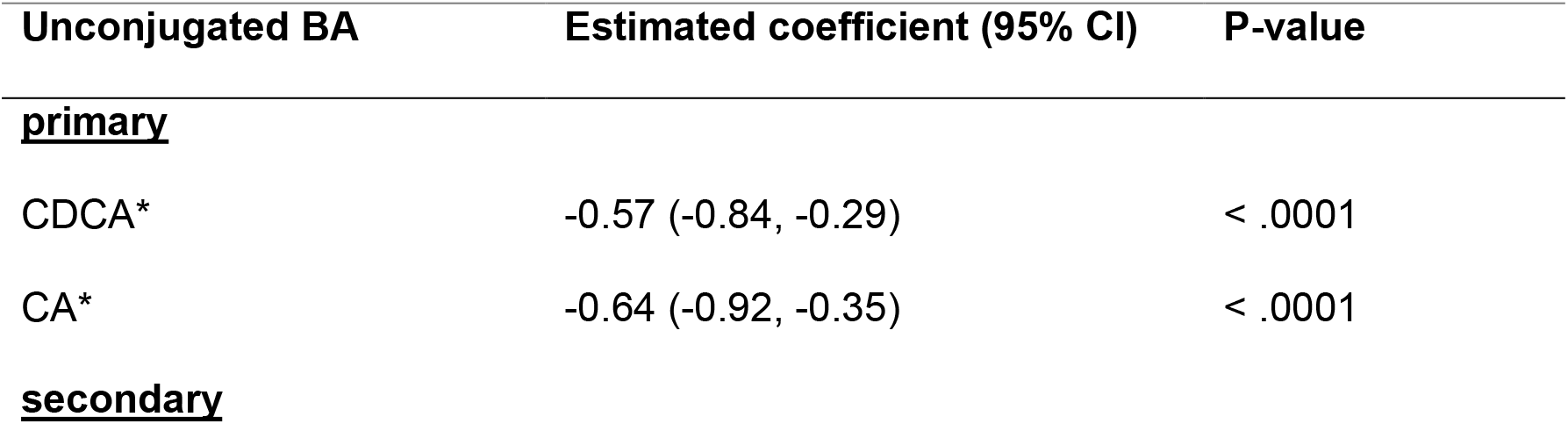
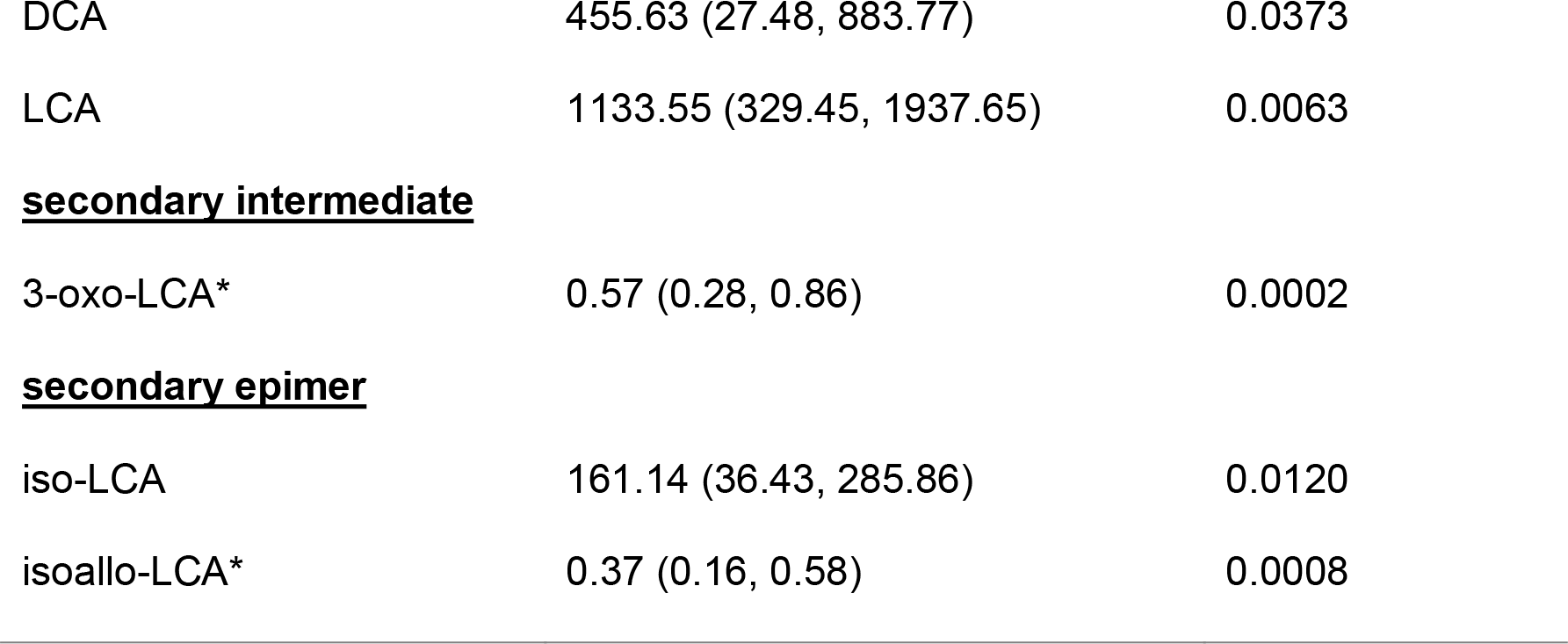
Estimated coefficient and 95% CI for selected unconjugated BA for *baiE* cpm (50% identity threshold) in linear mixed models treating FMT group as clustering effect. * indicates that the value was log-transformed, since the normality assumption was not satisfied.

| Unconjugated BA | Estimated coefficient (95% CI) | P-value |
| --- | --- | --- |
| **<u>primary</u>** |  |  |
| CDCA* | -0.57 (-0.84, -0.29) | < .0001 |
| CA* | -0.64 (-0.92, -0.35) | < .0001 |
| **<u>secondary</u>** |  |  |
| DCA | 455.63 (27.48, 883.77) | 0.0373 |
| LCA | 1133.55 (329.45, 1937.65) | 0.0063 |
| **secondary intermediate** |  |  |
| 3-oxo-LCA* | 0.57 (0.28, 0.86) | 0.0002 |
| **secondary epimer** |  |  |
| iso-LCA | 161.14 (36.43, 285.86) | 0.0120 |
| isoallo-LCA* | 0.37 (0.16, 0.58) | 0.0008 |

**Table 7.**
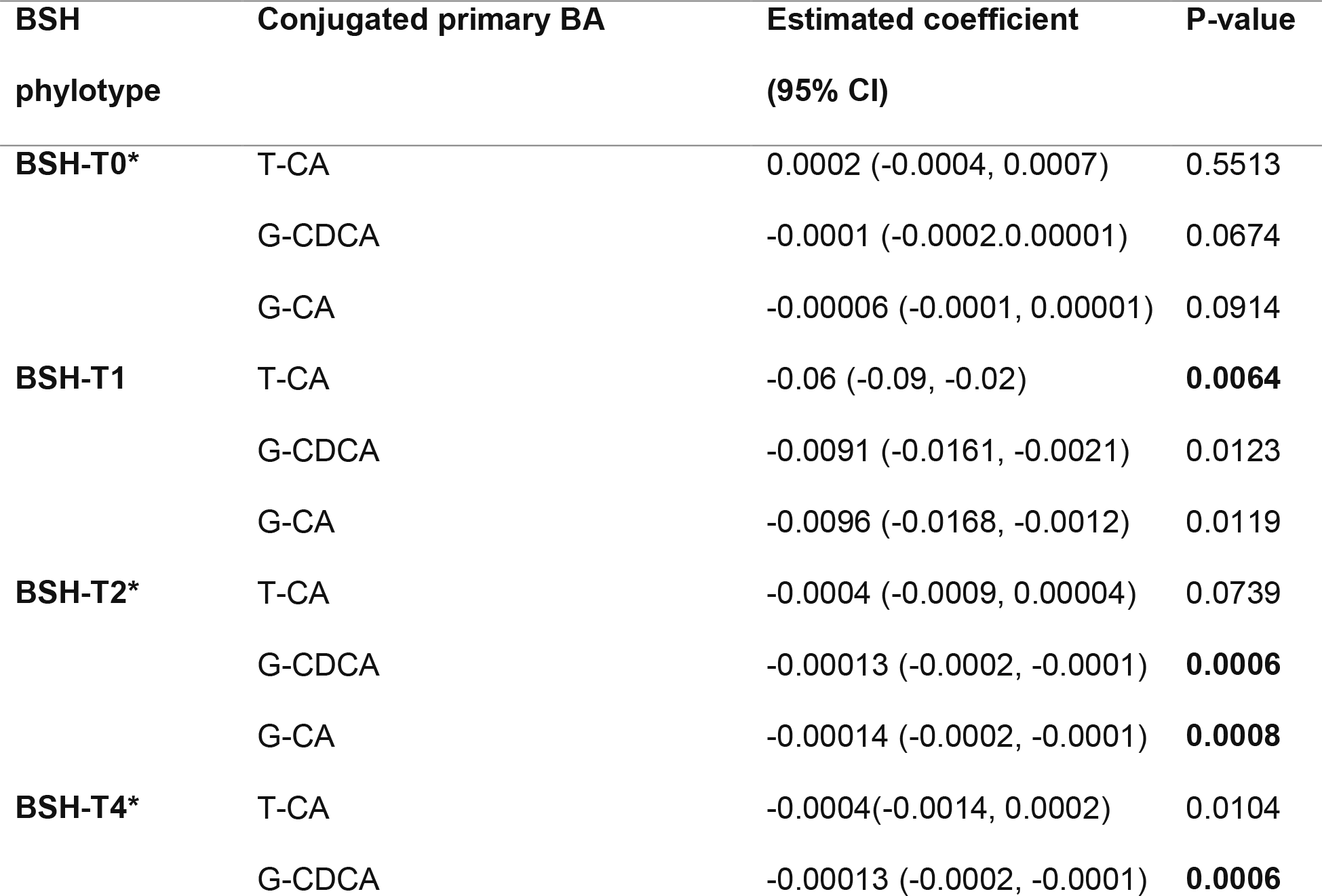
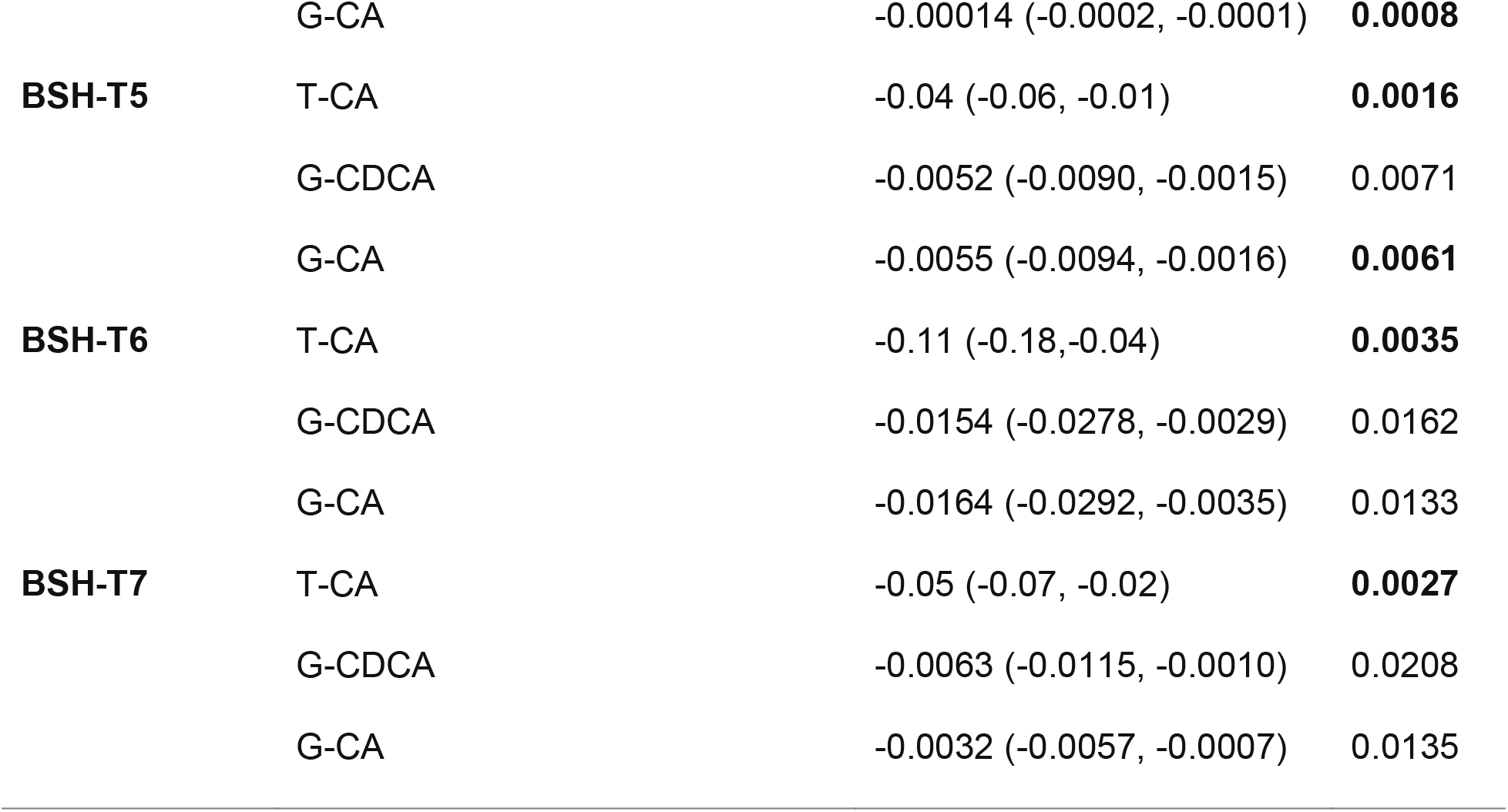
Estimated coefficient and 95% CI for each conjugated primary BA for 7 of the 8 BSH phylotype cpm (80 % identity threshold) in linear mixed models treating FMT group as clustering effect. * indicates that value was log-transformed.

| **BSH**<br>**phylotype** | **Conjugated primary BA** | **Estimated coefficient**<br>**(95% CI)** | **P-value** |
| --- | --- | --- | --- |
| **BSH-T0\*** | T-CA | 0.0002 (-0.0004, 0.0007) | 0.5513 |
|  | G-CDCA | -0.0001 (-0.0002, 0.00001) | 0.0674 |
|  | G-CA | -0.00006 (-0.0001, 0.00001) | 0.0914 |
| **BSH-T1** | T-CA | -0.06 (-0.09, -0.02) | **0.0064** |
|  | G-CDCA | -0.0091 (-0.0161, -0.0021) | 0.0123 |
|  | G-CA | -0.0096 (-0.0168, -0.0012) | 0.0119 |
| **BSH-T2\*** | T-CA | -0.0004 (-0.0009, 0.00004) | 0.0739 |
|  | G-CDCA | -0.00013 (-0.0002, -0.0001) | **0.0006** |
|  | G-CA | -0.00014 (-0.0002, -0.0001) | **0.0008** |
| **BSH-T4\*** | T-CA | -0.0004(-0.0014, 0.0002) | 0.0104 |
|  | G-CDCA | -0.00013 (-0.0002, -0.0001) | **0.0006** |
|  | G-CA | -0.00014 (-0.0002, -0.0001) | **0.0008** |
| **BSH-T5** | T-CA | -0.04 (-0.06, -0.01) | **0.0016** |
|  | G-CDCA | -0.0052 (-0.0090, -0.0015) | 0.0071 |
|  | G-CA | -0.0055 (-0.0094, -0.0016) | **0.0061** |
| **BSH-T6** | T-CA | -0.11 (-0.18, -0.04) | **0.0035** |
|  | G-CDCA | -0.0154 (-0.0278, -0.0029) | 0.0162 |
|  | G-CA | -0.0164 (-0.0292, -0.0035) | 0.0133 |
| **BSH-T7** | T-CA | -0.05 (-0.07, -0.02) | **0.0027** |
|  | G-CDCA | -0.0063 (-0.0115, -0.0010) | 0.0208 |
|  | G-CA | -0.0032 (-0.0057, -0.0007) | 0.0135 |

Because there were multiple zero counts for BSH-T3 phylotype abundance, this phylotype was excluded from our analysis. BSH-T3 is composed of lactobacillus BSH genes. To correct for multiple comparisons between the remaining 7 BSH phylotype relative abundances (cpm) and the fecal levels of the three conjugated primary BAs, T-CA, G-CDCA and G-CA we applied the Bonferroni correction to set the threshold p-value as 0.05/7 = 0.007. Based on this threshold, we observed significantly negative relationships between BSH-T1, BSH-T5 BSH-T6 and BSH-T7 phylotype relative abundances (cpm) and fecal T-CA levels. We observed significantly negative relationships between BSH-T2 and BSH-T4 phylotype abundances (cpm) and fecal G-CDCA and G-CA. These results may be consistent with varying substrate specificities of bacterial BSH enzymes for the taurine and glycine BA conjugates [47, 48].

## Discussion

We report the results of integrating 16S RNA gene sequencing, shotgun metagenomics sequencing and quantitative BA metabolomic analysis in human stools collected during two colonoscopic single donor fecal microbial FMT clinical trials conducted at SBUMC (2013-2020). Our quantitative analysis of primary and secondary BA metabolites in human stool is novel in that secondary BA epimers, which have recently reported to exhibit immunoregulatory activity *in vitro*, were also measured in this study.

Decreases in secondary BA have been observed in rCDI and more recently in IBD [28, 49]. In this study, we observed a general reduction in the levels of secondary BA including the immunomodulatory derivatives of LCA in the rCDI – IBD, the rCDI + IBD and CD - rCDI groups but not in the UC-rCDI group at baseline prior to transplant. Post FMT, the levels of secondary BA were restored in the rCDI – IBD group and to various extents in the rCDI + IBD and CD – rCDI groups.

Parallel estimation of the abundance of secondary BA pathway related protein/gene sequences by polymerase chain reaction assays or shotgun metagenomics sequence analysis and correlating these abundances with metabolite levels could potentially help us predict BA metabolic potential in the human gut [50]. Our approach has been to search the shotgun metagenomics data based on protein sequences of relevant BA-metabolizing enzymes. Because of the knowledge base accumulated on the bacterial metabolism of primary to secondary BA we focused initially on the genes in the *bai* operon, which is present in only a select number of *Clostridium* species. Based on our analysis supplemented by qualitative PCR assays, we have identified the *baiE* gene cpm (measured at 50% identity over 80% sequence) as the biomarker associated with a rate-limiting step in the conversion of unconjugated primary BAs to secondary BAs. It is not clear why a significant correlation between the abundances of the other genes, such as *baiCD*, in the bai operon and secondary BA metabolite levels were detected in this study. Based on gel electrophoresis analysis of products of the *baiCD* gene PCR assay, we suspect there may be significant homology between that gene sequence and other genes which are unrelated to the BA transformation pathway.

Primary BA must be deconjugated prior to transformation to secondary BA. Consequently loss of BSH activity may also affect secondary BA levels. In contrast to the limited number of taxa involved in the initial conversion of unconjugated primary BA to secondary BA, multiple BSH gene sequences from multiple taxa are implicated in deconjugation of conjugated primary BA. Furthermore some of the homologous gene sequences are pseudogenes that may not contribute to BSH activity [46]. Our approach to estimating BSH activity based on BSH gene abundances was to calculate the sum of selected BSH phylotype gene sequence abundances at 80% homology for each of 8 BSH phylotype described by Song *et al* [46]. This approach yielded donor profiles of BSH phylotype abundances that matched those reported for the US population [46]. BSH phylotype abundances were markedly reduced particularly in the rCDI + IBD recipients prior to FMT compared to healthy donors. Furthermore reduced BSH phylotype abundances were significantly correlated with increased levels of conjugated primary BA levels. These results suggest that IBD patients with rCDI have reduced secondary BA levels due to loss of both 7-α hydroxylase and BSH activities.

*B. dorei* 5β-reductase, 5α-reductase and 3β-HSDH gene abundances are reduced in rCDI ± IBD recipients compared to healthy donors. It has been reported that centenarians have microbiomes that are enriched for *B. dorei* and additional bacterial strain isolates capable of producing isoallo-LCA acid [51]. However it should also be pointed out that LCA levels are also reduced in the rCDI ±IBD recipients along with reduced abundance of the *baiE* gene encoding the 7-alpha hydroxylase step. The contribution of alterations in the abundances of the 5β-reductase, 5α-reductase and 3β-HSDH genes to the production of secondary BA epimers may be reduced simply because more upstream processes drastically reduce levels of secondary BA substrates, such as LCA and DCA.

A major limitation of this study is the small number of subjects in each of the three IBD recipient groups and that these studies were single arm in design. Unfortunately, the COVID pandemic has essentially forced a halt to these clinical trials. Nevertheless, the samples and multi-omic datasets generated from these samples will continue to provide an important resource for characterizing the effect of FMT on microbial function as further progress is made in identifying bacterial strains and enzymes involved in the biotransformation of BAs and other metabolites.

## Conclusion

Secondary BA metabolite levels, including epimers implicated in immune regulation, are reduced in rCDI ± IBD and CD – rCDI recipients compared to donors. Single donor colonoscopic FMT restored many of those levels in these recipients. Integration of shotgun metagenomics and quantitative targeted BA metabolomics analyses indicate that reduction of *baiE* gene abundance is correlated with reduction of secondary BA metabolite levels. In addition BSH phylotype gene abundance is also reduced in particularly rCDI + IBD recipients and is negatively correlated with conjugated primary BA levels.

## Data Availability

All data produced in the present work are contained in the manuscript.

## Acknowledgements

We want to thank the recipients and donors for participating in this study. We acknowledge the contributions of Drs. Ramona Rajapakse, Michael J Clores, Lionel D’Souza, Alexandra Guillaume in the Division of Gastroenterology and Hepatology, for performing some of the FMT procedures. We acknowledge the biostatistical consultation provided by Junying Wang and the Biostatistics and Data Science Shared Resource at the Stony Brook Cancer Center, Stony Brook University. We thank Dr. Daniel Littman for his helpful comments on the manuscript.

## Supporting information

**S1 text. Institutional review board approved protocol for ClinicalTrials.gov ID:NCT03268213.**

**S2 text. Institutional review board approved protocol for ClinicalTrials.gov ID NCT03267238.**

**S1 Table. Transparent Reporting of Evaluations with Nonrandomized Designs (TREND) checklist.**

**S2 Table. Chromatographic and quantitation strategy for the BA and internal standards (ISTD) measured.**

**S3 Table. Bile acid metabolizing enzyme genes used in DIAMOND custom search.**

**S4 Table. Individual characteristics of the rCDI – IBD recipients.**

**S5 Table. Individual characteristics of the rCDI + IBD recipients.**

**S6 Table. Individual characteristics of the UC - rCDI recipients.**

**S7 Table. Individual characteristics of the CD - rCDI recipients.**

**S8 Table. Significant (FDR < 0.05) estimated pre-FMT recipient/donor ratios (95% CI) of the relative abundances of Actinobacteria genus level OTUs for each of the four recipient groups.**

**S9 Table. Significant (FDR < 0.05) estimated pre-FMT recipient/donor ratios (95% CI) of the relative abundances of Bacteroidetes genus level OTUs for each of the four recipient groups.**

**S10 Table. Significant (FDR < 0.05) estimated pre-FMT recipient/donor ratios (95% CI) of the relative abundances of Ruminococcaceae (Clostridium Group IV) genus level OTUs for each of the four recipient groups.**

**S11 Table. Significant (FDR < 0.05) estimated pre-FMT recipient/donor ratios (95% CI) of the relative abundances of Lachnospiraceae (Clostridium Group XIVa) genus level OTUs for each of the four recipient groups.**

**S12 Table. Significant (FDR < 0.05) estimated pre-FMT recipient/donor ratios (95% CI) of the relative abundances of Bacilli genus level OTUs for each of the four recipient groups.**

**S13 Table. Significant (FDR < 0.05) estimated pre-FMT recipient/donor ratios (95% CI) of the relative abundances of Proteobacteria genus level OTUs for each of the four recipient groups.**

**S14 Table. Significant (FDR < 0.05) estimated pre-FMT recipient/donor ratios (95% CI) of the relative abundances of genus level OTUs in “Other Taxa” for each of the four recipient groups.**

**S15 Table. Median Pre-FMT BA levels (pmol/g wet weight of stool or µM) ± IQR in donor and four recipient groups.**

**S16 Table. Post-hoc Dunns test P-values with Benjamini–Hochberg adjustments, for 19 pre-FMT median BA metabolite levels in the four recipient groups (see S15 Table).**

**S17 Table. Estimated coefficient (95% CI) with baiE cpm as covariate and baiCD cpm as outcome in linear mixed model treating FMT group as clustering effect.**

**S18 Table. Estimated coefficient (95% CI) with R. gnavus 3α-HSDH/E. lenta 3α-HSDH/E. lenta 3β-HSDH cpm as covariate and R. gnavus 3β-HSDH/E. lenta 3α-HSDH cpm at 80% identity as outcome in linear mixed model treating FMT group as clustering effect**

**S19 Table. Correlation between *B. dorei* 3β-reductase, 5β-reductase and 3β-HSDH.**

