## Supplemental Text S1 for "Integrated metagenomic and bile acid metabolomic analysis of human fecal microbiota transplantation for recurrent Clostridioides difficile and/or inflammatory bowel diseases"

### Protocol

**TITLE:** Fecal microbial transplantation in patients with medication-refractory *Clostridium difficile* and/or Ulcerative colitis or indeterminate colitis.

**ClinicalTrials.gov ID:NCT03268213, 479696, IND 15642**

**INVESTIGATORS:**

| | |
|---|---|
| Anupama Chawla M.D. (Principal Investigator) | Professor of Pediatric Gastroenterology |
| Ellen Li M.D., PhD (co-PI) | Professor of Medicine/Gastroenterology |
| Jeffrey Morganstern M.D. | Associate Professor of Pediatric Gastroenterology |
| Grace Gathungu M.D. | Assistant Professor of Pediatric Gastroenterology |
| Lesley Small-Harary M.D. | Assistant Professor of Pediatric Gastroenterology |
| Michelle Tobin M.D. | Assistant Professor of Pediatric Gastroenterology |
| Jonathan Buscaglia M.D. | Associate Professor of Medicine/Gastroenterology |
| Juan Carlos Bucobo, M.D. | Assistant Professor of Medicine/Gastroenterology |
| Lionel D'Souza, M.D. | Assistant Professor of Medicine/Gastroenterology |
| Alexandra Guillaume, M.D. | Assistant Professor of Medicine/Gastroenterology |
| Bradley Morganstern, M.D. | Assistant Professor of Medicine/Gastroenterology |
| Farah Monzur, M.D. | Assistant Professor of Medicine/Gastroenterology |

#### A. SPECIFIC AIMS:

The following hypothesis will be tested in this study:

1. Fecal microbiota transplantation is a safe, tolerable and efficacious procedure.
2. The fecal microbial diversity, composition and function in stool recipients after fecal transplantation will change to a similar microbial diversity, composition and functionality as found in donor stool.

Primary objectives:

1. To determine the short term safety, tolerability and efficacy of fecal microbiota transplantation up to 12 weeks post-transplant in patients with
    a. Recurrent or refractory *Clostridium difficile* infection (CDI) not responsive to standard therapy in subjects without inflammatory bowel diseases
    b. Recurrent or refractory CDI not responsive to standard therapy in subjects with inflammatory bowel disease.
    c. Ulcerative colitis or indeterminate colitis without CDI who have required therapy beyond 5-ASA or mesalamine therapy alone

Secondary objectives:

1. To determine the long term safety, tolerability and efficacy of fecal microbiota transplantation (FMT) up to one year post transplant in patients with
    a. Recurrent or refractory CDI not responsive to standard therapy in subjects without inflammatory bowel diseases
    b. Recurrent or refractory CDI not responsive to standard therapy in subjects with inflammatory bowel disease.
    c. Ulcerative colitis or indeterminate colitis without CDI who have required therapy beyond 5-ASA or mesalamine therapy alone

2. To compare microbial diversity, composition and function in healthy donor stools compared to pre-FMT recipient stools collected from patients (recipients) with
    a. Recurrent or refractory CDI not responsive to standard therapy in subjects without inflammatory bowel diseases
    b. Recurrent or refractory CDI not responsive to standard therapy in subjects with inflammatory bowel disease.
    c. Ulcerative colitis or indeterminate colitis without CDI who have required therapy beyond 5-ASA or mesalamine therapy alone

3. To compare microbial diversity, composition and function in healthy donor stools and pre-FMT recipient stools with 1 week post transplant recipient stool samples collected from patients (recipients) with
    a. Recurrent or refractory CDI not responsive to standard therapy in subjects without inflammatory bowel diseases
    b. Recurrent or refractory CDI not responsive to standard therapy in subjects with inflammatory bowel disease.
    c. Ulcerative colitis or indeterminate colitis without CDI who have required therapy beyond 5-ASA or mesalamine therapy alone

4. To compare microbial diversity, composition and function in healthy donor stools and pre-FMT recipient stools with 12 week post transplant recipient stool samples collected from patients (recipients) with
    a. Recurrent or refractory CDI not responsive to standard therapy in subjects without inflammatory bowel diseases
    b. Recurrent or refractory CDI not responsive to standard therapy in subjects with inflammatory bowel disease.
    c. Ulcerative colitis or indeterminate colitis without CDI who have required therapy beyond 5-ASA or mesalamine therapy alone

5. Stool calprotectin levels will be measured in the recipient at baseline pre-FMT, 1 week and 12 weeks post FMT to determine if FMT causes a statistically significant change.

**B. BACKGROUND AND SIGNIFICANCE**

*Clostridium difficile* is the leading cause of antibiotic-associated diarrhea with increasing infection rates and economic burden in developed countries. [1-4] According to the United States Healthcare Cost and Utilization Project Kids' Inpatient Database (HCUP-KID), there has been an increase in pediatric *Clostridium difficile* from a rate of 7.24/10000 hospitalizations in 1997 to 12.80/10000 hospitalizations in 2006.[5] As the *C. difficile* epidemic worsens, the numbers of failed treatments and patients who experience relapses or recurrences also continues to grow. Patients being treated with usual first-line treatments such as metronidazole and vancomycin are encountering "hypervirulent" and resistant strains of *C. difficile* such as North American Pulsed Field type 1 (NAP1), restriction-endonuclease analysis (REA) type BI, or polymerase-chain reaction ribotype 027 (referred to collectively as the NAP1/B1/027 strain).[6] The NAP1/B1/027 strain of *C. difficile* has been deemed "hypervirulent" for its ability to produce binary toxin *C. difficile* 126 adenosine diphosphate-ribosyltransferase not typically found in other strains of *C. difficile*. This, in combination with its ability to produce excessive quantities of enteroxins A and B, compared with other strains of *C. difficile*, makes it hypervirulent[7] Even the newer agent, fidaxomicin, which was approved by the FDA in 2011, has a similar effectiveness to vancomycin with respect to the clinical resolution of acute diarrheal disease due to *C. difficile*.[6] The problem is that after completing antibiotic treatment 20-35% of CDI patients experience a recurrence of CDI. Vancomycin tapers or fidaxomicin may be associated with a lower rate of recurrence. Once recurrent CDI occurs, 45-65% of patients will continue to experience recurrent infections over several years and have a higher mortality.[7-11]

Fecal microbial transplantation (FMT) of healthy donor stool into the patient's gut has emerged as one of the most effective treatments of recurrent CDI.[12-14] Patients with inflammatory bowel diseases (IBD), particularly ulcerative colitis (UC) and indeterminate colitis (IC) but also Crohn's disease (CD), are at increased risk of developing CDI.[15-20] FMT is also highly effective in treatment of recurrent CDI with IBD patients, although the effectiveness may be reduced compared to patients with recurrent CDI without IBD.[21-23] While some patients with recurrent CDI and IBD appear to benefit overall from the FMT, there have been reports of exacerbation of IBD after eradication of CDI, although it remains to be determined whether FMT contributed to the exacerbation.[24,25]

Thus far studies on the effectiveness of FMT in the treatment of IBD without CDI, have utilized different protocols and have yielded mixed results.[26-38] Performing FMT in patients with IBD without CDI appears to be safe. As noted above, a review of FMT trials conducted on IBD patients with and without CDI noted that post-FMT exacerbation of IBD patients without recurrent CDI appeared to occur less often than in IBD patients with recurrent CDI.[25]

Colonoscopic instillation into the right side of the colon of ~50 g stool is associated with the highest rates of preventing further CDI (~90%). The effectiveness of the oral capsule route, may be related to the total amount of stool ingested and ranges from 68 to greater than 90%.[39-41] In a very recent randomized trial of oral capsule vs. colonic instillation of 150g of stool, ingestion of oral capsules was not inferior to colonoscopic administration (96% in both arms).[41]

FMT is believed to restore the normal microbiological balance in the recipient primarily by transferring microbial components of the donor stool, although there is some evidence that non-microbial molecular components, such as short-chain fatty acids and bile acids may also play a role.[42,43] The normal microbiological balance may be perturbed in patients with C. difficile infection and or IBD.[44]

The overall goal of this study protocol registered as ClinicalTrials.gov ID:NCT03268213, 479696, is to better define the microbial and non-microbial components in donor stool that contribute to post-FMT changes in the microbiome in the following three groups of recipients:

a. Recurrent or refractory CDI not responding to standard medical treatment without IBD,
b. Recurrent or refractory CDI not responding to standard medical therapy with IBD
c. Ulcerative colitis or indeterminate colitis without CDI who have required therapy beyond 5-ASA or mesalamine therapy alone

#### D. RESEARCH DESIGN AND METHODS

##### 1. Rationale/overview:

It is important to emphasize that this study is not designed to investigate clinical efficacy. FMT clinical efficacy for preventing further recurrence of CDI is now well established in the literature for recipients with recurrent or refractory CDI not responding to standard medical treatment, with and without IBD. However it remains to be determined what the key microbial or nonmicrobial components of the donor stool contribute to clinical efficacy. Consequently this study is designed to:

a. Measure baseline microbial imbalances in these three groups of recipients (*i.* recurrent or refractory CDI not responding to standard medical treatment without IBD, *ii.* recurrent or refractory CDI not responding to standard medical therapy with IBD *iii.* UC and IC without CDI.) compared to healthy donors.
b. Measure whether FMT will alter recipient microbial imbalances one week and 12 weeks after the procedure.
c. Measure short (12 weeks) and long term (one year) safety, tolerability and efficacy of FMT in these three groups of recipients.

##### 2. FDA statement on requirement for IND for FMT.

The March 2016 Enforcement Policy Regarding Investigational New Drug Requirements for Use of Fecal Microbiota for Transplantation to Treat Clostridium difficile Infection Not Responsive to Standard Therapies Draft Guidance for Industry[45] states: “We, FDA or Agency, are informing members of the medical and scientific community and other interested persons that we intend to exercise enforcement discretion under limited conditions, regarding the investigational new drug (IND) requirements for the use of fecal microbiota for transplantation (FMT) to treat Clostridium difficile (C. difficile) infection not responding to standard therapies. FDA intends to exercise this discretion, provided that: 1) the licensed health care provider treating the patient obtains adequate consent from the patient or his or her legally authorized representative for the use of FMT products. The consent should include, at a minimum, a statement that the use of FMT products to treat C. difficile is investigational and a discussion of its reasonably foreseeable risks; 2) the FMT product is not obtained from a stool bank; and 3) the stool donor and stool are qualified by screening and testing performed under the direction of the licensed health care provider for the purpose of providing the FMT product for treatment of the patient. A stool bank is defined, for the purpose of this guidance, as an establishment that collects, prepares, and stores FMT product for distribution to other establishments, health care providers, or other entities for use in patient therapy or clinical research. An establishment that collects or prepares FMT products solely under the direction of licensed health care providers for the purpose of treating their patients (e.g., a hospital laboratory) is not considered to be a stool bank under this guidance. FDA does not intend to extend enforcement discretion for the IND requirements applicable to stool banks distributing FMT products. Such distributions are subject to the requirements that a sponsor, typically the stool bank, have an IND in effect before distributing the FMT product to investigators for administration to subjects in accordance with the investigational plan under the Public Health Service (PHS) Act (42 U.S.C. 262(a)(3)) and 21 CFR Part 312. However, as described in this guidance, an IND sponsor may request a waiver of certain IND regulations applicable to investigators for those licensed health care providers receiving FMT product to treat patients with C. difficile infection not responsive to standard therapies. (See 21 CFR 312.10). FDA has developed this policy to assure that patients with C. difficile infection not responding to standard therapies may have access to this treatment, while addressing and controlling the risks that centralized manufacturing in stool banks presents to subjects. FDA intends for this to be an interim policy, while the Agency develops a comprehensive approach for the study and use of FMT products under IND." At this time, since administration of FMT products from stool bank(s) has not been approved by SBUH regulatory committees in the Endoscopy suite in Stony Brook University Hospital, administration of FMT products from stool banks cannot be offered to recipients with recurrent CDI.

An IND (IND 15642) was obtained from the FDA for performing FMT on UC and IC recipients without recurrent or refractory CDI not responsive to standard medical treatment.

**3. Research Site(s):**

The following research sites are all located within Stony Brook University Medical Center (SBUMC).

a. *Patient and donor recruitment and screening*. This will take place at Stony Brook University Medical Center (SBUMC) in both the hospital and outpatient Pediatric and Adult Gastroenterology Clinic setting. The Pediatric Gastroenterology Clinic is located at 4 Technology Drive, Suites 250 and 270, across the street from the Adult Gastroenterology Clinic, which is located at 3 Technology Drive, Suite 300.
b. *Preparation of donor stool for FMT.* The donor stool will be prepared for colonoscopic instillation following standard operating procedure, as described below, in a biosafety cabinet located within Dr. Li's laboratory within the Department of Medicine/Division of Gastroenterology research space located in in the Health Sciences Center Building (room HSC T17-080). The research laboratory in the HSC Building is connected by a bridge to Stony Brook University Hospital, allowing for rapid transport of the stool instillate to the Endoscopy unit on the 14th floor.
c. *Colonoscopic instillation of donor stool*. Colonoscopic administration of the FMT will be performed in the Stony Brook University Endoscopy Unit located
d. *Processing of stool samples for microbiome analysis*. The research stools samples obtained from the donor (at the time of transplant), and the recipient (pre-FMT, 1 week post-FMT, and 12 weeks post-FMT) will be processed in Dr. Li's laboratory within the Department of Medicine/Division of Gastroenterology research space

**4. Participants**

Each FMT will involve two participants, the **recipient** of the FMT, and a healthy known **donor** selected by the recipient or recipient's parents (pediatric recipient age <18). The inclusion and exclusion criteria for recipients with CDI with or without IBD are identical see **D4.a**). The inclusion and exclusion of the UC and IC only without recurrent CDI recipients are described separately under **D4.b**. The inclusion and exclusion criteria for the healthy donors are identical for all three recipient groups under **D4.c**.

**a. <u>Inclusion and exclusion criteria for recipients with recurrent or refractory CDI with or without IBD not responsive to standard medication:</u>**

*Inclusion*: (see Appendix A)

*i.* Recipient age ≥ 7 years of age.[46]
*ii.* Recurrent of refractory CDI with or without IBD not responding to standard medication therapy as one of the following criteria:
- The recipient has had at least three episodes or two recurrences of mild to moderate CDI (diarrheal stool ≤ 6 a day) diagnosed via positive polymerase chain reaction (PCR) or enzyme immunoabsorbant assay (EIA) for toxin after two courses of standard antibiotic treatment (including at least one course of either vancomycin or fidaxomicin).
- The recipient has had at least two episodes of severe CDI (diarrhea stool > 6 a day) within 6 months resulting in hospitalization and associated with significant morbidity
- The recipient has had moderate CDI (3-6 diarrheal stool not responding to successive standard therapy (e.g. metronidazole, vancomycin and/or fidaxomicin) lasting at least 28 days.

- The recipient has severe and/or fulminant CDI with no response to standard therapy after 48 h.

iii. A negative pregnancy test on day of FMT (female of childbearing age)
iv. Recipient agrees to share medical records for FMT consultation at SBUMC

*Exclusion* (see Appendix A):

i. Recipient age < 7 years of age.
ii. Scheduled for abdominal surgery within the next 12 weeks
iii. Pregnancy
iv. Grade 4 anemia (Hemoglobin < 6 g/dL)
v. Grade 1 neutropenia (Absolute Neutrophil Count <1500)
vi. Known diagnosis of Graft vs. host disease
vii. Major abdominal surgery within the past 3 months
viii. Administration of any investigational drug within the past 2 months
ix. Use of a TNF-α antagonist within 2 weeks of the proposed date of transplantation
x. Bacteremia within past 4 weeks (28 days)
xi. Recipients > 18 years of age unable or unwilling to give informed consent. The consent form will outline that although fecal microbiota transplantation appears safe based on past studies, a theoretical risk of transmission of an unrecognized infectious agent or substance exists and could result in an unexpected disease. For recipients with CDI and IBD, the consent form will outline that post FMT exacerbation of IBD has been reported in these patients.

**b. Inclusion and exclusion criteria for recipients with Ulcerative colitis (UC) or indeterminate colitis (IC) without CDI who have required therapy beyond 5-ASA or mesalamine therapy alone**

*Inclusion*: (see Appendix B)

i. Recipient age ≥ 7 years of age.
ii. Recipients diagnosed with UC or IC by primary gastroenterologist as documented by colonoscopy, pathology and imaging findings based on review of medical records.
iii. Recipients who have required therapy beyond 5-ASA or mesalamine therapy alone
iv. Recipient agrees to share medical records for FMT consultation at SBUMC

*Exclusion:* (see Appendix B)

i. Recipient age < 7 years of age.
ii. Scheduled for abdominal surgery within the next 12 weeks
iii. Pregnancy
iv. Grade 4 anemia (Hemoglobin < 6 g/dL)
v. Grade 1 neutropenia (Absolute Neutrophil Count <1500)
vi. Known diagnosis of Graft vs. host disease
vii. Major abdominal surgery within the past 3 months
viii. Administration of any investigational drug within the past 2 months
ix. Use of a TNF-α antagonist within 2 weeks of the proposed date of transplantation
x. Bacteremia within past 4 weeks (28 days)
xi. Any previous FMT

**c. Inclusion and exclusion criteria for healthy donors for all three groups of recipients (CDI with and without IBD and UC and IC only without CDI).**

*Inclusion* (see Appendix C):

i. Spouses, parents, family members, friends, or associates of the stool recipient who consent to completing donor screening questionnaire
ii. Age ≥ 7 years of age
iii. Agree to undergo laboratory testing for known pathogens within 4 weeks of FMT procedure (screening tests listed below).

*Exclusion:*

i. Potential donors who answer “yes” to and one of the following donor screening questionnaire questions (see Appendix C):

- Are you younger than 7 years of age?
- Do you have known HIV, tuberculosis, Hepatitis B or Hepatitis C infections?
- Have you been exposed to HIV, tuberculosis or viral hepatitis (within the past 12 months)
- Do you engage in any high-risk sexual behaviors (examples: sexual contact with anyone with HIV/AIDS, tuberculosis or hepatitis or engage in sex for money?
- Have you used illicit drugs within the past 3 months?
- Have you had a tattoo or body piercing within the past 6 months?
- Have you ever been incarcerated?
- Have you been to an area with Mad Cow disease (risk factor for Creutzfeld-Jakob disease?)
- Have you traveled within the past 3 months to developing countries?
- Do you have a history of inflammatory bowel disease or chronic diarrhea (i.e. > 3 loose stools daily for the past 3 months?
- Do you have a history of gastrointestinal malignancy or known polyposis?
- Have you used systemic antibiotics within the preceding 3 months?
- Are you currently using any major immunosuppressive medications (e.g. calcineurin inhibitors, systemic anti-neoplastic, exogenous glucocorticoids, biologic agents?)
- Do you have eczema, allergies, or asthma requiring steroids or immunomodulating therapy?
- Do you have autoimmune disease, metabolic syndrome, chronic pain syndrome, neurologic or developmental disorder?
- Do you have contact with hospital patients?
- Have you been hospitalized or in a long term care facility in the past 6 months?
- Do you attend outpatient medical or surgical clinics more than once a month?
- Have you engaged in medical tourism in the past 6 months?

ii. Potential Donors who have positive laboratory testing for any one of the following tests :

- *C. difficile* toxin B
- Salmonella
- Shigella
- Campylobacter
- Shiga toxin (screen for Sorbitol negative E. coli or E. coli O157-H7)
- Vancomycin resistant enterococci (VRE)
- Extended spectrum beta-lactamase-producing Enterobacteriaceae (ESBL)
- Carbapenem-resistant Enterobacteriaceae (CRE)
- Methicillin-resistant Staphylococcus aureus (MRSA)
- Giardia antigen
- Modified acid fast stain for Cryptosporidia, Cyclospora, Isospora
- Ova and parasite (including trichrome)
- HIV, type I and type II screen
- Hepatitis A IgM
- Hepatitis B (HepBs Ag, anti-HepB Core IgM)
- Hepatitis C Ab
- Syphilis RPR
- Syphilis Fluorescent treponemal antibody FTA-ABS
- Tuberculosis (QuantiFERON®-TB Gold)

iii. Donor reports having fever (T > 100.4 $^{o}$F) within two weeks of FMT.
iv. Donor reports of having a runny nose and cough within two weeks of FMT
v. Donor reports ingesting foods that the recipient has an allergy to within one week of FMT
vi. Donor reports having diarrhea within two weeks of FMT.
vii. Donor stool appears grossly abnormal (e.g. presence of blood, etc) prior to processing for colonoscopic instillation.

**5. Protocol for Recipient Involvement (CDI without IBD, CDI with IBD, UC or IC without CDI). (see Appendix D for flow sheet)**

*Pre-FMT events for recipient.*

a. Initial pre clinic visit screening of recipient eligibility. Recipients, families and referring physicians learn about the study through the ClinicalTrials.gov registry and have contacted by e-mail either the research coordinator, Dr. Chawla or Dr. Li for further information on **ClinicalTrials.gov ID:NCT03268213, 479696**. The research coordinator (or Drs. Li and Chawla) reply to the inquiry using a template e-mail that provides further information on the trial (see Appendix E). Pre-screening of recipients is carried out by Drs. Chawla (recipients age <18 y) and Li (recipients age ≥18 y) after they have reviewed the recipients' medical records to determine eligibility. **For recipients in the UC only without CDI group, Dr. Chawla or Dr. Li will contact the primary gastroenterologist to discuss whether the patient's gastroenterologist feels that FMT is an appropriate intervention in this patient, discuss the possibility that the patient could flare post-FMT and to discuss whether the primary gastroenterologist agrees to clinically manage the patient's IBD after FMT**.

   If the recipient is deemed eligible for the study based on pre-screening, he/she or the guardians of the patient will be given the donor screening criteria (Appendix C) and asked to identify a potential donor. The donor is deemed eligible if he/she answers **no** to each question on the checklist. .The recipient and donor will then be scheduled to be seen for formal screening, including a physical exam in Adult or Pediatric GI clinic by either Dr. Li or Dr. Chawla, respectively.

b. Screening and recruitment of recipient in clinic. The recipient will be evaluated and recruited in a face to face encounter with either Dr. Chawla or Dr. Li. At this screening and recruitment clinic visit, additional baseline clinical data will be collected to confirm recipient eligibility (see Appendix F1/F2). The information will include past medical history in terms of previous episodes of *C. difficile* associated disease and/or inflammatory bowel diseases (age of diagnosis, duration, medications) and standard clinical information such as age, sex, race, disease phenotype, medications, and smoking history. The recipient and family members will be instructed on how to prepare for the colonoscopy procedure. If they consent to provide research samples and dietary information, they will be instructed on how to collect the research stool samples, complete the daily food diary one week prior to the FMT samples, reminded to stop antibiotics 48 h before the procedure. The colonoscopic FMT procedure will be scheduled with colonoscopists on the study team, so as to allow for donor screening results to be sent to the study team for review prior to the procedure.

c. 7 days before FMT- initiate recipient daily food diary: The recipients in all three groups will be asked to record their diet using a diary provided by the investigators, during the week that they provide stool samples (Appendix G). The purpose of this is to ensure that the donor does not ingest any foods that may be potentially allergenic to the recipient. The ingested foods may also impact the microbial load of the donor stool

d. 2-5 days before FMT-recipient instruction: The recipients will be contacted by phone to review study instructions. CDI recipients will be reminded again to stop antibiotics used to suppress CDI at least 48h before transplant (Appendix H).

e. 1 day before FMT - Collection of recipient pre-FMT research stool sample: The kits given to the recipient will include gloves, a commode specimen collection system, two specimen containers, biohazard bag and refrigerator packs to keep the stools cold during transport to the endoscopy suite. One of the specimen containers will be empty and the second will be filled with 10 ml RNAlater (Qiagen), an RNA stabilization solution, to better improve recovery of bacterial and human nucleic acids during the transport period.

f. 1 day before FMT- Recipient bowel prep for colonoscopy: The stool recipient will be reminded about instructions for a bowel prep based on the patient's weight. Participants and/or guardians will be told that in order to properly visualize their intestines, they must be "prepped" or flushed of their contents. These medications will cause diarrhea which is the desired effect. The recipient should be in an area with easy access to the bathroom when the medication is given. The goal is to be passing clear fluid without any formed stool. The recipients will be told to call the study coordinator if this does not occur, so that it can be determined whether any additional medications are necessary for preparation**. Once the prep has begun, the recipient will be told to be on a diet consisting of clear liquids only. Clear liquids are those that they can see through and they will be told to avoid red, orange or purple liquids.** Acceptable liquids include water, ginger ale, chicken broth (no chicken or noodles), apple juice, Pedialyte, or jello. **For the last 7 hours prior to the procedure, the recipient should have nothing to eat or drink.** Medications may be taken with a very small amount of water. The colon cleanout prep will be determined by the patient's gastroenterologist. The recipient should be passing clear stools by 5 PM day before FMT. If the recipient is not passing loose to watery clear stools by this time, they will be told to call the study coordinator or principal investigator.

*Day of colonoscopic FMT for recipient.*

g. Day of FMT, undergoing colonoscopic FMT and having research blood sample drawn in SBUMC Endoscopy unit.

*Pre-procedure*. The recipient is consented for colonoscopy by the colonoscopist performing the procedure in the pre-procedure area of the Endoscopy Suite. Female recipients of child bearing age will undergo poc urine pregnancy test, and if positive the procedure will be aborted. Research blood samples (total 20 ml) will be collected either during insertion of the IV in the pre-procedure area, or drawn separately shortly after the procedure, while the patient is still sedated.

*Procedure*. The colonoscopist will record the appearance of the colon to the terminal ileum. In recipients with UC (with or without CDI), the colonoscopist will assign a Mayo endoscopic subscore (0-3, see Appendix I) based on the appearance of the worst affected mucosa in the rectosigmoid area. Upon reaching the ileum or the cecum, the donor stool (50-100 g in 250 to 300 ml of sterile saline, see below for standard operative procedure for preparing stool infusate), will be infused into the ileum and/or the cecum. No other colonoscopic intervention (e.g. biopsy or polypectomy) will performed during the procedure. If there is a colonoscopic finding that the colonoscopist feels needs to be immediately addressed for clinical care by a therapeutic intervention including biopsy, then the FMT will be aborted and the stool infusate will not be administered.

h. *Post procedure*. The recipient will be placed in the right lateral decubitus position and monitored for at least an hour in the post procedure area of the Endoscopy suite, prior to discharge. The recipient will be instructed on collection of the 1 wk and 12 wk post-FMT research stool samples, and be given the stool collection kits to bring home after discharge from the Endoscopy suite. Each recipient will receive a diary card (Appendix J) where he/she or the recipient's guardian will note when the stool recipient experiences a new symptom or exacerbation of current symptoms. This diary card will be reviewed during the followed up phone calls.

*Post-FMT events for recipient*.

i. 1 day post-FMT. The recipient and/or guardian are contacted by phone by the study team in order to complete the Day after Transplant Case Report Form (see Appendix K).

1st week post-FMT collection of recipient food diary and 1 wk post-FMT research stool sample. The recipient/family will complete a one week daily food diary during the week post-transplant and collect the 1 wk-post FMT research stool sample which will be shipped overnight in refrigerator packs to Dr. Li's research lab.

j. 1 week to 12 weeks post FMT. - weekly phone follow up of recipient. The recipient and/or recipient's (pediatric recipient) guardian are contacted by phone by the study team weekly during the first 12 weeks post-FMT to complete the Follow up Phone Call/Adverse Event Form (Appendix L1 for CDI recipient without IBD, Appendix L2 for UC or IC recipient with or without CDI). The forms contain a prompt to inform the Principal Investigator of any adverse events. Patients will be triaged to either the Emergency Department or to Adult or Pediatric Gastroenterology Clinic if any concerning adverse events occur. Any serious adverse events (Grades 3,4,5) will be documented (Appendix L), evaluated by the Principal Investigator and reported immediately to IRB as well as Safety Monitoring Committee.

k. 12 week post-FMT - collection of recipient food diary and 12 week post-FMT research stool sample. . The recipient/family will complete a one week daily food diary during the week post-transplant and collect the 12 wk-post FMT research stool sample which will be shipped overnight in refrigerator packs to Dr. Li's research lab.

l. 3-4 mo post FMT flexible sigmoidoscopy. If the primary gastroenterologist caring for the patient, schedules the patient for a follow up flexible sigmoidoscopy, we will request that the colonoscopist obtain a Mayo endoscopic subscore during the procedure.

m. 4 mo to 12 mo post-FMT – monthly phone call follow up of recipient. The recipient and/or recipient's (pediatric recipient) guardian are contacted by phone by the study team monthly between 4 and 12 months post-FMT to complete the Follow up Phone Call/Adverse Event Form (Appendix L1 for CDI recipient without IBD Appendix L2 for UC or IC recipient with or without CDI). Any serious adverse events (Grades 3,4,5) will be documented (Appendix M), evaluated by the Principal Investigator and reported immediately to IRB as well as Safety Monitoring Committee.

**6. Protocol for Donor Involvement. (See Appendix D for flow sheet)**

*Pre-FMT events for donor.*

a. Screening and recruitment of potential donor in clinic. Once the recipient has been entered into the study, the recipient will identify the donor. If the donor is age > 18 y will be screened and evaluated by Dr. Li in the Adult Gastroenterology Clinic (see Research Site, section D.3) with a complete history and physical to ascertain eligibility. If the donor is age ≥7 y and ≤18 y, the donor will be screened and evaluated by Dr. Chawla in the Pediatric Gastroenterology Clinic (see Research Site, section D.3) with a complete history and physical to ascertain eligibility. The donor will be consented to participate in the study, which requires undergoing stool and serological testing to assess for infectious pathogens that may theoretically be passed from the donor to the recipient. The donor (as well as the recipient) will sign an acknowledgement form that if medical insurance does not cover the costs of these tests, the costs of the donor screening tests will be borne by the recipient or donor (Appendix N). (The study team is in the process of applying for research grant funding that will cover the costs of donor screening for pathogens, but currently there are no research funds available to cover these costs). Once the donor has consented to the study, the donor will be instructed on collecting the stool to be used for the transplant, completing the one week food diary prior to providing the sample. Dr. Li or Dr. Chawla will order the following laboratory tests for the donor to complete before the colonoscopic FMT is performed:

*Stool tests*:
- *C. difficile* toxin PCR (CPT 87493)
- Routine stool culture screen (CPT 87046) - includes Salmonella, Shigella, Campylobacter, Sorbitol negative E. coli or E. coli O157-H7 (Shiga toxin)
- Giardia Ag EIA (CPT 87328)
- Modified acid fast stain of stool (CPT 87015/87207) Cryptosporidia, Cyclospora,
- Ova and parasite with trichrome (CPT 87177) includes Isospora
- Vancomycin-resistant enterococci (VRE), (CPT 87081)
- Extended spectrum beta-lactamase-producing Enterobacteriaceae (ESBL), (CPT 87184)
- Carbapenem-resistant Enterobacteriaceae (CRE) ,(CPT 87081)
- Methicillin-resistant Staphylococcus aureus (MRSA), (CPT 87081)

*Blood tests*:
- HIV type 1 and type 2 screen (CPT 86701/86702)
- Acute Viral Hepatitis Panel: Hepatitis A IgM, Hepatitis Bs Ag, Hepatitis Bc IgM, Hepatitis C Ab (CPT 80074)
- Syphilis Rapid Plasma Reagin or RPR (CPT 86592)
- Fluorescence Treponemal Ab or FTA-Abs (CPT 86781)
- Tuberculosis (QuantiFERON®-TB Gold) (CPT 86480)

The laboratory test results must be dated within 4 weeks of the day the colonoscopic FMT is performed. The investigators are in the process of applying for grant support in order to cover the costs of donor screening, since this is not universally covered by medical insurance.

If the donor tests positive for a potential pathogen, Dr. Li or Dr. Chawla will inform the donor of the test results and will ask for the contact information for the donor's PCP to forward the test results to. The donor will be counseled to follow up with PCP to discuss the test results and the need for any further testing or treatment.

b. 7 days before FMT - initiate donor daily food diary. The donor will initiate recording daily food diary and avoid ingesting any foods the recipient is allergic to.

c. 2-5 days before FMT – donor instructions. The study team will contact the donor by phone to check that the donor has initiated the daily food diary, has not been febrile T > 100.4$^{o}$ F within 2 weeks of the procedure, and has avoided ingesting any of the recipient's food allergens during the week prior to the transplant (see Appendix O).

d. 1-day before FMT – initiate collection of donor stool. The donor will have been given two stool collection kits at the initial clinic visit. The donor is instructed to collect one stool within 24 h before the procedure and to keep the stool refrigerated with the refrigerator packs. If possible, the donor is instructed to collect a stool within 6 h before the procedure and kept the second stool refrigerated with the refrigerator packs. The donor will also have been instructed to aliquot research stool samples at both time points into specimen jars, one plain and the other filled with 10 ml of RNAlater as described for the recipient in section **D.5.e**. The donor is instructed to bring, or have the recipient bring both stool samples to the Endoscopy Suite for the research team to pick up 2 h before the FMT procedure is scheduled.

e. Day of FMT. – Collection and evaluation of donor stool
*Preprocedure*. The donor (or the recipient on behalf of the donor) will deliver the stools to the Endoscopy Suite for the research study team to pick up and process 2 h before the FMT procedure. The study team will complete Day of Transplant Donor Checklist Form (Appendix P) either face to face or by phone contact with the donor, to make sure no donor exclusions apply, prior to processing the stool to prepare the stool infusate, as described in the next section.

ii. **Protocol for preparation of donor stool infusate for colonoscopic infusion.**
The donor stool (refrigerated not frozen) is transported by the study team to Dr. Li's laboratory and processed for infusion via colonoscopy following standard operating procedures as previously described:[44]

- Stool preparation will occur in the biological safety hood
- Universal precautions will be adhered to. Those involved with mixing and/or handling the fecal transfusion material will wear a fluid-resistant gown, gloves, and mask with goggles, or eye shield
- Donor stool will be transferred to a single-use disposable container
- Preservative-free normal saline (250-500 mL) at room temperature will be used to dilute the stool sample until it reaches a liquid slurry consistency
- The stool slurry will then be filtered to remove as much particulate matter as possible. This will be accomplished using gauze pads lining the inside portion of a plastic disposable funnel
- The filtered stool slurry will be divided into aliquots of 50 mL and will be used within an hour of preparation

iii. **Protocol for Processing and Analysis of Recipient and Donor Samples:** As noted in sections 4 and 5, blood and stool samples may be collected from the recipient and from the donor for further analysis. Those recipients and donors who have agreed to provide clinical data, blood and stool samples are consented under a separate consent under a separate IRB ((IRB net ID: 163184-15) for collection and storage of samples within the Stony Brook GI satellite Biobank, which operates as a module under the Stony Brook Biobank New York State License for Biobanking. The clinical data, stripped of patient health identifiers, will be assigned a patient ID and a visit ID and will be stored in the Stony Brook University Digestive Diseases Research Tissue Procurement Facility clinical database. The patient samples will also be stripped of patient health identifiers and assigned a patient ID, visit ID and sample ID and will be archived within the Stony Brook University GI Biobank. The analysis of the samples and downstream products will including the following:

a. Genotyping for IBD risk alleles**.** DNA will be extracted from peripheral mononuclear blood cells (in blood) as previously described in Dr. Li's laboratory.[43] The DNA will be assigned the same patient ID, visit ID and sample ID from the original sample. We have used Taqman PCR assays, Sequenom, and array methods for analyzing genotypes thus far, but the technology is rapidly evolving.[22] We will seek the most cost effective means of obtaining accurate genotyping of established IBD risk alleles. If we submit the samples for genotyping at another facility, each of these samples will be assigned another sample ID, to further protect the privacy of the subjects, prior to shipping these samples to an outside facility.

b. 16S rRNA sequencing. 16S rRNA sequencing will provide a first tier of microbiome surveillance and will measure taxa based microbial diversity and composition.[43] This may involve shipping stool DNA to outside facilities for library construction and sequencing (University of Colorado). If we submit these DNA samples to outside institutions for analysis, each of these samples will be assigned another sample ID, to further protect the privacy of the subjects, prior to submission of these samples to an outside facility. For all rRNA sequence datasets, we routinely screen for and remove contaminating human DNA/RNA sequences by use of bowtie2 read-mapping software; thus, no human sequences will be analyzed beyond this step or will be included in public repositories. We propose to measure the alterations in microbial diversity and composition before and after fecal microbial transplantation.

c. Targeted qPCR sequencing. To detect targeted microbial taxa and functions we will utilize targeted qPCR and reverse transcriptase qPCR. These analyses will be carried out in Dr. Li's and Dr. Gathungu's laboratory.[46]
d. Fecal calprotectin. Fecal calprotectin is a marker of intestinal inflammation that will complement the colonoscopic scores and patient questionnaires. We propose to measure fecal calprotectin in each of the collected stools using the PhiCal commercial ELISA kit (CALPRO, Oslo, Norway) according to the manufacturer's instructions within Dr. Li's laboratory. Frozen stools archived at -80$^o$C will be batch extracted and analyzed along with the manufacturer's control to avoid interassay variability.
e. Shotgun metagenomics, bacterial metatranscriptomics, fecal metabolomics, fecal proteomic. We propose to conduct multiple omic studies on microbial function using the collected stool samples if we are successful in obtaining outside funding to support these studies. This may require sending aliquots of samples or their downstream products to other institutions for analysis. If so, the samples will be assigned another sample ID to further protect the privacy of the subjects before shipping these samples out to other institutions. For all meta-omic sequence datasets, we routinely screen for and remove contaminating human DNA/RNA sequences by use of bowtie2 read-mapping software; thus, no human sequences will be analyzed beyond this step or will be included in public repositories.

#### E. STATISTICS:

The number of patients enrolled will not be limited since a primary goal of this study is to establish safety, tolerability and efficacy of FMT. It is anticipated that data from 80 FMTs with a balanced design of 20 FMTs performed on each of the following four groups of recipients (CDI without IBD, CDI with UC, CDI with CD, and UC without CDI) associated with this IRB protocol (ClinicalTrials.gov ID:NCT03268213, 479696, IND for UC only 15642), in combination with data from 20 FMTs performed on CD without CDI recipients recruited through a sister protocol (IRB 973349-5, ClinicalTrials.Gov ID NCT03267238, IND16795) will be needed to generate a power analysis on the multi-omic datasets. A linear mixed model will used to compare alpha-diversity (ShannonH) and beta-diversity (Bray-Curtis and Jaccard distance) between each time point (donor, pre-FMT, 1 week and 3 month post-FMT) and each disease group as previously described.[46] The two-way interaction term, Group*FMT, will be used to estimate the differences between any two time points within a specific disease group. Covariance structure to model correlation among measurements from the same patient and his/her corresponding donor will be selected based on Akaike Information Criteria (AIC). Possible covariance structures to model correlation among longitudinal measurement from the same patient and measurement in the corresponding donor included unstructured (UN), compound symmetry (CS), Toeplitz (TOEP), Heterogeneous CS (CSH) and Heterogeneous TOEP (HTOEP). Using the two-way interaction term between Group and FMT, generalized linear mixed models (GLMMIX) will be used to examine the possible change in individual OTU relative abundance after FMT in recipients and such difference between recipient and donor over time. The actual counts of each OTU will be assumed to follow a negative binomial distribution.[47] The log-transformed overall sequence count for each individual at each time point will be considered as an offset. In case that there is model converging issue GLMMIX, generalized estimating equation (GEE) will be used. In GEE, the dependence structure was chosen based on Quasi Information Criteria (QIC). The p-values from comparison analysis of all OTUs will adjusted for multiple comparisons by the Bonferroni correction or by the Benjamin-Hochberg method (FDR < 0.05). Principal coordinate analyses will be carried out using the R package *vegan*.

Similar models will be used to examine the difference in the expression level of a specific gene or protein based on metatranscriptomic data and metabolomic data between recipient and donor and the change in such difference over time. Beyond analysis of individual data-types integr-omic analyses that combine all data types (i.e. metabolomics, microbiome, and clinical) will rely on the use of Similarity Network Fusion,[47] which performs unsupervised clustering of patients based on each datatype and then fuses the obtained networks, strengthening links in complementary networks. These analyses will reveal the relationships among taxonomic composition, microbiome diversity and function and metabolomics which implies the mechanism how FMT works. All analysis will be performed in SAS 9.4 (SAS institute Inc., Cary, NC) and R 3.4.0 (R Foundation for Statistical Computing, Vienna, Austria).

#### F. FUNDING STATUS, DETAILS:

There is currently no funding for the procedure or evaluation of the recipient and donors in clinic, or the laboratory screening of the donor. The Simons Foundation Award is currently used to cover the costs of processing, sequencing and analysis of stool and blood samples.

#### G. HUMAN SUBJECTS RESEARCH PROTECTION FROM RISK

##### 1. Risk to Subjects:

The risks to the recipient and the donor are described separately.

a. Risk to recipient.

i. *Risk of recipient undergoing colonoscopy*. The risks associated with the colonoscopy are infection, bleeding, and < 0.1% chance of perforation, which may require surgery. Additionally, there are risks associated with receiving anesthesia during colonoscopy including anaphylaxis, aspiration and respiratory distress.

ii. *Risk of undergoing FMT in recipient with recurrent CDI without IBD*. Risks to FMT include bloating/flatulence, abdominal pain/cramping, diarrhea, blood in the stool, fatigue, and fever.

iii. *Risk of undergoing FMT in recipient with recurrent CDI with IBD.* Risks to FMT include bloating/flatulence, abdominal pain/cramping, diarrhea, blood in the stool, fatigue, and fever. In addition, there is a risk that even if the C. difficile is treated that there may be exacerbation of the underlying IBD.[25]

iv. *Risk of undergoing FMT in recipient with UC or IC without CDI*. Risks to FMT include bloating/flatulence, abdominal pain/cramping, diarrhea, blood in the stool, fatigue, and fever. In addition, there is a risk that there may be continued progression or exacerbation of the underlying IBD.[25]

v. *Collection of stool and blood from recipient*. These are minimally invasive procedures. The risks of collecting blood are infection at the site of needle stick and bruising.

vi. *Risk of collecting recipient clinical data*. There is an extremely rare possibility that the patient data can be shared in a way that identifies the recipient. Great lengths will be made to restrict access to patient and sample codes.

b. Risk to donor

i. *Collection of stool from recipient*. This is minimally invasive procedure.

ii. *Risk of collecting recipient clinical data*. There is an extremely rare possibility that the clinical data can be shared in a way that identifies the donor. Great lengths will be made to restrict access to patient and sample codes.

##### 2. Adequacy of Protection Against Risks:

The clinical data and stool samples will be stripped of patient health identifiers and assigned a patient code and sample code. The subject's participation will end after the collection of stool and clinical data is completed 1 year after enrollment in the study. We are only requesting that we be able to collect clinical metadata by prospective questionnaires and reviewing the medical records, and to collect recipient fecal and blood samples before FMT and after FMT. The consents will be obtained by the physicians or research staff, who have been trained in good clinical practices and are highly experienced in providing and obtaining informed consent in this patient population.

Additionally, all recipients will have adverse event monitoring as outlined below:

*Day of fecal microbiota transplantation:*

a. During the colonoscopy procedure: an anesthesiologist will be assigned to the recipient and the endoscopy unit will have full-code preparedness

b. Post colonoscopy procedure: the recipient will be monitored in the post-operative area of the endoscopy suite for one hour by continuous monitoring

*Post fecal microbial transplant*ation

The study team will closely monitor the recipient by phone call and an algorithm will be attached to the checklist to refer patients to either clinic or the Emergency Department if necessary.

a. 1 day post-transplant: The transplant recipient will receive a phone call the day after the procedure and questions will be asked from a standardized checklist containing the following items:

i. General well being

ii. Presence of abdominal pain

iii. Presence of temperature greater than 100.4 degrees Fahrenheit
iv. Presence of diarrhea (loose, watery stools) or blood in the stool
v. Presence of new rash

b. Weekly monitoring 1-12 weeks post transplant: All recipients will have weekly monitoring via telephone call for 12 weeks. This will include the following additional questions:
i. Has diarrhea stopped or improved?
ii. If the diarrhea had stopped, is there a recurrence of diarrhea?
iii. If the diarrhea has not improved, have you tried other treatments?
iv. Has there been any need of antibiotics since the transplant?
v. Did any medical condition you had before your fecal transplant go away after your fecal transplant? (for example, arthritis and chronic skin rash)
vi. Has abdominal pain if present prior to fecal microbiota transplant resolved?
vii. Have you developed any new medical conditions since the fecal transplant?

c. Monthly monitoring 4-12 months post transplant: All recipients will have monthly monitoring via telephone call between 4 and 12 months after the first twelve weeks.
d. Recipients and/or their guardians will be advised to call either the Pediatric Gastroenterology or Adult Gastroenterology offices for any questions or health related concerns for up to 1 year post-transplant.
e. The study team will meet with Drs. Chawla and Li on a monthly basis (last Wednesday of every month) to review follow up phone calls.

**3. Potential Benefits of Proposed Research to the Subjects and Others:**

The potential benefits of having the fecal transplantation for treatment of recurrent or refractory *CDI* not responsive to standard medical therapy is prevention of recurrent CDI.

The potential benefits of having the fecal transplantation for treatment of recurrent or refractory CDI not responsive to standard medical therapy, in addition to prevention of recurrent CDI is improvement in their IBD symptoms.

The potential benefits of having the fecal transplantation for treatment of UC or IC without CDI is improvement in their IBD symptoms.

Conducting research on these samples, including the research proposed in this application may create new tests, treatments, or cures. If it does the subjects (recipients or donors) who have donated samples will not receive any money from those products.

**4. Importance of the Knowledge to be Gained:**

This study will enable us to contribute to the literature regarding the safety, efficacy, and tolerability of fecal microbiota transplantation. Additionally, by collecting well phenotyped linked samples, future integrations of the microbiome data not only with phenotype but also with genetic data and metabolomics data will be possible. This will enable further studies investigating the mechanism(s) by which gut bacteria can affect susceptibility to medication refractory *Clostridium difficile* and/or ulcerative colitis or indeterminate colitis.

**H. SAFETY MONITORING COMMITTEE (for more than minimal risk studies):**

A safety monitoring committee (SMC) has been established and is composed of the three following experts:

a) Sharon Nachman, M.D., Chair of Safety Monitoring Committee, Professor of Pediatrics, Chief, Division of Infectious Diseases, Stony Brook University School of Medicine. Expertise in clinical trials.
b) Roy Steigbigel, M.D., Professor of Medicine, Pathology, Pharmacology and Microbiology, Division of Infectious Diseases, Stony Brook University School of Medicine. Expertise in infectious diseases.
c) Matthew Ciorba, M.D., Associate Professor of Medicine, Director of Research in the IBD Program Division of Gastroenterology and Hepatology, Washington University School of Medicine. Clinical and research expertise in the microbiome in inflammatory bowel diseases and colon cancer.

The committee will be asked to perform an interim assessment of safety 4 weeks after every ten subjects are enrolled in the study and undergo fecal transplant. This will help in determining whether or not this study protocol should continue to be implemented or requires modifications.

Adverse events (AE) will be recorded by follow up phone calls as per the following grading (Division of AIDS (DAIDS) Table for Grading the Severity of Adult and Pediatric Adverse Events:[50]

Grade 1 Mild; asymptomatic or mild symptoms; clinical or diagnostic observations only; intervention not indicated.
Grade 2 Moderate; minimal, local or noninvasive intervention indicated; limiting age-appropriate instrumental Activities of Daily Living (ADL).
Grade 3 Severe or medically significant but not immediately life-threatening; hospitalization or prolongation of hospitalization indicated; disabling; limiting self-care ADL.
Grade 4 Life-threatening consequences; urgent intervention indicated.
Grade 5 Death related to AE.

Grades 3, 4 and Grade 5 adverse events will trigger the team to request a safety monitoring committee review of the data and pause further enrollment.

Adverse events will be recorded when follow-up phone calls are made. Intensity and relationship of adverse events with FMT will be described using Common Terminology Criteria for Adverse Events (CTCAE version 4.0) and Toxicity Grading Guidance from Vaccine Clinical Trials (U.S. Food and Drug Administration, September 2008).

Four weeks after every 10th subject's enrollment and completion of fecal transplant a cumulative report will be submitted to SMC for formal review. SMC will have 1 week to respond. During this period there will be a safety hold for the Ulcerative colitis subjects. If no issues are identified, the study will be reopened and further enrollment will be allowed.
After the data on the next 10 subjects have been reviewed by SMC and no concerns are raised, the hold period will no longer be necessary.

Principal investigator may request an ad hoc SMC review if needed at any time point.

There will be no safety hold on enrollment of *Clostridium difficile* subjects for FMT procedure as this procedure has been widely accepted in the medical field and comparable to standard of care procedures for refractory *Clostridium difficile*. FDA does not require an IND status for FMT treatment of refractory *C. Difficile*. The committee may also require changes to the protocol to ameliorate the safety concerns that pose significant risk to stool recipients. In the absence of protocol changes, the SMC must follow the protocol-specified stopping rules.

**APPENDIX A**

### Stool Recipient Screening Questionnaire (C. difficile) with or without IBD

Recipient Name:______________________ Date:___________

Inclusion: Recipient is deemed eligible if any one of the following criteria are met:

___ Patient is 7 years of age or older with at least two recurrences (total three CDI infections) of mild to moderate C. difficile (<6 diarrheal stools/day) diagnosed by positive toxin PCR or EIA after completing standard medical therapy with metronidazole, vancomycin or fidaxomicin.

___ Patient is 7 years of age or older with at least two episodes of severe C. difficile infection (>6 diarrheal stools/day requiring hospitalization and associated with significant morbidity).

___ Patient is 7 years of age or older with moderate C. difficile infection (3-6 diarrheal stools/day not responding to successive standard therapy, e.g. metronidazole, vancomycin and/or fidaxomicin) lasting at least 28 days.

___ Patient is 7 years of age or older with severe and/or fulminant C. difficile colitis (> 6 diarrheal stools/day) with no response to standard therapy after 48 hours.

Exclusion: Recipient is deemed ineligible if the answer to any of the following questions is yes:

| Question | Yes | No |
|---|---|---|
| Is the patient younger than 7 years old? | | |
| Is the patient scheduled for abdominal surgery within the next 12 weeks? | | |
| Has the patient had major abdominal surgery within the past 3 months? | | |
| Is the patient pregnant (if applicable)? | | |
| Does the patient have a Hgb < 6 g/dL? | | |
| Is the patient's absolute neutrophil count less than 1500/mm$^3$? | | |
| Does the patient have a known diagnosis of graft vs. host disease? | | |
| Has the patient used an investigational drug within the past 2 months? | | |
| Has the patient used a TNFα agonist within the past 2 weeks? | | |
| Has the patient been diagnosed with Bacteremia within the past 4 weeks? | | |

_______________________________ _______________________________

Name of person completing this form Signature of person completing this form

**APPENDIX B**

### Stool Recipient Screening Questionnaire (Ulcerative Colitis or Indeterminate Colitis)

Recipient Name:______________________ Date:___________

Inclusion: Recipient is deemed eligible if any one of the following criteria are met**.**

___ Patient is 7 years of age or older and has been treated with steroid therapy for at least one month.

___ Patient is 7 years of age or older and has been treated with immunomodulatory therapy for at least one month

___ Patient is 7 years of age or older and has been treated with biological therapy for at least one month.

Exclusion: Recipient is deemed ineligible if the answer to any of the following questions is yes:

| **Question** | **Yes** | **No** |
|---|---|---|
| Is the patient younger than 7 years old? | | |
| Is the patient scheduled for abdominal surgery within the next 12 weeks? | | |
| Has the patient had major abdominal surgery within the past 3 months? | | |
| Is the patient pregnant (if applicable)? | | |
| Does the patient have a Hgb < 6 g/dL? | | |
| Is the patient's absolute neutrophil count less than 1500/mm$^3$? | | |
| Does the patient have a known diagnosis of graft vs. host disease? | | |
| Has the patient used an investigational drug within the past 2 months? | | |
| Has the patient used a TNFα agonist within the past 2 weeks? | | |
| Has the patient been diagnosed with Bacteremia within the past 4 weeks? | | |
| Has the patient had any previous FMT? | | |

______________________________ ______________________________
Name of person completing this form Signature of person completing this form

#### APPENDIX C
### Stool Donor Screening Questionnaire

Donor Name:______________________ Date:___________

| Question | Yes* | No |
|---|---|---|
| **1**. Are you younger than 7 years of age? | | |
| **2.** Do you have known HIV, tuberculosis, Hepatitis B, or C infections? | | |
| **3**. Have you been exposed to HIV, tuberculosis or viral hepatitis (within the previous 12 months)? | | |
| **4**. Do you engage in any high-risk sexual behaviors (examples: sexual contact with anyone with HIV/AIDS, tuberculosis or hepatitis, sex for drugs or money)? | | |
| **5**. Have you used illicit drugs within the past 3 months? | | |
| **6**. Have you had a tattoo or body piercing within the past 6 months? | | |
| **7**. Have you ever been incarcerated? | | |
| **8**. Have you been to an area with Mad Cow Disease (risk factor for Creutzfeld-Jakob disease)? | | |
| **9**. Have you traveled (within the last 3 months) to developing countries? | | |
| **10**. Do you have a history of inflammatory bowel disease or chronic diarrhea (i.e. greater than 3 loose stools daily for the past 3 months)? | | |
| **11**. Do you have a history of gastrointestinal malignancy or known polyposis? | | |
| **12**. Have you used systemic antibiotics in the past 3 months? | | |
| **13**. Are you currently using any major immunosuppressive medications (e.g., calcineurin inhibitors, systemic anti-neoplastic, exogenous glucocorticoids, biologic agents)? | | |
| **14.** Do you have eczema, allergies or asthma requiring steroids or immune-modulating therapy? | | |
| **15.** Do you have an autoimmune disease, metabolic syndrome, chronic pain syndrome, neurologic or developmental disorder? | | |
| **16.** Do you have contact with hospital patients? | | |
| **17.** Have you been hospitalized or in a long term care facility in the past 6 months? | | |
| **18.** Do you attend outpatient medical or surgical clinics more than once a month? | | |
| **19.** Have you engaged in medical tourism in the past 6 months? | | |

***Answer of "yes" to any of the questions results in exclusion.**

______________________________ Name of person completing this form

______________________________ Signature of person completing this form

### Stool Donor Screening Questionnaire

Donor Name:______________________ Date:___________

| Did the Donor Test Positive For: | Yes | No |
|---|---|---|
| **1**. Clostridium difficile toxin A or B? | | |
| **2**. Stool culture? | | |
| **3**. Giardia antigen? | | |
| **4**. Cryptosporidium antigen? | | |
| **5**. Ova and parasites? | | |
| **6**. Acid-fast stain for Cyclospora, Isospora? | | |
| **7**. HIV, type 1 and 2? | | |
| **8**. Hepatitis A (HAV IgM)? | | |
| **9**. Hepatitis B (HBsAg, anti-HBc, anti-HBs)? | | |
| **10**. Hepatitis C (HCV Ab)? | | |
| **11**. Syphilis (RPR and FTA-ABS)? | | |
| **12.** Sorbitol negative E. Coli? | | |
| **13.** Tuberculosis? | | |
| **14.** Vancomycin resistant enterococci (VRE) | | |
| **15.** Extended spectrum beta-lactamase-producing Enterobacteriaceae (ESBL)? | | |
| **16.** Carbapenem-resistant Enterobacteriaceae (CRE)**?** | | |
| **17.** Methicillin-resistant Staphylococcus aureus (MRSA)? | | |

***Answer of "yes" to any of the questions results in exclusion.**

________________________________ ________________________________

Name of person completing this form Signature of person completing this form

**APPENDIX D:**

### Study Flowsheet

| Protocol | Day(s) | | |
|---|---|---|---|
| Recruitment of stool recipient (Appendix F1 for recurrent CDI ±IBD) (Appendix F2 for UC or IC without IBD) | -28 | Performed? | Yes / No |
| Donor screening (Appendix C) | -28 | Eligible? | Yes / No |
| Donor Serum and Stool Screening | -28 | Eligible? | Yes / No |
| Diet Record completed by Recipient? (Appendix G) | -7 to 0 | Yes | No |
| Diet Record completed by Donor? (Appendix G) | | Yes | No |
| Pre-transplant stool collection? | -3 to -1 | Yes | No |
| Recipient has stopped antibiotics | -3 to -2 | Yes | No |
| Recipient undergoing bowel prep with resultant clear yellow stool? | -1 | Yes | No |
| Donor stool processed? (Appendix P) | 0 | Yes | No |
| Day of transplant stool collection? | 0 | Yes | No |
| Recipient blood obtained? | 0 | Yes | No |
| Patient monitored? | 0 | Yes | No |
| Follow-up Phone Call? (Appendix L1 for recurrent CDI ±IBD ) (Appendix L2 for UC or IC) | 1 | Yes | No |
| **Protocol** | **Week (s)** | | |
| Follow-up Phone Call? (Appendix L1 for recurrent CDI ±IBD ) (Appendix L2 for UC or IC) | 1 | Yes | No |
| Post-transplant diet record? | | Yes | No |
| Post-transplant stool collection? | | Yes | No |
| Follow-up Phone Call? (Appendix L1 for recurrent CDI ±IBD ) (Appendix L2 for UC or IC) | 2 | Yes | No |
| | 3 | Yes | No |
| | 4 | Yes | No |
| | 5 | Yes | No |
| | 6 | Yes | No |
| | 7 | Yes | No |
| | 8 | Yes | No |
| | 9 | Yes | No |
| | 10 | Yes | No |
| | 11 | Yes | No |

| | | | |
|---|---|---|---|
| Follow-up Phone Call?<br>(Appendix L1 for recurrent CDI ±IBD )<br>(Appendix L2 for UC or IC) | 12 | Yes | No |
| Post-transplant diet record? | | Yes | No |
| Post-transplant stool collection? | | Yes | No |
| **Protocol** | **Month (s)** | | |
| Follow-up Phone Call?<br>(Appendix L1 for recurrent CDI ±IBD )<br>(Appendix L2 for UC or IC) | 4 | Yes | No |
| | 5 | Yes | No |
| | 6 | Yes | No |
| | 7 | Yes | No |
| | 8 | Yes | No |
| | 9 | Yes | No |
| | 10 | Yes | No |
| | 11 | Yes | No |
| | 12 | Yes | No |

**APPENDIX E**

**TEMPLATE E-MAIL IN RESPONSE TO INQUIRY ABOUT NCT03268213, 479696,**

Dear -

Thank you for your interest. I am referring you to
(IF ADULT RECIPIENT) Dr. Ellen Li, Professor of Medicine/Gastroenterology,Stony Brook Medicine
(IF PEDIATRIC RECIPIENT) Dr. Chawla, Chief and Professor of Pediatrics/ Gastroenterology, Stony Brook Medicine.
We are currently open to enrollment and accept local and out-of-state candidates.

However, we need your gastroenterologist to be on board with this study, which is strictly observational and simply asking if we can change your microbiome. In addition, we need you to select a healthy donor who fulfills the attached screening criteria. The evaluation of both the patient (recipient of the transplant) and donor takes place at Stony Brook Medicine, NY. So, this requires two trips to Stony Brook. One for the initial recruitment and the second for the colonoscopic delivery of the transplant. We do not cover travel costs, donor screening costs or the cost of the colonoscopy (usually covered by your health insurance company). However, costs of analyzing the stools for all microbiome-related studies will be covered by us.

We prefer no change in your medications and no biologic therapy for at least 2 weeks prior to the transplant.

Please contact your gastroenterologist. If they are in agreement to proceed and you wish to move ahead, please send us the name, e-mail and fax of your primary gastroenterologist. The last office visit note should be sent to us via fax

(IF ADULT RECIPIENT) (631-444-5225) ATTN: DR. ELLEN LI or email:


(IF PEDIATRIC RECIPIENT) (631-444-6045) ATTN: DR. ANUPAMA CHAWLA or email:


It should also include the most recent colonoscopy, pathology and imaging reports.

Please see the attached documents, which you can share with your gastroenterologist.
1. Recipient Permission/Consent form
2. Recipient Screening form
3. Donor Permission/Consent form
4. Donor Screening form
5. Cost Discussion form
6. Food Diary

Regards,
Katherine Markarian
Research Coordinator
631-444-3868

Approved: 6/10/2021
Expiration Date: 6/9/2022
Stony Brook University Human Subjects Committee (IRB)

**APPENDIX F1 (*Clostridium difficile with or without IBD*):**

### STUDY PATIENT CHARACTERISTICS

#### Within 2 weeks prior to Fecal microbial transplantation

RECIPIENT CODED No.: ______________________________ SEX: ☐M/☐F
DOB: ___ ___/___ ___/___ ___
Assigned Code Number: ____
WEIGHT: ____ KGS BMI: ____
HEIGHT: ____ CMS

| | | | |
|---|---|---|---|
| **1.** Duration of symptoms related to *C. difficile* | ____ Month(s) | | |
| **2.** *C. difficile* PCR and/or toxin date(s) | __ __/__ __/__ __<br>__ __/__ __/__ __ | | |
| **3.** Antibiotics for *C. difficile* the patient has been treated with: | Name | | Duration (days) |
| **4.** Probiotics for *C. difficile* the patient has been treated with: | Name | | Duration (days) |
| **5.** Diarrhea (BMs/day): | <3 | 3-6 | >6 |
| **6.** Blood in stool: | Yes | No | |
| **7.** Mucous in stool: | Yes | No | |
| **8.** Abdominal pain (Scale 0-10) | ___/10 | | |
| **9.** Fatigue | Yes | No | |
| **10.** Weight loss during illness | ___KGS | | |
| **11.** Previous abdominal surgery | Yes<br>Location:___________ | No | |
| **12.** Smoker? | Yes<br>Packs/day:__________ | No | |

Approved: 6/10/2021
Expiration Date: 6/9/2022
Stony Brook University Human Subjects Committee (IRB)

**APPENDIX F2 (Ulcerative Colitis or Indeterminate Colitis with or without CDI):**

### STUDY PATIENT CHARACTERISTICS

**Within 2 weeks prior to Fecal Transplantation**

RECIPIENT CODED No.: ________________________ SEX: ☐M/☐F
DOB: ___ ___/___ ___/___ ___
Assigned Code Number: ____
WEIGHT: ____ KGS BMI: ____
HEIGHT: ____ CMS

| | | | |
|---|---|---|---|
| **1.** Duration of symptoms related to Ulcerative colitis or indeterminate colitis | ____ Month(s) | | |
| **2.** Date of IBD diagnosis | __ __/__ __/__ __ | | |
| **3.** Current medications | Name | | |
| **4.** Medications for IBD the patient has been treated with: | Name | | Duration (days) |
| **5.** Probiotics the patient has been treated with: | Name | | Duration (days) |
| **6.** Diarrhea (BMs/day): | <3 | 3-6 | >6 |
| **7.** Blood in stool: | Yes | | No |
| **8.** Mucous in stool: | Yes | | No |
| **9.** Abdominal pain (Scale 0-10) | ___/10 | | |
| **10.** Fatigue | Yes | | No |
| **11.** Weight loss during illness | Yes ____KGs | | No |
| **12.** Previous abdominal surgery | Yes Location___________ | | No |
| **13.** Smoker? | Yes Packs/day:___________ | | No |

Approved: 6/10/2021
Expiration Date: 6/9/2022
Stony Brook University Human Subjects Committee (IRB)

**APPENDIX G**

### Stony Brook University Medical Center Food Diary

**Coded Number:** _______________

**Physician:** _____________________

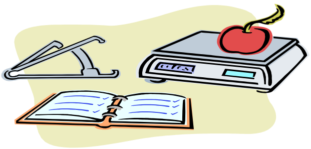

**Record all food, fluid, supplement intake including Probiotics**
**Record all condiments, sauces, oils, etc.**

**Date:**

| MEAL | FOOD: Brand/Description (i.e. Whole milk, rye bread) | HOW WAS IT PREPARED (i.e. 3 ounces fried chicken) |
|---|---|---|

****Record each day on separate sheet****

**APPENDIX H**

#### Checklist for instructing Recipient 2-5 days prior to Fecal Microbial Transplantation

RECIPIENT CODED No.: ________________________________
DATE/TIME OF CALL: ___ ___/___ ___/___ ___:___AM/PM

RECIPIENT DATA:

| | | |
|---|---|---|
| **1.** Do you fully understand the colonoscopy prep procedure? | YES | NO |
| **2.** Have you completed tapering of any antibiotics (if applicable)? It is important that antibiotics must be stopped 48 h before the transplant because the antibiotics will kill the bacteria in the transplant* | YES | NO |
| **3.** Do you fully understand the stool collection instructions & kit? | YES | NO |
| **4.** Will you arrive to Endoscopy 1.5-2 hrs before the scheduled procedure? | YES | NO |
| **5.** Have you been recording your diet in the food diary? | YES | NO |

***FMT will not be performed if answered "no".**

RECIPIENT NOTES:

**APPENDIX I:**

### Mayo Score

DAY OF TRANSPLANT___

RECIPIENT CODED No.: ____________________

| Item | Points |
|---|---|
| **1. Stool Frequency** | |
| Normal number of stools for the patient | 0 |
| 1-2 stools more than normal | 1 |
| 3-4 stools more than normal | 2 |
| 5 or more stools more than normal | 3 |
| **2. Rectal bleeding** | |
| None | 0 |
| Streaks of blood with stool less than half of the time | 1 |
| Obvious blood with stool most of the time | 2 |
| Blood alone passed | 3 |
| **3. Endoscopic Findings (Rectosigmoid)** | |
| Normal or inactive disease | 0 |
| Mild disease (erythema, decreased vascular pattern, mild friability) | 1 |
| Moderate disease (marked erythema, absent vascular pattern, friability, erosions) | 2 |
| Severe disease (spontaneous bleeding, ulceration) | 3 |
| **4. Physician's Global Assessment** | |
| Normal | 0 |
| Mild disease | 1 |
| Moderate disease | 2 |
| Severe disease | 3 |
| **Total Score** | _________ |

**APPENDIX J:**

### DIARY CARD

DATE:__________________

DESCRIPTION OF SYMPTOMS THAT ARE NEW OR WORSENED:

DATE:__________________

DESCRIPTION OF SYMPTOMS THAT ARE NEW OR WORSENED:

DATE:__________________

DESCRIPTION OF SYMPTOMS THAT ARE NEW OR WORSENED:

DATE:__________________

DESCRIPTION OF SYMPTOMS THAT ARE NEW OR WORSENED:

**Contact us (631-444-7225) immediately if you are seen in the emergency room or hospitalized <u>for any reason.</u>**

**APPENDIX K:**

### DAY AFTER TRANSPLANT: CASE REPORT FORM

RECIPIENT CODED No.: ___________________
DATE/TIME OF CALL: ___ ___/___ ___/___ ___ ___:___AM/PM

RECIPIENT DATA:

| | | | |
|---|---|---|---|
| **1.** How do you feel today? | WELL | | **UNWELL** |
| **2.** Have you had a fever > 100.4 degrees Fahrenheit in the past 2 weeks? | **YES** | | NO |
| **3.** Is your abdominal pain: | BETTER | **WORSE** | SAME |
| **4.** Is your diarrhea: | BETTER | **WORSE** | SAME |
| **5.** Is the blood in your stool: | BETTER | **WORSE** | SAME |
| **6.** Is there a presence of a new rash on your body? | **YES** | | NO |

DESCRIPTION OF SYMPTOMS THAT HAVE WORSENED:

NAME OF PHYSICIAN INFORMED IF PATIENT RESPONSE CORRESPONDS TO ANY OF THE BOLDED ITEMS ABOVE:

________________________MD/DO

PHYSICIAN DECISION OF FURTHER EVALUATION:

| | | |
|---|---|---|
| CLINIC | EMERGENCY ROOM | PHONE FOLLOW-UP |

**APPENDIX L1 (*Clostridium difficile with or without IBD*):**

### FOLLOW UP PHONE CALL/ADVERSE EVENT: CASE REPORT FORM

#### WEEK(S)/MONTH(S) POST-TRANSPLANT ___

RECIPIENT CODED No.: ___________________

DATE/TIME OF CALL: ___ ___/___ ___/___ ___ ___:___AM/PM

| | | | | |
|---|---|---|---|---|
| **1.** How do you feel today? | WELL | | **UNWELL** | |
| **2.** Have you had a fever > 100.4 degrees Fahrenheit since the last phone call? | **YES** | | NO | |
| **3.** Has your abdominal pain: | RESOLVED | IMPROVED | **WORSENED** | N/A |
| **4.** Has your diarrhea: | RESOLVED | IMPROVED | **WORSENED** | N/A |
| **5a.** Frequency of stools/day: | <3 | 3-6 | **>6** | |
| **5b.** Has the blood in your stool: | RESOLVED | IMPROVED | **WORSENED** | N/A |
| **6.** Have you been fatigued? | **YES** | | | NO |
| | **Mild** | **Moderate** | **Severe** | |
| **7.** Have you experienced weight changes since the last phone call? | NO | **YES** | ± ____ LBS | |
| **8.** Have you required antibiotics since the fecal transplant? If yes, what is the name of the antibiotic(s)? | YES<br>Name of antibiotic(s): _______________ | | NO | |
| **9.** Have you taken any new medication(s), including OTC or probiotics since the last phone call? | YES<br>Name of medication(s): _______________ | | NO | |
| **10.** Have you developed any NEW medical conditions since the last phone call? | **YES**<br>**Please specify: _______________** | | NO | |
| **11a.** Did any medical condition(s) you had before your fecal transplant go away after your fecal transplant? | YES<br>Please specify: _______________ | | NO | |
| **11b.** Please list all medication(s) you take on a regular basis (dosage not necessary) | Name of medication(s): ________________________________________ | | | |

| |
|---|
| including chemotherapeutic agents, if applicable |

#### DESCRIPTION OF ADVERSE EVENT(S):

(An adverse event will be defined as any unfavorable or unintended sign, symptoms, disease, syndrome, abnormal laboratory finding, or concurrent illness that emerges or worsens relative to the recipient's pre-transplant baseline, whether or not it is considered to be related to the fecal transplantation)
All Grade 3 – 5 adverse events will be sent to the Safety Monitoring Committee

#### NAME OF PHYSICIAN INFORMED IF PATIENT RESPONSE CORRESPONDS TO ANY OF THE BOLDED ITEMS ABOVE:

________________________MD/DO

#### PHYSICIAN DECISION OF FURTHER EVALUATION:

| | | |
|---|---|---|
| CLINIC | EMERGENCY ROOM | PHONE FOLLOW-UP |

| | | |
|---|---|---|
| FOLLOW UP *C. DIFFICILE* TESTING<br>DATE OF TESTING: __ __/__ __/__ __ | POSITIVE | NEGATIVE |

#### DOCUMENTATION SENT TO SAFETY MONITORING COMMITTEE ON ___ ___/___ ___/___ ___:

| | |
|---|---|
| YES | NO |

**APPENDIX L2 (Ulcerative Colitis or Indeterminate Colitis):**

### FOLLOW UP PHONE CALL/ADVERSE EVENT: CASE REPORT FORM

WEEK(S)/MONTH(S) POST-TRANSPLANT ___

RECIPIENT CODED No.: ___________________

DATE/TIME OF CALL: ___ ___/___ ___/___ ___ ___:___AM/PM

| | | | | |
|---|---|---|---|---|
| **1.** How do you feel today? | WELL | | **UNWELL** | |
| **2.** Have you had a fever > 100.4 degrees Fahrenheit since the last phone call? | **YES** | | NO | |
| **3.** Has your abdominal pain: | RESOLVED | IMPROVED | **WORSENED** | N/A |
| **4.** Has your diarrhea: | RESOLVED | IMPROVED | **WORSENED** | N/A |
| **5a.** Frequency of stools/day: | <3 | 3-6 | **>6** | |
| **5b.** Has the blood in your stool: | RESOLVED | IMPROVED | **WORSENED** | N/A |
| **6.** Have you been fatigued? | **YES**<br>**Mild** | **YES**<br>**Moderate** | **YES**<br>**Severe** | NO |
| **7.** Have you experienced weight changes since the last phone call? | NO | YES | ± ____ LBS | |
| **8.** Have you required antibiotics since the fecal transplant? If yes, what is the name of the antibiotic(s)? | YES<br>Name of antibiotic(s):<br>_______________ | | NO | |
| **9.** Have you taken any new medication(s), including OTC or probiotics since the last phone call? | YES<br>Name of medication(s):<br>_______________ | | NO | |
| **10.** Have you developed any NEW medical conditions since the last phone call? | **YES**<br>**Please specify:**<br>_______________ | | NO | |
| **11a.** Did any medical condition(s) you had before your fecal transplant go away after your fecal transplant? | YES<br>Please specify:<br>_______________ | | NO | |
| **11b.** Please list all medication(s) you take on a regular basis (dosage not necessary) | Name of medication(s):<br>________________________________________ | | | |

| |
|---|
| including chemotherapeutic agents, if applicable |

DESCRIPTION OF ADVERSE EVENT(S):

(An adverse event will be defined as any unfavorable or unintended sign, symptoms, disea1se, syndrome, abnormal laboratory finding, or concurrent illness that emerges or worsens relative to the recipient's pre-transplant baseline, whether or not it is considered to be related to the fecal transplantation)

All Grade 3 – 5 adverse events will be sent to the Safety Monitoring Committee.

NAME OF PHYSICIAN INFORMED IF PATIENT RESPONSE CORRESPONDS TO ANY OF THE BOLDED ITEMS ABOVE:

_______________________MD/DO

PHYSICIAN DECISION OF FURTHER EVALUATION:

| | | |
|---|---|---|
| CLINIC | EMERGENCY ROOM | PHONE FOLLOW-UP |

DOCUMENTATION SENT TO SAFETY MONITORING COMMITTEE ON ___ ___/___ ___/___ ___:

| | |
|---|---|
| YES | NO |

**APPENDIX M:**

### SERIOUS ADVERSE EVENT (SAE): CASE REPORT FORM

WEEK(S)/MONTH(S) POST-TRANSPLANT ___

☐ INITIAL REPORT/☐ FOLLOW-UP REPORT

RECIPIENT CODED No.: __________________
ONSET OF EVENT: ___ ___/___ ___/___ ___

| Severity of SAE | Grade 3 | Grade 4 | Grade 5 |
|---|---|---|---|

| | | |
|---|---|---|
| Outcome of SAE: | ☐ RECOVERED WITH SEQUELAE | RECOVERY DATE ___ ___/___ ___/___ ___ |
| | ☐ RECOVERED WITHOUT SEQUELAE | |
| | ☐ PERSISTING | ☐ UNKNOWN/LOST TO FOLLOW-UP |
| | ☐ DEATH | |

| | | |
|---|---|---|
| SAE category: | ☐ DEATH* | ☐ LIFE THREATENING |
| | ☐ HOSPITALIZATION REQUIRED# | ☐ PROLONGED IN-PATIENT HOSPITALIZATION# |
| | ☐ PERSISTENT/SIGNIFICANT DISABILITY OR INCAPACITY | ☐ OTHER MEDICALLY IMPORTANT CONDITION |

#### *IF DEATH HAS OCCURRED:

| | |
|---|---|
| **1.** Date of death | ____ ____/____ ____/____ ____ |
| **2.** Primary cause of death (if known) | ______________________________ |

#### #IF RECIPIENT HAS REQUIRED HOSPITALIZATION:

| | |
|---|---|
| **1.** Hospital admission date | ____ ____/____ ____/____ ____ |
| **2.** Current duration of hospitalization | ____________ (WEEKS/MONTHS) |

#### DESCRIPTION OF SERIOUS ADVERSE EVENT(S):

NAME OF PHYSICIAN INFORMED OF SAE: ________________________MD/DO

DOCUMENTATION SENT TO SAFETY MONITORING COMMITTEE ON ___ ___/___ ___/___ ___:

| YES | NO |
|---|---|

Approved: 6/10/2021
Expiration Date: 6/9/2022
Stony Brook University Human Subjects Committee (IRB)

**APPENDIX N**

### Research Study: Fecal microbial transplantation in patients with medication-refractory *Clostridium difficile* and/or Ulcerative colitis or indeterminate colitis (IRBNet #: 479696)

#### Principal Investigator: Dr. Anupama Chawla

FMT recipient ______________________________

FMT donor ______________________________

Dr. Anupama Chawla has discussed the screening test for the Donor with both recipient and donor as named above and explained that the costs involved in these tests might not be covered by the insurance. The research study does not provide any monetary support for these donor-screening tests.

Signature of Recipient____________________________

Signature of Donor ____________________________

_______________________________________________

Full name of person obtaining consent

_______________________________________________

Signature of person obtaining consent Date

**APPENDIX O**

### Checklist for instructing Donor 2-5 days prior to Fecal Microbial Transplantation

**DONOR CODED No.: ___________________________________**
**DATE/TIME OF CALL: ___ ___/___ ___/___ ___:___AM/PM**

DONOR DATA:

| | | |
|---|---|---|
| **1.** Do you fully understand the stool collection instructions & kit? | YES | NO |
| **2.** Have you been recording your diet in the food diary? | YES | NO |
| **3.** Have you ingested any known allergy/sensitivity of the recipient?* If so, see notes below | YES | NO |
| **4.** Will you be present with the recipient on day of transplant? (If no, complete TPF questionnaire over phone and schedule a time to complete "Day Of Transplant Case Report" form) | YES | NO |

DONOR NOTES:

***Reschedule fecal microbial transplant.**

**APPENDIX P:**

### DAY OF TRANSPLANT: DONOR CASE REPORT FORM

DATE OF STUDY: ___ ___/___ ___/___ ___ TIME OF STUDY: ___:___

DONOR CODED No.: ____________________________________

RECIPIENT CODED No.: _________________________________

DONOR DATA:

| | | |
|---|---|---|
| **1.** Do you feel unwell today? | YES | NO |
| **2.** Have you had a fever > 100.4 degrees Fahrenheit in the past 2 weeks? | YES | NO |
| **3.** Have you had cough or runny nose within the last 2 weeks? | YES | NO |
| **4.** Have you ingested ________ (recipient allergen) in the past week? | YES | NO |

**If answer to any question is "yes", discuss with attending physician.**

DONOR STOOL DATA:

| | | | |
|---|---|---|---|
| **1.** What date and time was the stool produced? | __ __/__ __/__ __ AM/PM | | |
| **2.** If produced > 6 hours before time of colonoscopy, was the stool kept at room temperature? | YES | NO | N/A |
| **3.** Was the stool collected with any contamination with the toilet bowl? | YES | NO | |
| **4.** Is there presence of blood in the stool? | YES | NO | |
| **5.** Is there presence of mucous in the stool? | YES | NO | |
| **6.** What is the consistency of stool? | Hard | Soft | Liquidy |

**If answer to any question from #2 to #5 is "yes", FMT will not be performed.**

Approved: 6/10/2021
Expiration Date: 6/9/2022
Stony Brook University Human Subjects Committee (IRB)

**APPENDIX Q:**

### Pediatric Ulcerative Colitis Activity Index

WEEK(S)/MONTH(S) POST-TRANSPLANT ___

RECIPIENT CODED No.: ____________________

| Item | Points |
|---|---|
| **1. Abdominal pain** | |
| No pain | 0 |
| Pain can be ignored | 5 |
| Pain cannot be ignored | 10 |
| **2. Rectal bleeding** | |
| None | 0 |
| Small amount only, in <50% of stools | 10 |
| Small amount with most stools | 20 |
| Large amount (>50% of stool content) | 30 |
| **3. Stool consistency of most stools** | |
| Formed | 0 |
| Partially formed | 5 |
| Completely formed | 10 |
| **4. Number of stools per 24 hours** | |
| 0-2 | 0 |
| 3-5 | 5 |
| 6-8 | 10 |
| >8 | 15 |
| **5. Nocturnal stools (any episode causing wakening)** | |
| No | 0 |
| Yes | 10 |
| **6. Activity level** | |
| No limitation of activity | 0 |
| Occasional limitation of activity | 5 |
| Severe restricted activity | 10 |
| **Total Score** | ________ |
