## Supplemental Table S2 for "Integrated metagenomic and bile acid metabolomic analysis of human fecal microbiota transplantation for recurrent Clostridioides difficile and/or inflammatory bowel diseases"

**S2 Table Chromatographic and quantitation strategy for the BA and internal standards (ISTD) measured.** see Methods.

| **BA** | **Retention time or RT (min)** | **Assigned ISTD** | **ISTD RT (min)** | **Method** | **Quantitation strategy** |
|---|---|---|---|---|---|
| **Primary conjugated** | | | | | |
| T-CA | 12.727 | T-CA-d4 | 12.706 | Method 2 | Single point |
| G-CDCA | 10.479 | G-CDCA-d4 | 14.479 | Method 2 | Single point |
| G-CA | 10.591 | G-CA-d4 | 10.591 | Method 2 | Single point |
| **Primary unconjugated** | **RT (min)** | **Assigned ISTD** | **ISTD RT (min)** | **Method** | **Quantitation strategy** |
| CDCA | 10.408 | CDCA-d4 | 10.399 | Method 1 | Single-point |
| CA | 8.237 | CA-d4 | 8.236 | Method 1 | Single point |
| CDCA-3-sulfate | 7.700 | CDCA-d4 | 10.399 | Method 1 | Single point |
| CA-3-sulfate | 5.755 | βMCA-d5 | 7.070 | Method 1 | External CC |
| **Secondary conjugated** | **RT (min)** | **Assigned ISTD** | **ISTD RT (min)** | **Method** | **Quantitation strategy** |
| G-DCA | 15.197 | G-DCA-d4 | 15.064 | Method 2 | Single-point |
| **Secondary unconjugated** | **RT (min)** | **Assigned ISTD** | **ISTD RT (min)** | **Method** | **Quantitation strategy** |
| DCA | 10.673 | DCA-d4 | 10.660 | Method 1 | Single-point |
| LCA | 12.627 | LCA-d4 | 12.611 | Method 1 | Single-point |
| UCDA | 8.242 | UDCA-d4 | 8.223 | Method 1 | Single-point |
| iso-CDCA | 9.220 | CDCA-d4 | 10.399 | Method 1 | Single-point |
| iso-DCA | 9.308 | CDCA-d4 | 10.399 | Method 1 | Single-point |
| iso-LCA | 11.951 | LCA-d4 | 12.611 | Method 1 | Single-point |
| isoallo-LCA | 11.600 | LCA-d4 | 12.611 | Method 1 | Single-point |
| 3-oxo-LCA | 12.925 | LCA-d4 | 12.611 | Method 1 | External CC |
| ursocholic acid (UCA) | 9.305 | βMCA-d5 | 13.409 | Method 2 | Single-point |
| 3-oxo-CDCA | 18.750 | CDCDA-d4 | 18.276 | Method 2 | Single-point |
| DCA-3-sulfate | 7.900 | CDCA-d4 | 10.399 | Method 1 | External |
| LCA-3-sulfate | 9.451 | LCA-d4 | 12.611 | Method 1 | External |
| UCDA-3-sulfate | 5.423 | CDCA-d4 | 10.399 | Method 1 | External |
