## Supplemental Table S3 for "Integrated metagenomic and bile acid metabolomic analysis of human fecal microbiota transplantation for recurrent Clostridioides difficile and/or inflammatory bowel diseases"

**S3 Table Bile acid metabolizing enzyme genes used in DIAMOND custom search.** See Methods.

| Gene/ phylotype | Taxa | UniPro/ Genbank |
|---|---|---|
| BaiE | *Clostridium scindens* | P19412.1 |
| BaiCD | *Clostridium scindens* | P19410.2 |
| 3α-HSDH | *Ruminococcus gnavus* | A7B3K3 |
| 3α-HSDH | *Eggerthella lenta* | C8WMP0 |
| 3β-HSDH | *Ruminococcus gnavus* | AZAZH2 |
| 3β-HSDH | *Eggerthella lenta* | A7B3K3 |
| 3β-HSDH | *Bacteroides dorei* | B6WON1 |
| 5β-reductase | *Bacteroides dorei* | B6WON2 |
| 5α- reductase | *Bacteroides dorei* | B6WON3 |
| BSH-T0 | *Lactobacillus salivarius* | WP_204770828.1 |
| BSH-T0 | *Enterococcus* | WP_002287105.1 |
| BSH-T0 | *Clostridium butyricum* | WP_191023471.1 |
| BSH-T0 | *Intestinibacter bartletti* | WP_147617098.1 |
| BSH-T1 | *Ruminococcus gnavus* | WP_173900661.1 |
| BSH-T1 | *Bacteroides pectinophilus* | EEC56367.1 |
| BSH-T1 | *Faecalibacterium prausnitzii* | WP_154265717.1 |
| BSH-T1 | *Catenibacterium mitsuokai* | WP_195921032.1 |
| BSH-T2 | *Streptococcus equinus* | WP_204983623.1 |
| BSH-T2 | *Ligilactobacillus ruminus* | EFZ35634.1 |
| BSH-T2 | *Listeria monocytogenes* | WP_205272850.1 |
| BSH-T2 | *Enterococcus faecalis* | WP_194178372.1 |
| BSH-T3 | *Lactobacillus crispatus* | WP_204777261.1 |
| BSH-T3 | *Lactobacillus ultunensis* | WP_201247470.1 |
| BSH-T4 | *Bifidobacterium animalis* | WP_130079430.1 |
| BSH-T4 | *Bifidobacterium gallicum* | WP_006294150.1 |
| BSH-T4 | *Bifidobacterium longum* | WP_212105292.1 |
| BSH-T4 | *Collinsella aerofaciens* | WP_195919494.1 |
| BSH-T5 | *Fusobacterium mortiferum* | EEO35279.1 |
| BSH-T5 | *Bacteroides ovatus* | EDO11390.1 |
| BSH-T5 | *Bacterooides intestinalis* | EDV03500.1 |
| BSH-T5 | *Parabacteroides distasonis* | QCY55526.1 |
| BSH-T6 | *Bateroides thetaiotaomicron (2086)* | 6UH4_D (PDB) |
| BSH-T6 | *Bacteroides eggerthii* | WP_138350149.1 |
| BSH-T6 | *Bacteroides fragilis* | KXU42764.1 |
| BSH-T6 | *Bacteroides caccae* | WP_149928375.1 |
| BSH-T6 | *Parabacteroides johnsonii* | WP_008158507.1 |
| BSH-T7 | *Anaerotruncus* | WP_120483948.1 |
| BSH-T7 | *Anaerostipes* | WP_212385494.1 |
| BSH-T7 | *Marvinbryantia* | WP_006862905.1 |
| BSH-T7 | *Blautia obeum* | WP_015542091.1 |
