## Supplemental Table S4 for "Integrated metagenomic and bile acid metabolomic analysis of human fecal microbiota transplantation for recurrent Clostridioides difficile and/or inflammatory bowel diseases"

**S4 Table. Individual characteristics of the rCDI – IBD recipients.** The fecal calprotectin level is shown at 1 week post-FMT. The arrow (↓) indicates a decrease and the arrow (↑) indicates an increase in the level compared to the pre-FMT fecal calprotectin level. (*) indicate recipients that developed rCDI within a year post-FMT. (‡) indicates recipients that reported a SAE within a year post-FMT.

| Subject | Prior surgery | 1 week post FMT fecal calprotectin *µg/g* |
|---|---|---|
| **FMT-4R‡** | none | 289 (↓88) |
| **FMT-5R*‡** | s/p LAR + diversion colostomy | 1250 (↑1080) |
| **FMT-9R** | none | 0 (no Δ) |
| **FMT-10R** | none | 20 (↑20) |
| **FMT-12R** | none | 0 (↓32) |
| **FMT-13R** | none | 84 (↑20) |
| **FMT-14R** | none | 31 (↑6) |
| **FMT-15R** | none | 55 (↑21) |
| **FMT-17R** | none | 138 (↑86) |
| **FMT-20R** | none | 31 (↓33) |
| **FMT-21R** | none | 118 (↓76) |
| **FMT-23R** | none | 24 (↑24) |
| **FMT-24R** | none | 55 (↑15) |
| **FMT-26R** | none | 41 (↑35) |
| **FMT-27R** | none | 0 (no Δ) |
| **FMT-29R** | none | 75 (↓15) |
| **FMT-30R** | none | 103 (↑26) |
| **FMT-32R** | none | 169 (↑4) |
| **FMT-33R*** | none | 61 (↑61) |
| **FMT-34R** | none | 0 (no Δ) |
| **FMT-35R‡** | none | 0 (no Δ) |
| **FMT-37R** | none | 17 (↑17) |
| **FMT-39R** | none | missing (missing) |
| **FMT-41R** | none | 34 (↑12) |
| **FMT-44R** | none | 56 (↓1) |
| **FMT-47R** | none | 76 (↓7) |
| **FMT-50R** | none | missing (missing) |
