## Supplemental Table S5 for "Integrated metagenomic and bile acid metabolomic analysis of human fecal microbiota transplantation for recurrent Clostridioides difficile and/or inflammatory bowel diseases"

**S5 Table. Individual characteristics of the rCDI + IBD recipients.** The fecal calprotectin level is shown at 1 week post-FMT. The arrow (↓) indicates a decrease and the arrow (↑) indicates an increase in the level compared to the pre-FMT fecal calprotectin level. (*) indicate recipients that developed rCDI within a year post-FMT. (‡) indicates recipients that reported a SAE within a year post-FMT.

| Subject | IBD phenotype (Montreal classification) | Prior surgery | 1 week post FMT fecal calprotectin µg/g | 3 month post FMT Δ IBD Rx |
|---|---|---|---|---|
| **FMT-2R*** | left sided UC (E2) | none | 859 (↑759) | none |
| **FMT-6R‡** | ileal CD (L1) | s/p ICR | 221 (↓53) | perianal surgery |
| **FMT-11R** | extensive UC (E3) | none | 1250 (↑590) | Δ biologic |
| **FMT-18R** | left sided UC (E2) | none | 443 (↑360) | add biologic |
| **FMT-25R** | extensive UC (E3) | none | 250 (↓879) | Δ biologic |
| **FMT-28R** | left sided UC (E2) | none | 83 (↓16) | none |
| **FMT-31R** | ileocolonic CD (L3) | s/p ICR | 118 (↓114) | none |
| **FMT-42R*** | ileal CD (L1) | none | 31 (↓49) | add biologic |
