## Supplemental Table S6 for "Integrated metagenomic and bile acid metabolomic analysis of human fecal microbiota transplantation for recurrent Clostridioides difficile and/or inflammatory bowel diseases"

**S6 Table. Individual characteristics of the UC - rCDI recipients.** The fecal calprotectin level is shown at 1 week post-FMT. The arrow (↓) indicates a decrease and the arrow (↑) indicates an increase in the level compared to the pre-FMT fecal calprotectin level. 6-mercaptopurine (6-MP) was discontinued in one patient. (‡) indicates recipients that reported a SAE within a year post-FMT.

| **Subject** | **IBD phenotype (Montreal classification)** | **Prior surgery** | **Hx CDI** | **Mayo endo score** | **1 week post FMT fecal calprotectin *µg/g*** | **3 month post FMT Δ IBD Rx** |
|---|---|---|---|---|---|---|
| **FMT-1R** | extensive UC (E3) | none | no | 0 | 46 (↓43) | d/c steroids |
| **FMT-3R** | extensive UC (E3) | none | no | 2 | 307 (↑136) | none |
| **FMT-7R** | extensive UC (E3) | none | no | 2 | 277 (↓307) | add biologic |
| **FMT-8R** | extensive UC (E3) | none | no | 3 | 176 (↓1076) | none |
| **FMT-19R** | extensive UC (E3) | none | no | 2 | 1250 (↑545) | none |
| **FMT-38R** | extensive UC (E3) | none | no | 0 | 0 (no Δ) | none |
| **FMT-43R** | extensive UC (E3) | none | no | 1 | 96 (↓11) | none |
| **FMT-45R‡** | left sided UC (E2) | none | no | 2 | 157 (↑14) | colectomy |
| **FMT-46R** | extensive UC (E3) | none | yes | 2 | 1250 (no Δ) | none |
| **FMT-48R** | proctitis (E1) | none | no | 0 | 0 (↑16) | d/c 6-MP |
| **FMT-49R** | left sided UC (E2) | none | no | 3 | 3 (↓350) | Δ biologic |
