## Supplemental Table S7 for "Integrated metagenomic and bile acid metabolomic analysis of human fecal microbiota transplantation for recurrent Clostridioides difficile and/or inflammatory bowel diseases"

**S7 Table. Individual characteristics of the CD - rCDI recipients.** The fecal calprotectin level is shown at 1 week post-FMT. The arrow (↓) indicates a decrease and the arrow (↑) indicates an increase in the level compared to the pre-FMT fecal calprotectin level. (‡) indicates recipients that reported a SAE within a year post-FMT.

| Subject | IBD phenotype (Montreal classification) | Surgery | Hx CDI | SES-CD score | 1 week post FMT fecal calprotectin *μg/g* | 3 month post FMT Δ IBD Rx |
|---|---|---|---|---|---|---|
| **CD1-R** | colonic + UGI CD (L2 + L4) | none | no | 1 | 188 (↓408) | none |
| **CD2-R** | colonic + UGI CD (L2 + L4) | none | no | 0 | 927 (↑927) | Δ biologic |
| **CD3-R** | colonic + perianal CD(L2+p) | none | no | 7 | 523 (↓434) | + steroids |
| **CD4-R** | colonic+ UGI CD (L2 + L4) | none | no | 14 | 1250 (↑1112) | none |
| **CD5-R** | colonic + UGI CD (L2 + L4) | none | no | 14 | 952 (↑923) | +ASA steroids |
| **CD6-R‡** | ileocolonic + perianal CD (L3+p) | none | no | 16 | 1250 (↑864) | + steroids |
| **CD7-R** | ileocolonic + perianal CD (L3+p) | s/p ICR | no | 14 | 690 (↓560) | Δ biologic |
| **CD8-R** | colonic CD (L2) | none | no | 0 | 9 (↑9) | none |
| **CD9-R** | ileal CD (L1) | s/p ICR x 2 | no | 8 | 165 (missing) | Δ biologic |
