## Supplemental Table S8 for "Integrated metagenomic and bile acid metabolomic analysis of human fecal microbiota transplantation for recurrent Clostridioides difficile and/or inflammatory bowel diseases"

**S8 Table. Significant (FDR < 0.05) estimated pre-FMT recipient/donor ratios (95% CI) of the relative abundances of Actinobacteria genus level OTUs for each of the four recipient groups.** A ratio > 1 indicates ↑ relative abundance in recipient pre-FMT compared to donor samples. A ratio of < 1 indicated ↓ relative abundance in pre-FMT samples compared to donor samples. The 95% CI are shown in parenthesis. Only the estimated ratios with an FDR < 0.05 are listed. The * indicates those OTUs that also had significant estimated ratios of post-FMT/pre-FMT relative abundances at with 1 week and/ or 3 month time point, indicating that FMT had a significant effect.

| **Actinobacteria OTU** | **rCDI - IBD** | **rCDI + IBD** | **UC - rCDI** | **CD - rCDI** |
|---|---|---|---|---|
| *Actinomyces* | | | | 4.15* (1.71,10.04) |
| *Bifidobacterium* | 6.64 (1.98,22.29) | | | |
| *Coriobacteriaceae/ Unspecified* | 0.01* (0.004,0.04) | 0.02* (0.002,0.146) | | |
| *Collinsella* | 0.04* (0.006,0.22) | 0.001* (0,0.002) | | 0.34* (0.18,0.62) |
| *Gordonibacter* | | 0.02* (0.01,0.03) | | 0.26 (0.09,0.79) |
