## Supplemental Table S9 for "Integrated metagenomic and bile acid metabolomic analysis of human fecal microbiota transplantation for recurrent Clostridioides difficile and/or inflammatory bowel diseases"

**S9 Table. Significant (FDR < 0.05) estimated pre-FMT recipient/donor ratios (95% CI) of the relative abundances of Bacteroidetes genus level OTUs for each of the four recipient groups.** A ratio > 1 indicates ↑ relative abundance in recipient pre-FMT compared to donor samples. A ratio of < 1 indicated ↓ relative abundance in pre-FMT samples compared to donor samples. The 95% CI are shown in parenthesis. Only the estimated ratios with an FDR < 0.05 are listed. The * indicates those OTUs that also had significant estimated ratios of post-FMT/pre-FMT relative abundances at with 1 week and/ or 3 month time point, indicating that FMT had a significant effect.

| Bacteroidetes OTU | rCDI - IBD | rCDI + IBD | UC - rCDI | CD - rCDI |
|---|---|---|---|---|
| *Unassigned* | 0.01* (0.002,0.05) | 0.06* (0.03,0.14) | 0.07 (0.008,0.52) | 0.20* (0.11,0.37) |
| *Bacteroides* | 0.15* (0.06,0.37) | 0.08 (0.01,0.63) | | |
| *Barnesiella* | | 0.002* (0.001,0.01) | 4.91* (1.58,15.27) | |
| *Butyricimonas* | 0.07* (0.02,0.27) | 0.15* (0.04,0.65) | | |
| *Odoribacter* | 0.0014* (0.0006,0.003) | 0.0001* (0.00001,0.001) | | |
| *Parabacteroides* | | 0.002* (0.001,0.003) | | |
| *Prevotella* | | 0.03* (0.005,0.19) | | |
| *Alistipes* | 0.002* (0.001,0.005) | 0.0005* (0.0001,0.002) | | 0.13 (0.03,0.48) |
| *S24-7* | 0.20* (0.05,0.75) | | | 0.24* (0.09,0.64) |
| *VC2.1-Bac22* | 0.01* (0.001,0.10) | 0.03* (0.005,0.17) | | |
