## Supplemental Table S10 for "Integrated metagenomic and bile acid metabolomic analysis of human fecal microbiota transplantation for recurrent Clostridioides difficile and/or inflammatory bowel diseases"

**S10 Table. Significant (FDR < 0.05) estimated pre-FMT recipient/donor ratios (95% CI) of the relative abundances of Ruminococcaceae (Clostridium Group IV) genus level OTUs for each of the four recipient groups.** A ratio > 1 indicates ↑ relative abundance in recipient pre-FMT compared to donor samples. A ratio of < 1 indicated ↓ relative abundance in pre-FMT samples compared to donor samples. The 95% CI are shown in parenthesis. Only the estimated ratios with an FDR < 0.05 are listed. The * indicates those OTUs that also had significant estimated ratios of post-FMT/pre-FMT relative abundances at with 1 week and/ or 3 month time point, indicating that FMT had a significant effect.

| Ruminococcaceae OTU | rCDI - IBD | rCDI + IBD | UC - rCDI | CD - rCDI |
|---|---|---|---|---|
| *Unspecified* | 0.23* (0.11,0.47) | 0.07* (0.01,0.37) | | 0.28* (0.18,0.42) |
| *Anaerofilum* | 0.15* (0.06,0.36) | 0.37* (0.16,0.84) | | 0.43 (0.24,0.76) |
| *Anaerotruncus* | | | | 0.12 (0.03,0.47) |
| *Faecalibacterium* | 0.004* (0.002,0.007) | 0.003* (0.002,0.004) | | |
| *Ocillospira* | 0.04* (0.01,0.13) | | | |
| *Ruminococcus* | 0.02* (0.003,0.14) | 0.02* (0.003,0.07) | 0.37 (0.23,0.61) | |
| *Subdoligranum* | 0.01* (0.002,0.07) | 0.003* (0.002,0.005) | | 0.07* (0.02,0.28) |
