## Supplemental Table S11 for "Integrated metagenomic and bile acid metabolomic analysis of human fecal microbiota transplantation for recurrent Clostridioides difficile and/or inflammatory bowel diseases"

**S11 Table. Significant (FDR < 0.05) estimated pre-FMT recipient/donor ratios (95% CI) of the relative abundances of Lachnospiraceae (Clostridium Group XIVa) genus level OTUs for each of the four recipient groups.** A ratio > 1 indicates ↑ relative abundance in recipient pre-FMT compared to donor samples. A ratio of < 1 indicated ↓ relative abundance in pre-FMT samples compared to donor samples. The 95% CI are shown in parenthesis. Only the estimated ratios with an FDR < 0.05 are listed. The * indicates those OTUs that also had significant estimated ratios of post-FMT/pre-FMT relative abundances at with 1 week and/ or 3 month time point, indicating that FMT had a significant effect.

| **Lachnospiraceae OTU** | **rCDI - IBD** | **rCDI + IBD** | **UC - rCDI** | **CD - rCDI** |
|---|---|---|---|---|
| *Anaeorostipes* | 0.24* (0.08,0.74) | | | |
| *Blautia* | 0.44* (0.25,0.78) | | | |
| *Coprococcus* | | 0.002* (0.001,0.004) | | |
| *Dorea* | 0.003* (0.001,0.008) | 0.003* (0.001,0.005) | | |
| *Lachnospira* | | 0.113 (0.024,0.527) | | |
| *Marvinbryantia* | 0.09* (0.02,0.47) | 0.02* (0.001,0.22) | | |
| *Pseudobutyrivibrio* | 0.03* (0.01,0.16) | | | |
| *Roseburia* | | 0.17 (0.05,0.61) | | |
