## Supplemental Table S12 for "Integrated metagenomic and bile acid metabolomic analysis of human fecal microbiota transplantation for recurrent Clostridioides difficile and/or inflammatory bowel diseases"

**S12 Table. Significant (FDR < 0.05) estimated pre-FMT recipient/donor ratios (95% CI) of the relative abundances of Bacilli genus level OTUs for each of the four recipient groups.** A ratio > 1 indicates ↑ relative abundance in recipient pre-FMT compared to donor samples. A ratio of < 1 indicated ↓ relative abundance in pre-FMT samples compared to donor samples. The 95% CI are shown in parenthesis. Only the estimated ratios with an FDR < 0.05 are listed. The * indicates those OTUs that also had significant estimated ratios of post-FMT/pre-FMT relative abundances at with 1 week and/ or 3 month time point, indicating that FMT had a significant effect.

| **Bacilli OTU** | **rCDI - IBD** | **rCDI + IBD** | **UC - rCDI** | **CD - rCDI** |
|---|---|---|---|---|
| *Unspecified* | 138.75* (58.52,328.99) | 44.62* (12.95,153.76) | | |
| *Planomicrobium* | 190.46* (24.01,1511.17) | 120.81* (14.87,981.47) | | |
| *Staphylococcus* | 6.94* (2.37,20.31) | | 13.24* (2.61,67.31) | |
| *Lactobacillales/ Unspecified* | 282.10* (99.02,803.69) | 115.94* (35.08,383.18) | 6.17* (1.84,20.73) | |
| *Enterococcus* | 147.86* (32.49,672.95) | | 39.72 (12.42,127.00) | 17.23* (5.33,55.67) |
| *Lactobacillus* | 151.55* (40.69,564.42) | 891.53* (300.81,2642.33) | 94.61* (29.69,301.42) | |
| *Streptococcacea/ Unspecified* | 13.50* (4.47,40.79) | 9.05 (4.99,16.41) | | |
| *Lactococcus* | 3.42* (1.49,7.84) | | 18.71 (6.94,50.45) | 0.03 (0.004,0.24) |
| *Streptococcus* | | | 7.94 (1.95,32.36) | 2.73* (1.67,4.44) |
