## Supplemental Table S13 for "Integrated metagenomic and bile acid metabolomic analysis of human fecal microbiota transplantation for recurrent Clostridioides difficile and/or inflammatory bowel diseases"

**S13 Table. Significant (FDR < 0.05) estimated pre-FMT recipient/donor ratios (95% CI) of the relative abundances of Proteobacteria genus level OTUs for each of the four recipient groups.** A ratio > 1 indicates ↑ relative abundance in recipient pre-FMT compared to donor samples. A ratio of < 1 indicated ↓ relative abundance in pre-FMT samples compared to donor samples. The 95% CI are shown in parenthesis. Only the estimated ratios with an FDR < 0.05 are listed. The * indicates those OTUs that also had significant estimated ratios of post-FMT/pre-FMT relative abundances at with 1 week and/ or 3 month time point, indicating that FMT had a significant effect.

| Proteobacteria OTU | rCDI - IBD | rCDI + IBD | UC - rCDI | CD - rCDI |
|---|---|---|---|---|
| *Unspecified* | 35.03* (12.71,96.54) | 32.85 (8.35,129.20) | | |
| *Bilophila* | | 0.004* (0.002,0.008) | | 0.037* (0.009,0.15) |
| *Gammaproteobacteria/ Unspecified* | 99.54* (18.80,527.12) | 300.78* (83.24,1086.85) | 7.42* (2.23,24.66) | 26.65 (3.95,179.90) |
| *B38* | 16.86* (6.24,45.52) | 214.04* (23.54,1946.14) | | |
| *Enterobacteriaceae/ Unspecified* | 57.60* (8.71,380.92) | 1724.82* (517.90,5744.37) | 5.01* (1.77,14.21) | |
| *Citrobacter* | 14.20* (3.46, 58.35) | 1022.05* (240.83,4337.51) | | |
| *Enterobacter* | 154.98* (25.78,931.71) | 303.65* (30.73,3000.34) | | 0.00003* (0.000003, 0.00025) |
| *Escherichia-Shigella* | 15.59* (4.29,56.68) | 43.75* (12.06,158.73) | | 76.65 (9.39,625.70) |
| *Klebsiella* | 895.51* (241.57,3319.72) | 505.91* (115.21,2221.56) | 5.85* (2.07,16.55) | |
