## Supplemental Table S14 for "Integrated metagenomic and bile acid metabolomic analysis of human fecal microbiota transplantation for recurrent Clostridioides difficile and/or inflammatory bowel diseases"

**S14 Table. Significant (FDR < 0.05) estimated pre-FMT recipient/donor ratios (95% CI) of the relative abundances of genus level OTUs in "Other Taxa" for each of the four recipient groups.** A ratio > 1 indicates ↑ relative abundance in recipient pre-FMT compared to donor samples. A ratio of < 1 indicated ↓ relative abundance in pre-FMT samples compared to donor samples. The 95% CI are shown in parenthesis. Only the estimated ratios with an FDR < 0.05 are listed. The * indicates those OTUs that also had significant estimated ratios of post-FMT/pre-FMT relative abundances at with 1 week and/ or 3 month time point, indicating that FMT had a significant effect.

| Other Taxa OTU | rCDI - IBD | rCDI + IBD | UC - rCDI | CD - rCDI |
|---|---|---|---|---|
| **Other Firmicutes OTU** | | | | |
| *Christensenellaceae/ Unspecified* | 0.059* (0.011,0.324) | 0.002* (0.001,0.005) | | |
| *Anaerofustis* | | 0.039* (0.006,0.263) | | |
| *Eubacterium* | 0.16* (0.07,0.39) | | | |
| *Anaerococcus* | | | 514.3* (41.7,6343.2) | |
| *Parvimonas* | | | 6.99* (5.38,9.07) | |
| *Peptoniphilus* | | | 9.30 (3.42,25.27) | 3.22* (1.61,6.45) |
| *Family-XIII-Incertae-Sedis/ Unspecified* | | 0.003* (0.001,0.01) | 0.33* (0.15,0.69) | |
| *Peptococcaceae/ Unspecified* | 0.003* (0.001,0.02) | 0.002* (0.0001,0.038) | | |
| *Peptococcus* | 0.03* (0.02,0.06) | 0.26* (0.10,0.63) | | 0.04* (0.01,0.14) |
| *Turicibacter* | | | 5.35* (2.63,10.89) | |
| *Catenibacterium* | 0.03* (0.005,0.13) | 0.004* (0.001,0.013) | | |
| *Acidaminococcus* | 11.53* (2.23, 59.72) | 0.023* (0.001,0.43) | | |
| *Phascolarctobacterium* | | 0.005* (0.001,0.02) | | 0.19* (0.17,0.22) |
| *Veillonellaceae/ Unspecified* | 1706.7* (527.4, 5522.9) | 449.6* (152.3,1327.8) | | |
| *Megamonas* | 0.06* (0.01,0.32) | 0.001* (0.0001,0.009) | 0.002* (0.0001,0.02) | |
| *Megasphaera* | | | 14.49* (2.61,80.33) | 0.57* (0.42,0.77) |
| *Veillonella* | 322.2* (125.3,828.6) | 332.10* (144.7,762.0) | | 34.8 (9.36,129.4) |
| **Fusobacteria OTU** | | | | |
| *Fusobacteriales/ Unspecified* | 286.0* (78.1,1047.7) | 1734.28* (473.9,6347.1) | | |
| *Fusobacterium* | 640.5* (147.2,2786.8) | 3209.9* (875.9,11763) | | 13.51* (1.88,96.8) |
| **Candidate-division-TM7 OTU** | | | | |

| | | | | |
|---|---|---|---|---|
| *Candidate-division-TM7* | | 6.25* (2.18,17.89) | | |
| **Verrucomicrobia OTU** | | | | |
| *Akkermansia* | 6.66* (2.89,15.34) | | | 0.003* (0.0003,0.04) |
| **Cyanobacteria OTU** | | | | |
| *4C0d-2* | | | | 0.07 (0.01,0.40) |
| *Chloroplast* | | | | 0.51 (0.39,0.67) |
| **Bacteria/ Unassigned OTU** | | | | |
| *Unassigned* | 0.29* (0.15,0.53) | 0.09* (0.02,0.41) | | 0.19 (0.09,0.40) |
