## Supplemental Table S15 for "Integrated metagenomic and bile acid metabolomic analysis of human fecal microbiota transplantation for recurrent Clostridioides difficile and/or inflammatory bowel diseases"

**S15 Table. Median Pre-FMT BA levels (*pmol/g* wet weight of stool or *µM*) ± IQR in donor and four recipient groups.**

| BA (*µM*) | Donor n = 55 (2 missing) | rCDI -IBD n = 27 (4 missing) | rCDI + IBD n = 8 | UC – rCDI n = 11 | CD – rCDI n = 9 | P-value |
|---|---|---|---|---|---|---|
| **Primary conjugated** | | | | | | |
| T-CA | 2.5 ± 0.0 | 2.5 ± 295.5 | 810.8 ±1354.5 | 2.5 ± 34.2 | 2.5 ± 36.5 | 0.0095 |
| G-CDCA | 51.3 ± 80.1 | 177.8 ± 230.5 | 1339.9 ± 2322.9 | 45.8 ± 84.7 | 59.8 ± 85.5 | 0.0002 |
| G-CA | 38.8 ± 77.6 | 136.5 ± 362.6 | 1554.6 ± 1797.4 | 31.1 ± 96.8 | 46.0 ± 62.1 | 0.0012 |
| **Primary unconjugated** | **Donor** | **rCDI - IBD** | **rCDI + IBD** | **UC – rCDI** | **CD – rCDI** | **P-value** |
| CDCA | 140.1 ± 803.0 | 2802.2 ± 7396.5 | 1813.4 ± 2791.5 | 124.4 ± 258.7 | 2163.1 ± 5023.2 | 0.002 |
| CA | 74.3 ± 509.8 | 6194.4 ± 6964.8 | 3242.1 ± 5514.4 | 211.7 ± 395.0 | 4310.7 ± 4189.2 | <.0001 |
| CDCA-3-sulfate | 29.3 ± 732.7 | 1949.4 ± 2547.4 | 803.7 ± 2111.9 | 515.4 ± 1036.8 | 341.1 ± 620.8 | <.0001 |
| CA-3-sulfate | 2.7 ± 124.4 | 191.9 ± 328.9 | 30.5 ± 88.2 | 134.1 ± 299.5 | 33.0 ± 88.8 | 0.0002 |
| **Secondary conjugated** | **Donor** | **rCDI - IBD** | **rCDI + IBD** | **UC – rCDI** | **CD – rCDI** | **P-value** |
| G-DCA | 40.3 ± 60.6 | 2.5 ± 45.9 | 2.5 ± 238.1 | 2.5 ± 52.9 | 2.5 ± 0.0 | 0.0034 |
| **Secondary unconjugated** | **Donor** | **rCDI -IBD** | **rCDI + IBD** | **UC – rCDI** | **CD – rCDI** | **P-value** |
| DCA | 5722.2 ± 4678.1 | 2.6 ± 583.9 | 2.6 ± 5491.2 | 4771.9 ± 2523.6 | 247.7 ± 759.7 | <.0001 |
| LCA | 10448.3 ± 7510.3 | 3.5 ± 642.0 | 3.5 ± 7034.7 | 7166.1 ± 9287.4 | 114.3 ± 1452.7 | <.0001 |
| UDCA | 57.4 ± 302.5 | 2.5 ± 199.3 | 60.1 ± 599.7 | 70.5 ± 372.3 | 133.9 ± 270.8 | 0.4932 |
| iso-CDCA | 2.5 ± 39.8 | 35.6 ± 130.2 | 2.5 ± 36.7 | 2.5 ± 0.0 | 126.4 ± 477.9 | 0.0101 |
| iso-DCA | 1866.3 ± 1364.6 | 3.4 ± 37.4 | 3.4 ± 706.2 | 1226.7 ± 933.9 | 54.3 ± 252.3 | <.0001 |
| iso-LCA | 1015.5 ± 1127.6 | 2.7 ± 71.0 | 2.7 ± 773.1 | 480.7 ± 750.4 | 2.7 ± 40.7 | <.0001 |
| isoallo-LCA | 35.9 ± 119.6 | 2.5 ± 0.0 | 2.5 ± 0.0 | 2.5 ± 32.2 | 2.5 ± 0.0 | <.0001 |
| 3-oxo-DCA | 1307.0 ± 1041.2 | 4.5 ± 359.5 | 4.5 ± 2218.5 | 931.4 ± 1401.7 | 4.5 ± 69.1 | <.0001 |
| 3-oxo-CA | 2.5 ± 26.1 | 36.6 ± 289.7 | 2.5 ± 26.2 | 2.5 ± 31.3 | 77.6 ± 124.7 | 0.0013 |
| 3-oxo-LCA | 1018.1 ± 1343.3 | 2.5 ± 281.5 | 2.5 ± 902.0 | 280.8 ± 933.7 | 2.5 ± 22.6 | <.0001 |
| UCA | 28.8 | 32.2 | 104.0 | 52.1 | 114.7 | 0.3837 |

| | | | | | | |
|---|---|---|---|---|---|---|
| | ± 178.4 | ± 2087.8 | ± 2372.7 | ± 737.2 | ± 118.6 | |
| 3-oxo-CDCA | 2.5 | 40.1 | 2.5 | 2.5 | 119.8 | 0.0002 |
| | ± 0.0 | ± 362.3 | ± 0.0 | ± 0.0 | ± 476.5 | |
| DCA-3-sulfate | 2.5 | 2.5 | 2.5 | 921.9 | 55.9 | 0.7952 |
| | ± 644.0 | ± 281.6 | ± 2040.9 | ± 2205.7 | ± 338.2 | |
| LCA-3-sulfate | 68.0 | 189.9 | 48.0 | 47.5 | 40.5 | 0.7004 |
| | ± 363.2 | ± 302.7 | ± 1316.0 | ± 1046.3 | ± 460.1 | |
| UCDA-3-sulfate | 2.5 | 439.9 | 59.9 | 212.3 | 74.5 | 0.0008 |
| | ± 266.9 | ± 786.9 | ± 844.0 | ± 624.8 | ± 74.7 | |
