## Supplemental Table S16 for "Integrated metagenomic and bile acid metabolomic analysis of human fecal microbiota transplantation for recurrent Clostridioides difficile and/or inflammatory bowel diseases"

**S16 Table. Post-hoc Dunns test P-values with Benjamini–Hochberg adjustments, for 19 pre-FMT median BA metabolite levels in the four recipient groups compared to Donors (see S15 Table)**

| BA | rCDI – IBD | rCDI + IBD | UC – rCDI | CD – rCDI |
|---|---|---|---|---|
| **Primary conjugated** | | | | |
| T-CA | 0.5366 | **0.0027** | 0.8094 | 0.7684 |
| G-CDCA | **0.0071** | **0.0038** | 0.6696 | 0.7098 |
| G-CA | **0.0304** | **0.0056** | 0.8355 | 0.8846 |
| **Primary unconjugated** | | | | |
| CDCA | **0.0071** | 0.2396 | 0.7435 | 0.0845 |
| CA | **<.0001** | 0.0754 | 0.9117 | **0.0340** |
| CDCA-3-sulfate | **<.0001** | 0.1941 | 0.4037 | 0.4904 |
| CA-3-sulfate | **<.0001** | 0.9631 | 0.1090 | 0.9661 |
| **Secondary conjugated** | | | | |
| G-DCA | **0.0128** | 0.5268 | 0.1825 | **0.0343** |
| **Secondary unconjugated** | | | | |
| DCA | **<.0001** | **0.0153** | 0.5050 | **0.0070** |
| iso-CDCA | 0.1283 | 0.9285 | 0.4617 | **0.0156** |
| LCA | **<.0001** | **0.0036** | 0.4413 | **0.0042** |
| iso-DCA | **<.0001** | **0.0017** | 0.4614 | **0.0022** |
| iso-LCA | **<.0001** | **0.0111** | 0.2232 | **0.0004** |
| isoallo-LCA | **<.0001** | **0.0036** | 0.0867 | **0.0029** |
| 3-oxo-DCA | **<.0001** | **0.0385** | 0.7059 | **0.0005** |
| 3-oxo-CA | **0.0048** | 0.9269 | 0.8699 | **0.0382** |
| 3-oxo-LCA | **<.0001** | **0.0162** | 0.1627 | **0.0004** |
| 3-oxo-CDCA | **0.0060** | 0.3418 | 0.7394 | **0.0146** |
| UDCA-3-sulfate | **0.0002** | 0.3788 | 0.3962 | 0.5887 |
