## Supplemental Table S17 for "Integrated metagenomic and bile acid metabolomic analysis of human fecal microbiota transplantation for recurrent Clostridioides difficile and/or inflammatory bowel diseases"

**S17 Table. Estimated coefficient (95% CI) with baiE cpm as covariate and baiCD cpm as outcome in linear mixed model treating FMT group as clustering effect.**

| | ***baiCD* cpm 80% identity** | | ***baiCD* cpm 50% identity** | |
|---|---|---|---|---|
| | Estimated coefficient (95% CI) | P-value | Estimated coefficient (95% CI) | P-value |
| ***baiE* cpm 80% identity** | 5.01 (4.65, 5.36) | <.0001 | 1.18 (0.69, 1.68) | <.0001 |
| ***baiE* cpm 50% identity** | 17.72 (12.77, 22.66) | <.0001 | 8.31 (5.49, 11.12) | <.0001 |
