## Supplemental Table S18 for "Integrated metagenomic and bile acid metabolomic analysis of human fecal microbiota transplantation for recurrent Clostridioides difficile and/or inflammatory bowel diseases"

**S18 Table. Estimated coefficient (95% CI) with *R. gnavus* 3α-HSDH/*E. lenta* 3α-HSDH/*E. lenta* 3β-HSDH cpm as covariate and *R. gnavus* 3β-HSDH/*E. lenta* 3α-HSDH cpm at 80% identity as outcome in linear mixed model treating FMT group as clustering effect.**

| | ***R. gnavus 3β-HSDH*** **cpm** | | ***E. lenta 3α-HSDH*** **cpm** | |
|---|---|---|---|---|
| | Estimated coefficient (95% CI) | P-value | Estimated coefficient (95% CI) | P-value |
| ***R. gnavus 3α-HSDH*** **cpm** | 0.89 (0.79, 0.99) | <.0001 | 0.06 (0.02, 0.09) | 0.0014 |
| ***E. lenta 3α-HSDH*** **cpm** | 1.09 (0.12, 2.06) | 0.0288 | - | - |
| ***E. lenta 3β-HSDH*** **cpm** | 0.88 (-0.14, 1.90) | 0.0882 | 0.97 (0.90, 1.04) | <.0001 |
