## Supplemental Table S19 for "Integrated metagenomic and bile acid metabolomic analysis of human fecal microbiota transplantation for recurrent Clostridioides difficile and/or inflammatory bowel diseases"

**S19 Table. Correlation between *B. dorei* 3β-reductase, 5β-reductase and 3β-HSDH.** The estimated coefficients were based on linear mixed models with 1st gene as outcome and 2nd gene as covariate treating FMT group as clustering effect.

| | **Estimated coefficient (95% CI)** | **P-value** |
|---|---|---|
| **3β-HSDH & 5α-reductase** | 0.29 (0.25, 0.33) | <.0001 |
| **3β-HSDH & 5β-reductase** | 1.04 (0.99, 1.09) | <.0001 |
| **5β-reductase & 5α-reductase** | 2.51 (2.25, 2.78) | <.0001 |
